# DELTA: Fortifying Human Biological Resilience with an N=1 Digital Health and Dynamic Biomarker Protocol

**DOI:** 10.64898/2026.02.10.26345969

**Authors:** Peter Wang, Nigel Foo, Chengxun Su, Nicole Yong Ting Leung, Shang Wei Song, Gyula Seres, Yoann Sapanel, Lissa Hooi, Avelle Wong, Yoong Hun Ong, Poonam Rai, Hyunjin Park, Han Shi Jocelyn Chew, Laureen Yi Ting Wang, Jonathan Wei Jie Lee, Xavier Tadeo, Dean Ho

## Abstract

Societies are aging rapidly in parallel with the increasingly earlier onset of serious diseases in younger populations. These and other factors are creating a substantial disparity between healthspan, the period of life where an individual is free from serious chronic disease or disability, and lifespan — expanding the morbidity span. Extending healthspan has thus become a major priority. To pursue an integrated strategy toward healthspan support, we launched DELTA, a prospective, open-label, interventional, and participatory N=1 study (NCT06630637) conducted on a healthy individual (DELTA001, author D.H.). The study was conducted with methodological rigor to support repeatability, transparency, and balanced reporting. The core focus of the DELTA protocol was to assess and enhance human biological resilience. This was demonstrated through the subject’s adaptive capacity, revealed through changes and trajectories in cardiometabolic and pleiotropic biomarker levels based upon systematically administered challenges (e.g., fasting). Specifically, the interventional DELTA protocol integrates time-restricted eating (TRE, fasting), strength and cardiovascular fitness regimens, a Mediterranean-inspired dietary protocol, and supplementation alongside an analytics and reporting framework comprised of artificial intelligence (AI), digital health, and wearables-based sleep performance monitoring, microbiome assessment, and longitudinal tracking of biomarker dynamics and performance outcomes. This study introduces new methods and metrics for assessing these biomarker dynamics, including the development of digital biomarkers that reflect dynamic human functional resilience. Findings from DELTA may actionably guide the design of larger participatory human trials to monitor biomarker resilience, design appropriate interventions for dynamic administration, and subsequently fortify healthspan at a population level.

## INTRODUCTION

On a global scale, human lifespan continues to increase[1, 2]. However, extending human healthspan – the duration of life lived free from serious chronic disease – continues to be a challenge. Globally, the mean healthspan-lifespan gap is 9.6 years, while the US presents 12.4 years[3]. Increasing healthcare costs and reducing productivity, the widening gap poses a threat to a sustainable healthcare system[4]. Studies exploring healthspan improvement have taken diverse approaches, from preventive health measures to address chronic diseases and lifestyle management (e.g., sleep, diet, and exercise), to exploring biological mechanisms underlying aging processes, as well as pharmacologic and supplement-based geroprotection[5–65]. Unfortunately, as the field of medicine attempts to enact more pre-emptive measures to prevent the onset of chronic disease, relying on human compliance to prevent illness (e.g., “well care”) over conventional treatment that is implemented after a diagnosis (e.g., “sick care”) will be particularly challenging[66–69]. With the emergence of artificial intelligence (AI) and digital medicine alongside improved access to health monitoring methods, new approaches encompassing wearables and participatory health may potentially improve the frameworks with which humans can pre-emptively improve their healthspan. Among the approaches being explored to improve healthspan, digital solutions such as wearables have gained substantial traction due to longitudinal monitoring capabilities. From sleep, performance, to glucose and other markers, the field of digital health has democratized health insights and may also be harnessed for participatory routes to support sustained behavior change and efficacy of interventions.

In the clinical setting, healthcare decisions are still commonly made using snapshots of information, such as an annual blood test, which often includes the lipid panel, used frequently with cardiovascular risk calculators for screening cardiovascular health[70, 71]. This panel examines low-density lipoprotein cholesterol (LDL-C, commonly coined as “bad cholesterol”), high-density lipoprotein (HDL, or “good cholesterol”), triglycerides (TG), and total cholesterol levels. Despite serving as an evidence-based standard for patient management, static measurements may overlook lipid dynamics that can potentially uncover new insights into health status.

Longitudinal blood testing for preventive health beyond the annual health screen is rarely implemented due to logistical and economic constraints. Also, it is unclear which longitudinal information may be actionable toward refining treatment protocols. In particular, there is a scarcity of insight into the dynamics, or timescale of change and/or recovery to baseline levels for individual biomarkers in response to intervention. Furthermore, prior studies have shown that subjects evolve over time, with optimal drug dosing and other interventions being modulated dynamically[72–75]. Even drug synergy is a dynamic process that is dose-, time-, and patient-dependent[76–78]. However, it is established that biomarker response to therapy changes over time, the timescale of biomarker changes and recovery in response to interventions can both serve as potential biomarkers. For example, monitoring biomarker elevation to established interventions (e.g., pharmacologic, lifestyle, etc.) and subsequent return to baseline levels can offer insight into biological resilience for cardiometabolic, inflammatory, and other functions. Studies that examine these dynamics may lead to novel resilience biomarkers.

In addition to these considerations for longitudinal testing, it is important to determine which biomarkers may be suitable for resilience testing. Emerging studies have also explored apolipoprotein B (ApoB), the primary protein component of atherogenic lipoproteins (e.g., LDL), due to its potential for more accurately predicting cardiovascular disease than LDL as it directly reflects the number of cholesterol-bearing particles that can drive atherosclerosis[79–81]. In addition to serving as a potential predictor of cardiovascular disease, ApoB may also reveal key insights into lipoprotein metabolism in response to time-restricted eating (TRE) or fasting.

To assess interventions and longitudinal biomarker trajectories that may impact healthspan, we report the results of DELTA, a prospective, open-label, and interventional N=1 study conducted on a healthy individual (DELTA001) (Fig. S1). The rationale for conducting this open-label, unblinded, and participatory N=1 trial (DELTA001; author D.H.) was to personalize the DELTA regimen to DELTA001 with high adherence and participant awareness of each intervention’s (e.g., fasting, diet, fitness, sleep, etc.) role in fortifying healthspan. Moreover, methodological rigor was ensured with biological replicates to confirm the dynamics observed in this study. Additionally, DELTA001’s prior demonstration of metabolic switching in response to a health intervention provides a strong basis for interventionally supporting metabolic resilience during the current DELTA regimen[82]. By ensuring rigorous and detailed data collection, this N=1 study generated evidence that was directly relevant to individual health management, and may also help inform the design of future health optimization studies at the population level. The DELTA study was conducted using dynamic interventions such as TRE and coupled with longitudinal digital health and clinical biomarker measurement. Cardiometabolic health, sleep performance, microbiome dynamics, and performance metrics served as the primary outcomes of DELTA. Biomarkers were selected due to their evidence-based relevance as indicators of cardiometabolic and brain health, systemic inflammation, and increasing interest as predictors of healthspan. In addition, while these markers are also receiving broader attention in population-level studies in seniors, their profiling in young adults is less common, and the development of resilience biomarkers based upon the timescales of their dynamics in response to interventions may be important for addressing healthspan at a population scale. To provide ongoing feedback from biomarker monitoring in the context of healthspan impact, a custom generative pre-trained transformer (GPT)-based Healthspan Copilot was developed to assess biological age as an exploratory means of gamification toward regimen adherence. This work sought to derive potential population-relevant learnings toward possible wider-scale studies into biomarker development, gamification, and other factors that can contribute toward positive impact on healthspan outcomes.

The DELTA study and protocol are outlined in Fig. 1. The DELTA regimen comprised of a feeding window of 4 hours (minimum of 20-hour fast). Meals were typically consumed between 11am and 3pm, with limited exceptions for travel and professional obligations. This timeframe was also developed to provide sufficient time for digestion prior to sleep. For biomarker resilience and flexibility testing, as well as acute microbiome dynamics analysis, the primary intervention was a 48-hour water-only fast (no calories consumed, no supplementation) with serum biomarker readings taken before and after the fast. Daily physical activity was undertaken, with a primary focus on strength training (one focus muscle group per day), with 2-3 cardiovascular training events 3 times per week in addition to daily strength training (Norwegian protocol high-intensity interval training (HIIT)). Importantly, the strength training was classified between “Light” and “All Out” on the WHOOP wearable, with a majority of sessions classified between “Moderate” and “Strenuous” to prevent injury. The dietary regimen was Mediterranean-inspired, consisting of lean protein (fish, chicken, chickpea), leafy greens, red cabbage, extra virgin olive oil (no other oils were used for cooking), a daily blended shake (blueberries, red cabbage, broccoli, ginger), and pudding (unsweetened almond milk, chia seeds, flaxseeds, psyllium husk, and organic/unsweetened cacao, monkfruit sweetener) (Fig. 1B and Supplementary File 1). A comprehensive supplement regimen was also taken (Table S1). The general DELTA protocol is summarized in Fig. 1, and the key takeaways from this study are outlined in Fig. 1C. Summaries of food and supplement intake are shown in Figs. S2-S5, Table S1, and Supplementary File 1.

**Fig. 1.**
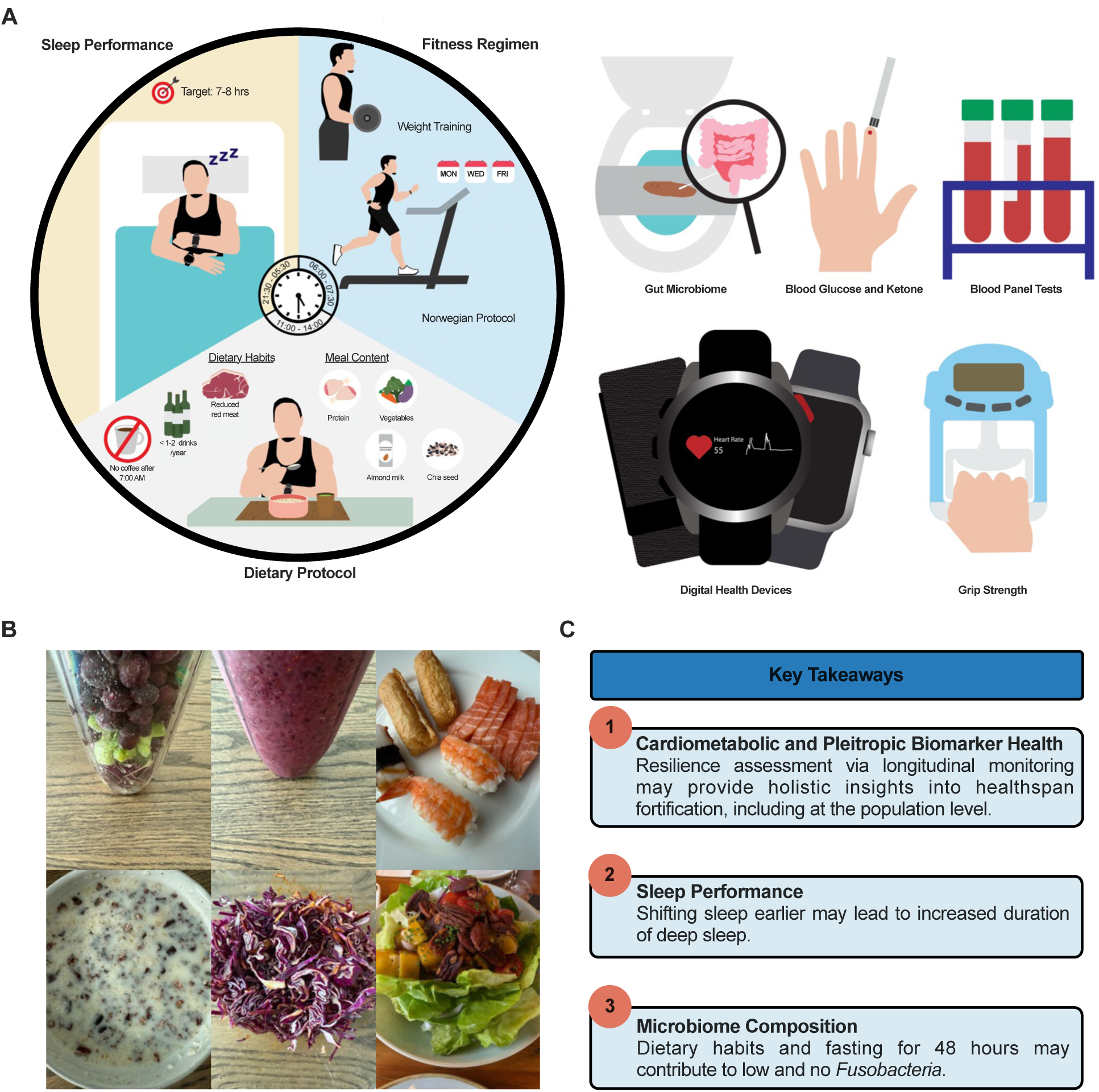
DELTA protocol. **(A)** Schematic summary of the DELTA protocol comprising of dietary protocol, fitness regimen, sleep schedule, gut microbiome and blood tests, sleep and fitness performance evaluation, and usage of digital health devices. **(B)** Representative food pictures. Photo library of DELTA001’s dietary regimen can be found in Supplementary File 1. **(C)** Key takeaways from this study.

The DELTA protocol was developed to render biomarkers (e.g., ketones, glucose, ApoB, ApoA, homocysteine) in a labile state such that their dynamics could be straightforwardly modulated based on interventions that range from straightforward (e.g., single set of push-ups), to challenging (e.g. 48-hour water fast) (Table S2). As such, while the absolute value of the biomarkers was monitored to assess for outcomes, the foundation of the DELTA protocol was to observe the magnitude of biomarker change, rate of change, and related factors to longitudinally reveal meaningful feedback in response to healthspan-supporting interventions. For example, subject DELTA001 previously undertook an N=1 study to understand the role of biomarker dynamics and frequent biomarker feedback as a strategy to gamify adherence to healthspan-supporting activities[82]. This first study pertained only to 2 biomarkers (ketone and glucose dynamics). The DELTA trial sought to markedly expand this protocol to address multiple pillars associated with healthspan outcomes, including cardiometabolic, neurological, microbiome, sleep, and performance assessment (Fig. 1).

The biomarkers selected for this study included ApoB, apolipoprotein A (ApoA), high-sensitivity C-reactive protein (hs-CRP), homocysteine, blood glucose, and blood ketones (Table S2). ApoB was selected due to its emerging promise as a superior marker over LDL-C to assess cardiovascular risk as well as an indicator of atherogenic particle burden[83–86]. ApoA was selected due to its accurate indication of HDL function, cardiovascular protection, and metabolic resilience[87, 88]. We selected hs-CRP as it accurately reflects systemic inflammation, which is an important driver of cardiovascular disease, inflammaging, and cognitive decline[89, 90]. Homocysteine was selected as it is a broadly applicable indicator of methylation and B-vitamin status, efficiency of detox pathways, as well as cardiovascular and neurological aging risk[85, 86]. Importantly, a number of large-scale trials have demonstrated the real-world actionability of these markers. The INTERHEART and AMORIS trials have placed ApoB above LDL-C as a stronger predictor of cardiovascular risk[79, 80]. For hs-CRP, the JUPITER trial, among others, demonstrated its ability to independently predict strokes and heart attacks in patients with normal LDL. For homocysteine, the Framingham Study demonstrated that elevated levels can predict dementia, stroke, and mortality risk[91, 92].

As previously noted, for biological resilience testing, a 48-hour fast served as the primary intervention to assess the dynamics of blood glucose, blood ketones, ApoB, ApoA, and the ApoB/ApoA ratio[84]. To assess broader biological resilience in the context of neurological, methylation, cardiometabolic, inflammaging, and other systems, homocysteine and hs-CRP served as pleiotropic markers to observe stress and rebound responses. A 48-hour fast also served as the primary intervention to assess biomarker resilience, with resulting changes observed, and an additional reading was conducted to assess the return to baseline. For sleep performance optimization, the primary intervention, alongside the multiple facets of the DELTA protocol, was a sleep phase advance/circadian intervention of moving the sleep time earlier by approximately 2.5 hours (9:15pm to bed/asleep by 9:45pm) compared to the control sleep time (12:00am). Systematic efforts were also made by DELTA001 to adapt to multiple time zones during international travel. Sleep duration, stages, time awake, scoring, heart rate variability (HRV), resting heart rate (RHR), and other relevant markers were monitored with a WHOOP 4.0, Garmin Epix Pro 2, and Apple Watch 9. Physical performance assessment was conducted via run speed, grip strength, and cardiovascular assessment spanning blood and pulse pressure. The 48-hour water fast also served as the primary intervention for the microbiome dynamics portion of the study, with samples taken prior to and at the conclusion of the fast. Next-generation sequencing was used to assess gut microbe composition changes.

Overall, the DELTA study provides insights into biomarker dynamics and resilience, sleep and physical performance as part of a combinatorial protocol consisting of TRE, fitness, and rationally-designed dietary regimen. Cardiometabolic flexibility, sleep, microbiome dynamics, and performance outcomes were assessed, with evidence of dose-dependent biomarker responses, exceptional cardiometabolic and pleiotropic biomarker resilience, marked sleep performance improvement, and platforms for longitudinal monitoring adherence. While this is an N=1 study, potentially generalizable frameworks for population-level investigation were observed.

## RESULTS

### Metabolic Resilience and Flexibility

In this portion of the study, high-resolution insight into metabolic resilience and flexibility were studied, with blood glucose and blood ketones readings taken when both markers were highly and DELTA001 was metabolically flipped in ketosis, at approximately 40-42 hours of fasting (Figs. 2 and S6-S13). In these studies, DELTA001 performed weightlifting, with biomarker readings taken before exercise start, and continuously in 5-20-minute intervals until exercise completion to monitor biomarker dynamics during glycolysis and recovery (Fig. 2A and S6-S8). Three segments are noted in Fig. 2A. Segment 1 represents exercise start. Segment 2 represents the peak of the glucose ketone index (GKI), a reflection of the state of ketosis. Segment 3 represents ketotic recovery. Of note, rapid glycogenesis and exit from ketosis were observed immediately following the start of exercise (Fig. 2A, Segment 1), with glucose levels rising and ketone levels dropping, indicating rapid substrate switching in response to physical stress. Post-exercise, we observed sustained ketone rebound (Fig. 2A Segment 2), with levels rising continuously for at least ∼100 minutes post-exercise before reaching a plateau (Fig. 2A, Segment 3). Notably, we observed that ketone level during the exercise-recovery period followed a consistent second-order polynomial pattern (Fig. S14).

**Fig. 2.**
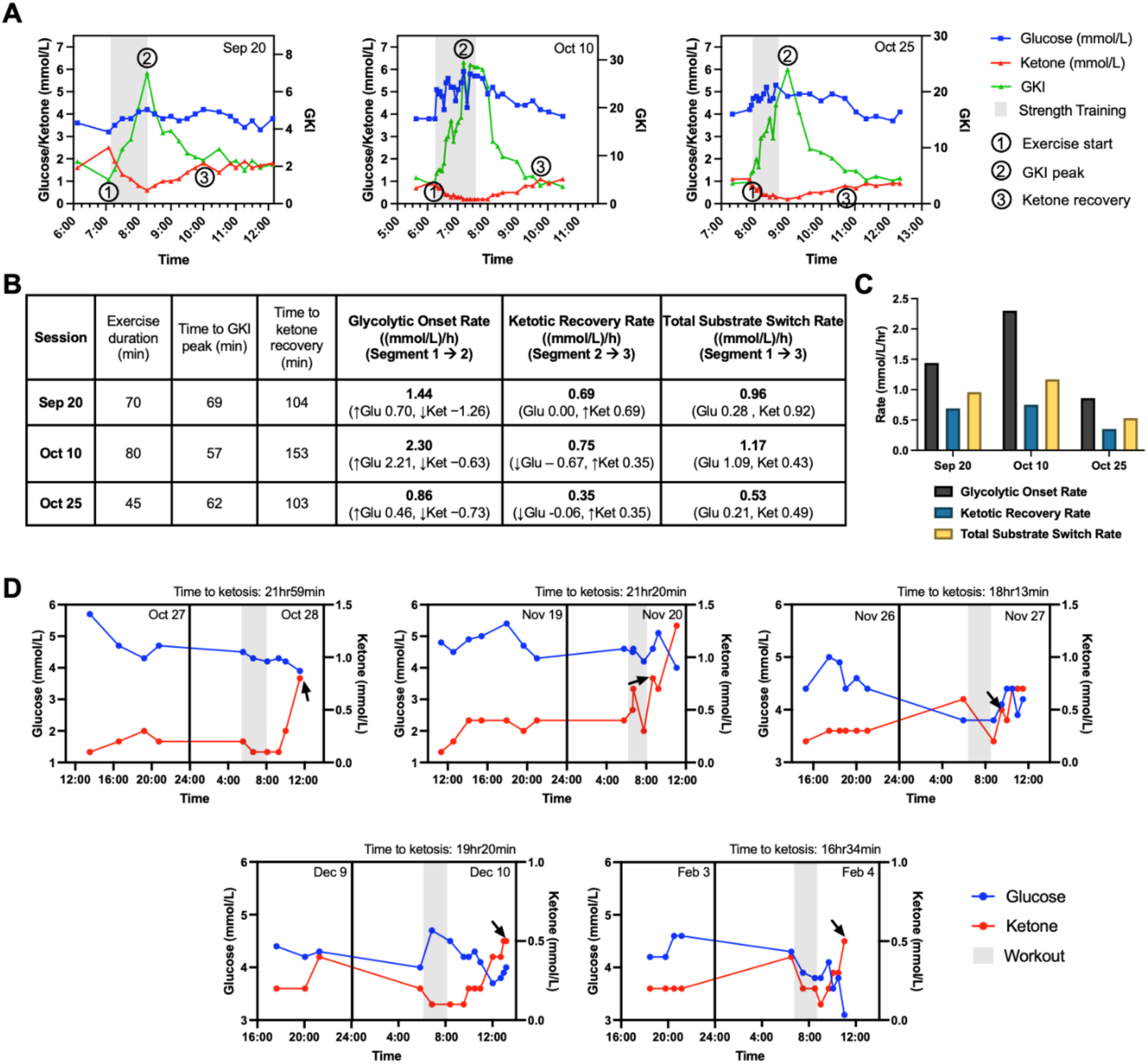
Metabolic flexibility. **(A)** Glucose-ketone dynamics during the 48-hour fasting strength training regimen. All sessions were performed on the 2nd day of a 48-hour fast. **(B)** Analysis of metabolic switches for the 48-hour fasting strength training regimen based on changing rates of glucose and ketone. **(C)** The glycolytic onset rate, ketotic recovery rate, and total substrate switch rate during the 48-hour fasting strength training regimen. **(D)** Glucose-ketone dynamics during the one-meal-a-day (OMAD) ketosis entry regimen. The workout consists of yoga, weightlifting, and intermittent sessions of walking and running. The first measurement on day 1 was taken immediately after meal consumption. Black arrow indicates post-exercise ketosis state (ketone level ≥ 0.5 mmol/L). Source data can be found in Figs. S6-S13.

Asymmetric substrate switching kinetics were observed across the three sessions outlined in Fig. 2A, where ketone decline during exercise consistently exceeded the rate of post-exercise ketotic recovery, indicating rapid exit from ketosis under physical stress and moderate re-entry during recovery (Fig. 2B). Of note, a dose-dependent correlation of glycolytic onset rate, ketotic recovery rate, and total substrate switch rate were observed (Fig. 2C). More specifically, these 3 rates are positively correlated with the exercise duration, indicating the potential of this metric as a metabolic resilience biomarker.

The speed of metabolic switching across single cycles of OMAD ketosis entry sessions was monitored (Figs. 2D and S9-S13). In total, 5 independent sessions revealed initial switches into ketosis between 20 hours to a recent reading of approximately 16.5 hours. No claims are made in this study with regards to gradually increasing rates of metabolic switch. However, metabolic adaptation, metabolic memory, and physiological priming toward increased metabolic switch efficiency has been previously documented[93–95]. Furthermore, similar to the trajectories observed in Fig. 2A, ketone level during the exercise-recovery period in Fig. 2D also followed a consistent second-order pattern (Fig. S14).

### Cardiometabolic and Pleiotropic Markers

The fasting dynamics of ApoB, ApoA, ApoB/ApoA ratio, homocysteine, and hs-CRP were assessed as a function of pre-48-hour fasting (PRE48), post-48-hour fasting (POST48) and unfasted states that were measured as part of a 3-meal-a-day regimen (3MAD) (Fig. 3A-C and Supplementary Data 1). Please note that for the purposes of comparison between groups, data sets are repeated across different subfigures. Fig. 3A examines post fasting levels against unfasted controls, POST48 ApoB was 84±3.1 mg/dL and 3MAD ApoB was 74±2.7 mg/dL. POST48 ApoA was 17±4.9 mg/dL and 3MAD ApoA was 167±2.5 mg/dL. The POST48 ApoB/ApoA ratio was 0.48±0.02 and 3MAD ApoB/ApoA ratio was 0.44±0.02. A statistically significant reduction in ApoB was observed when comparing POST48 ApoB with 3MAD ApoB, while visible but non-significant reductions in ApoA and ApoB/ApoA ratio were observed.

**Fig. 3.**
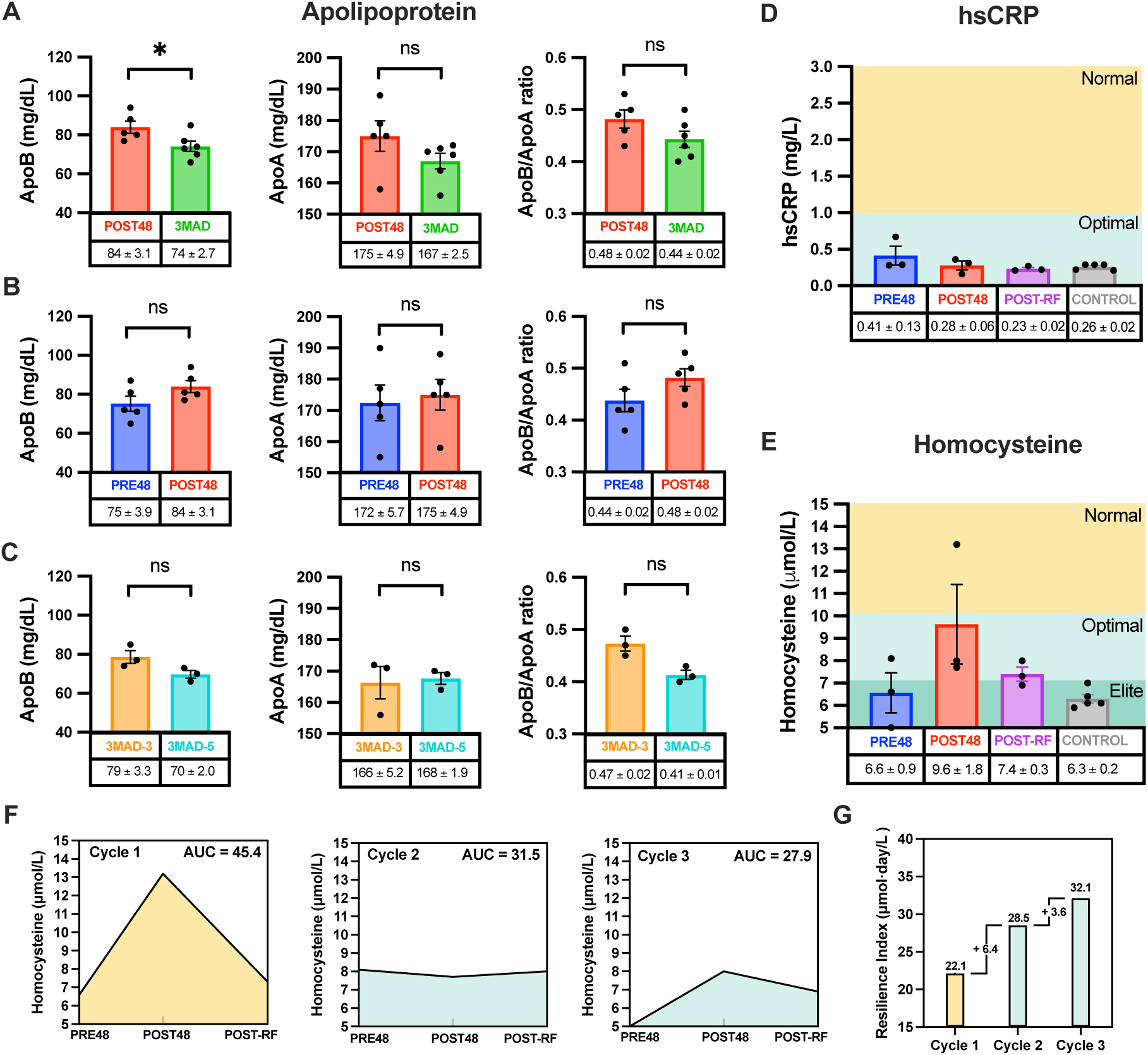
Cardiometabolic health biomarkers. **(A)** Fasted and unfasted apolipoprotein levels. Fasted measurements were obtained after a 48-hour fast (POST48), while unfasted measurements were collected on either day 3 or day 5 during an intervention period of a 3-meals-a-day (3MAD) regimen. **(B)** Apolipoprotein levels before and after a 48-hour fast (PRE48 vs. POST48). **(C)** Apolipoprotein levels on day 3 and day 5 of a 3MAD regimen (3MAD-3 vs. 3MAD-5). Data points in the POST48 group were used for analysis in both PRE48 vs. POST48, and POST48 vs. 3MAD. Data points in the 3MAD-3 and 3MAD-5 groups were pooled into one group (3MAD) for analysis in POST48 vs. 3MAD. Statistical significance between groups was assessed using non-parametric t-tests (* represents p<0.05, ‘ns’ represents non-significant). **(D)** Hs-CRP levels measured before, after a 48-hour fast, and after 2-3 days of refeeding (PRE48 vs. POST48 vs. POST-RF). The normal range is indicated as 1.0-3.0 mg/L, and the optimal (low-risk) range is indicated as below 1.0 mg/L. **(E)** Homocysteine levels measured before, after a 48-hour fast, and after 2-3 days of refeeding. The normal range is indicated as 10-15 *μ*mol/L, the optimal (low-risk) range is indicated as 7-10 *μ*mol/L, and the elite range is indicated as 5-7 *μ*mol/L. The refeeding period and control samples both follow a one-meal-a-day regimen. **(F)** Expanded analysis of homocysteine dynamics across three cycles. Shaded areas represent the AUC of homocysteine across PRE48, POST48, and POST-RF, expressed in *μ*mol·day/L. **(G)** Resilience index computed from homocysteine dynamics across three cycles. Yellow (cycle 1) indicates a normal response, while green (cycle 2 and 3) indicates an optimal response. Data in (F, G) are derived from homocysteine levels shown in (E). Statistical significance between groups was assessed using non-parametric one-way ANOVA. Only statistically significant comparisons are annotated. Data is represented by mean ± standard error of mean (SEM). Source data can be found in Supplementary Data 1.

Fig. 3B examines PRE48 and POST48 differences in ApoB, ApoA, and ApoB/ApoA ratio. PRE48 ApoB was 75±3.9 mg/dL, and POST48 ApoB was 84±3.1 mg/dL. PRE48 ApoA was 172±5.7 mg/dL and POST48 ApoA was 175±4.9 mg/dL. The PRE48 ApoB/ApoA ratio was 0.44±0.02 and POST48 ApoB/ApoA ratio was 0.48±0.02. Visible but non-significant increases in ApoB, ApoA, and ApoB/ApoA ratios were observed.

Fig. 3C assessed ApoB changes between day 3 (3MAD-3) and day 5 (3MAD-5), visible but non-significant reductions were observed. The same was observed for ApoB/ApoA ratio for 3MAD-3 and 3MAD-5. The 3MAD-3 ApoB was 79±3.3 mg/dL and 3MAD-5 ApoB was 70±2.0 mg/dL. The 3MAD-3 ApoA was 16±5.2 mg/dL and 3MAD-5 ApoA was 168±1.9 mg/dL. The 3MAD-3 ApoB/ApoA ratio was 0.47±0.02 and 3MAD-5 ApoB/ApoA ratio was 0.41±0.01.

It should be noted that baseline ApoB values, represented by the PRE48 values, were within the optimal range of 60-80 mg/dL. A temporary rise of POST48 ApoB levels was observed and return to baseline as an indicator of metabolic flexibility and lipoprotein clearance will be discussed (Fig. 3B). Interestingly, some unfasted ApoB readings were observed to be near the lower limit of the normal ApoB range (65mg/dL), and included a reading of 65mg/dL, 66 mg/dL, and 70 mg/dL, potentially signifying efficient ApoB metabolism. The unfasted dynamics of ApoB, ApoA, and ApoB:ApoA ratio levels did not result in the rise that was seen during the fasting dynamics study (Fig. 3C).

hs-CRP levels PRE48, POST48, post-refeeding (POST-RF), and during the baseline regimen (CONTROL) were recorded (Fig. 3D and Supplementary Data 1). PRE48 levels were 0.41±0.13 mg/L, POST48 levels were 0.28±0.06 mg/L, POST-RF levels were 0.23±0.02, and CONTROL levels were 0.26±0.02. Of note, no apparent increases in inflammation were observed between the PRE48 and POST48 hs-CRP levels. Of note, initial inflammation readings were assessed via C-Reactive Protein (CRP) (Supplementary Data 1). With the exception of the first 2 readings (1.0 µmol/L and 1.2 µmol/L), and another reading of 0.6, which proceeded an intense workout of over 1,500 pushups performed by DELTA001, all CRP values were <0.6 µmol/L and undetectable, which resulted in a change to hs-CRP measurements.

Homocysteine levels were also monitored under the same conditions (Fig. 3E and Supplementary Data 1). PRE48 homocysteine was 6.6±0.9 µmol/L, POST48 homocysteine was 9.6±1.8 µmol/L, POST-RF homocysteine was 7.4±0.3 µmol/L and CONTROL homocysteine was 6.3±0.2 µmol/L.

Homocysteine resilience monitoring revealed an interesting outcome (Fig. 3E-G). Following the first cycle of 48-hour fasting, homocysteine rose from 6.6 to 13.2 µmol/L, which is considered elevated and well outside of the optimal (low-risk) levels of 10 µmol/L or below (7-10 µmol/L considered optimal)[96, 97]. However, a follow-up homocysteine test post-refeeding within the next 2-3 days revealed a return to baseline, potentially signifying strong methylation efficiency and metabolic resilience (Fig. 3F). Of note, this homocysteine resilience test was repeated for 2 additional cycles. In the first repeat, the pre-fast homocysteine level was 8.1 µmol/L. The POST48 homocysteine was 7.7 µmol/L, a markedly reduced value compared to the level observed during the first cycle. The POST-RF homocysteine recovery level was 8.0 µmol/L, effectively revealing no apparent change despite a 48-hour fast, further indicating potential methylation and metabolic adaptation (Fig. 3F). Interestingly, the 3rd cycle also revealed the potential of sustained methylation and metabolic adaptation. In this cycle, the PRE48 homocysteine was 5.0 µmol/L, the POST48 homocysteine was 8.0 µmol/L, and the POST-RF homocysteine was 6.9 µmol/L, all within optimal levels (Fig. 3F). Subsequent baseline readings of homocysteine were 6.1 µmol/L, 7.0 µmol/L, 6.4 µmol/L, 5.9 µmol/L, and 6.1 µmol/L. These values are classified as elite, signifying exceptional methylation efficiency, B-vitamin stores, and metabolic health[98, 99]. Additionally, Area Under the Curve (AUC) analysis was performed for homocysteine vs. days across the three cycles. AUC values decreased across cycles (45.4 → 31.5 → 27.9 μmol·day/L) (Fig. 3F). Using the AUC values, a resilience index (RI), defined as a quantitative measure of an individual’s ability to maintain durable homocysteine response following exposure to a stressor (TRE), was calculated as 22.1→ 28.5 → 32.1 μmol·day/L (Fig. 3G). This suggests attenuated responses to the stressor and the development of adaptive and durable methylation resilience. Please note that the AUC’s and RI were derived from homocysteine levels shown in Fig. 3E.

### General Health Outcomes

General health outcomes were assessed using overnight HRV trends (Garmin Epix Pro (Gen2)), highest 5-minute overnight HRV average (Garmin Epix Pro (Gen2)), blood and pulse pressure, and longitudinal weight assessments (Fig. 4 and Supplementary Data 1). Overnight HRV readings and baseline HRV range data showed standard day-to-day variation (red dots) with a 7-day smoothed baseline (red line) and baseline range (gray shaded region) (Fig. 4A). DELTA001’s HRV fluctuated over time with noticeable dips and increases, which potentially indicate interspersed periods of recovery (rising) and possible stress or fatigue episodes (HRV dips). The highest overnight 5-minute averages were also plotted (Fig. 4B). Some nights were not recorded due to rare instances of DELTA001 not wearing the device (forgetting to wear the device), or the device not recording for unknown reasons.

**Fig. 4.**
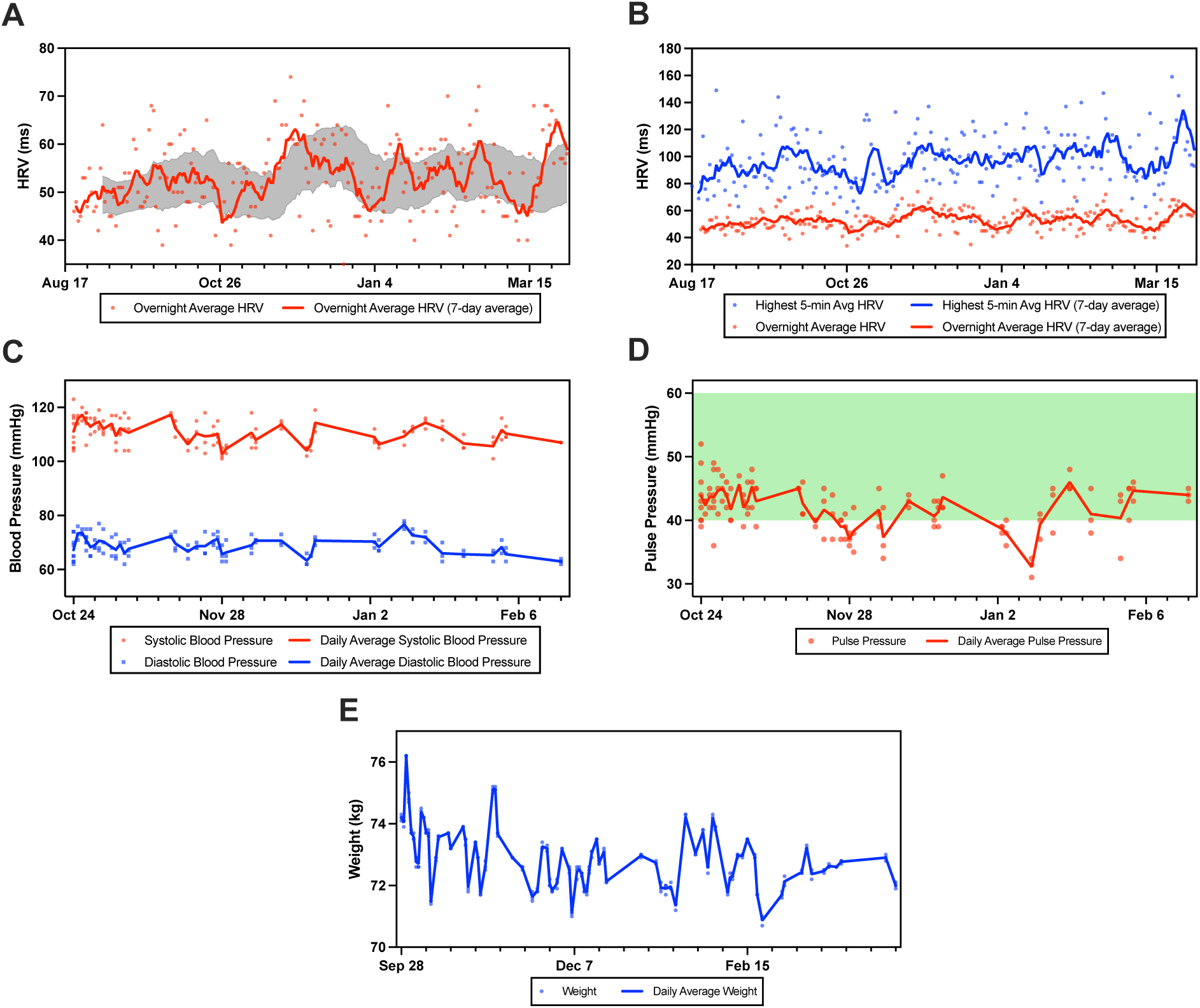
Cardiovascular health and weight. **(A)** Overnight heart rate variability (HRV) readings plotted daily (blue dots), with smoothed 7-day baseline (solid blue line), and a baseline range (shaded area between red dashed lines). **(B)** Overnight highest 5-min average HRV. **(C)** Systolic (SYS) and diastolic (DIA) blood pressure readings plotted over time, with the corresponding daily averages (solid lines) and linear trendlines (dotted lines). **(D)** Pulse pressure values (difference between SYS and DIA) are shown with individual readings and the daily average. The shaded green zone denotes the clinically healthy pulse pressure range of 40-60 mmHg. **(E)** Daily weight readings with their corresponding daily averages. Source data can be found in Supplementary Data 1.

Daily blood pressure (BP) readings were taken, with the daily average and trendlines shown (Fig. 4C and Supplementary Data 1). DELTA001 did not take any prescription anti-hypertensive or cholesterol medication. Systolic pressure (red) was shown to be generally stable (110–120 mmHg). Diastolic pressure (blue) was generally stable between 70–80 mmHg. While modestly declining trends were shown for both systolic and diastolic pressure, no claims are made causation by the DELTA protocol. Pulse pressure was also recorded longitudinally (Fig. 4D and Supplementary Data 1). DELTA001 pulse pressure (Systolic pressure-diastolic pressure) remained within the healthy range of 40–60 mmHg (green shaded area)[100], indicating generally healthy arterial compliance. Longitudinal weight readings and daily averages were taken (Fig. 4E and Supplementary Data 1). Weight fluctuates between 71 kg and 76 kg were observed, ending at approximately 72.5 kg.

### Performance Outcomes

DELTA001’s performance was assessed longitudinally with grip strength, correlation of grip strength with blood pressure, and running speed as part of a Norwegian protocol of HIIT, with 4 sets of 4-minute run cycles and 3-minute low intensity cycles. Dominant (right) and left hand measurements were taken with daily averages and standard error of the mean (Fig. 5A and Supplementary Data 1). Of note, an industrial grade grip strength meter with improved resistance against grip slippage was intermittently used toward the end of the study, with some dominant hand readings as high as 79.65kg also recorded (Fig. S15). Normalized grip strength (grip strength divided by body weight) was also plotted over time (Fig. 5B and Supplementary Data 1). The daily grip strength as well as normalized grip strength trends showed a gradually increasing trend. No claims are made with regards to the impact of the DELTA protocol on this trend. Normalized right and left grip strength was also plotted against diastolic and systolic pressure to observe correlations, which have also been previously reported in prior studies (Fig. 5C)[101]. The HIIT run times showed a gradual increase over time (Fig. 5D and Supplementary Data 1). During the course of training, the starting set of run speeds was gradually increased in order to sufficiently advance toward the highest running speed on the 4th set. This training approach of stepwise increases in training difficulty may have played a role in the performance improvement over time. We also do not discount the possibility that the gradual increase in time was due to DELTA001 not running at his absolute full capacity during each running trial.

**Fig. 5.**
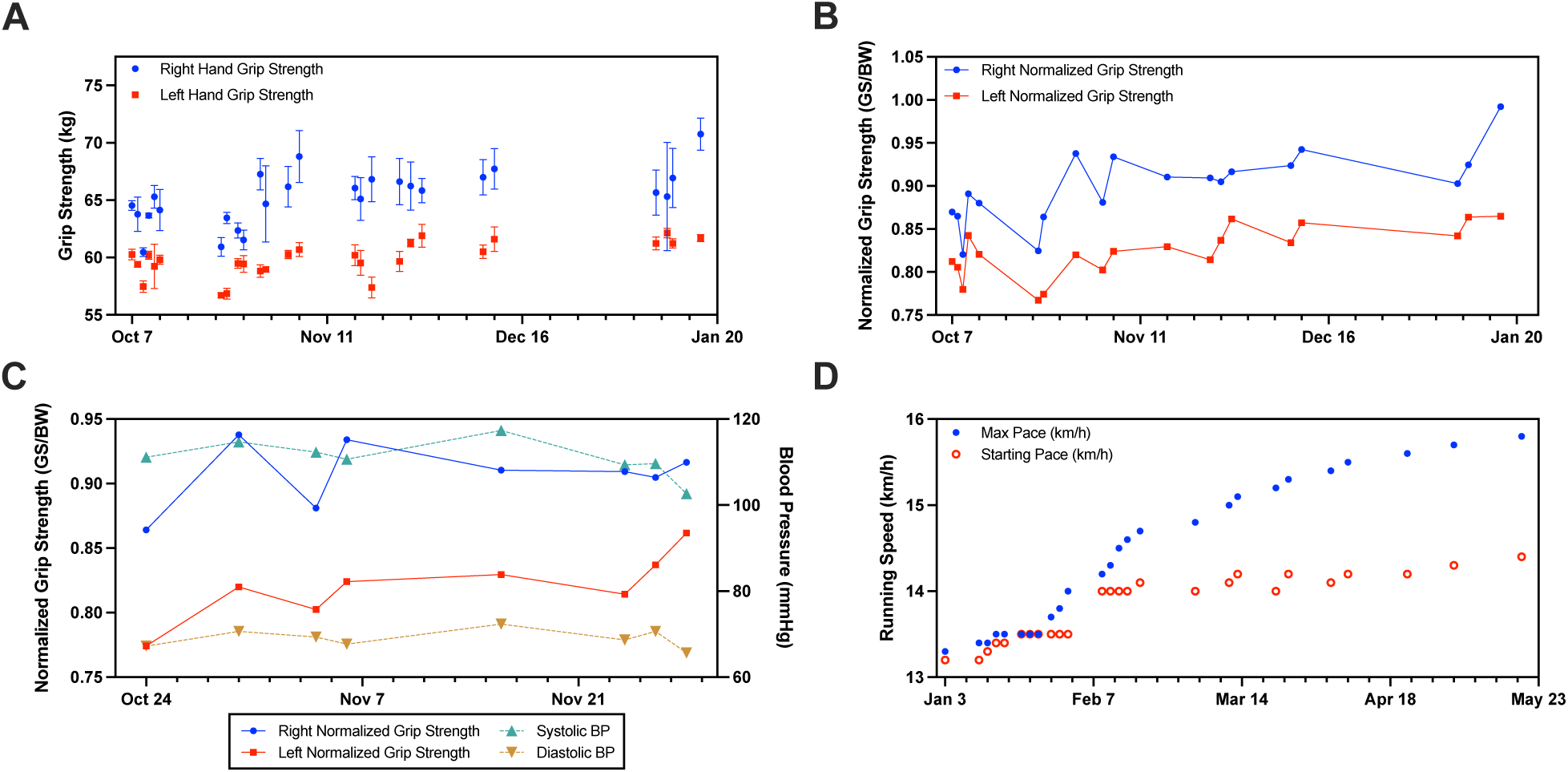
Fitness performance. **(A)** Grip strength readings for the left (green) and right (blue) hands over time, with individual measurements and daily averages where the error bars denote the SEM. **(B)** Normalized grip strength (grip strength (GS) divided by body weight (BW)) for each hand plotted over time. **(C)** Normalized grip strength (left and right) plotted alongside systolic and diastolic blood pressure readings to explore their temporal relationship. **(D)** Treadmill run performance over time, showing both maximum speed and starting speed. The subject demonstrated a consistent upward trend in both starting and maximum speed. Source data can be found in Supplementary Data 1.

### Sleep Performance

A sleep study spanning the start of DELTA through April (nearly 7 months) was conducted. The sleep intervention regimen included multiple elements. While the primary intervention was moving the sleep time earlier by approximately 2 hours and 45 minutes (from a control time of 12am midnight to 9:15pm), the DELTA regimen included consistent morning fitness sessions, a Mediterranean-based dietary regimen, and other factors that may have collectively enabled the sleep outcomes observed. These included the stoppage of caffeine intake from coffee at 7am on weekdays and 8am on weekends to allow for sufficient clearance, the elimination of afternoon naps, and the prohibition of screen time before bed. It is important to note that prior to and during the study, DELTA001 slept alone.

Unless otherwise stated, Fig. 6 contains data from the Apple Watch 9 (Supplementary Data 1). A control period of sleep undertaken by DELTA001, with sleep times of approximately 12 am midnight, was analyzed retrospectively (Fig. 6A). The sleep patterns were shown to be fragmented, with generally inconsistent sleep onset time, variability in wake-up times. Sleep architecture was suboptimal, with sparse and insufficient deep sleep, insufficient rapid eye movement (REM) sleep, and light sleep comprising a large proportion of sleep architecture. This baseline sleep regimen was comprised of irregular sleep patterns, potentially due to a lack of interventions.

**Fig. 6.**
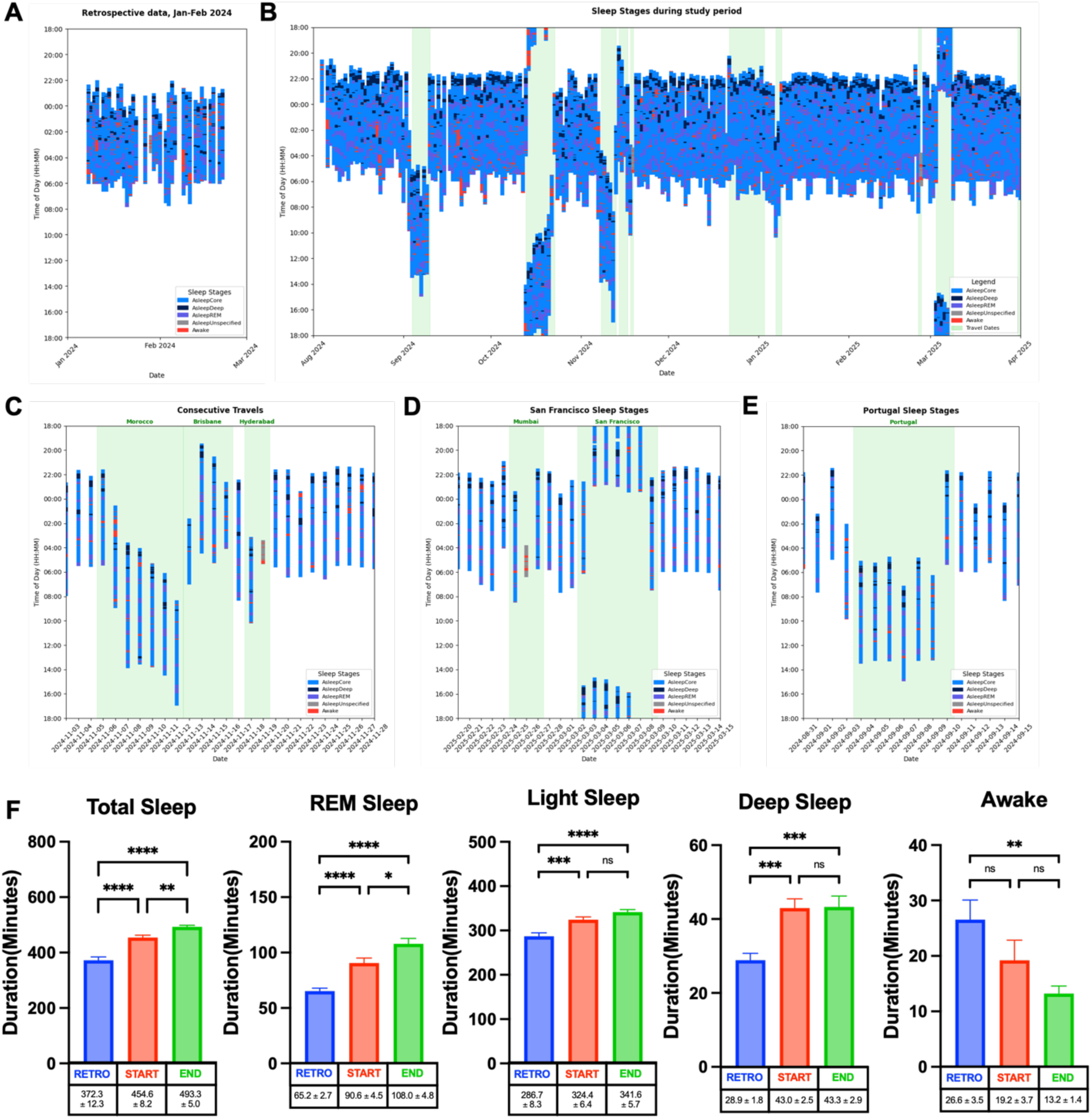
Sleep performance. **(A)** Retrospective sleep data from first quarter of 2024. **(B)** Overview of sleep data during the study from August 2024 to April 2025, where hours recorded are according to the Singapore time zone (UTC+8:00). Sleep data collected over periods of international travel are highlighted in green. **(C-E)** Sleep data during international travels with inset HRV data. **(F)** Comparison of duration of sleep stages over three distinct time points: retrospective data (RETRO), start of study (START), and end of study (END). An ordinary one-way ANOVA followed by Tukey’s multiple comparisons was used to assess differences between groups. P-values are represented as follows: p-value ≤ 0.05 (*), p-value ≤ 0.01 (**), p-value ≤ 0.001 (***), and p-value < 0.0001 (****), with “ns” representing non-significance. Data is represented by mean ± SEM. Source data can be found in Supplementary Data 1.

During the DELTA intervention period, the consistent presence of deep sleep during the early segment of the sleep architecture was clearly noted (Fig. 6B). Also, a consolidation in sleep timing was apparent after study initiation. Sleep continuity was also shown to improve compared to the control sleep period.

This study was not without sleep disturbances. DELTA001’s travel sleep performance across diverse time zones is also shown. This portion of the study represents concerted efforts to adapt to international travel. Key interventions included: (1) Using evening long-haul flights where possible; (2) Immediate sleep with eye shades and earplugs upon take-off; (3) Consolidation of take-off meals with landing meals, served as close as possible to landing; (4) Where applicable, if arriving to long-haul destinations in the morning, additional feeding to ensure sufficient caloric intake, with no eating through the rest of the day and evening in order to resume DELTA feeding window; (5) Immediate resumption of morning fitness routine. The hours recorded are presented according to the Singapore time zone (UTC+8:00), and international travel periods are highlighted by shaded green regions. Sleep data in Fig. 6 is derived from the Apple Watch 9 to visualize sleep performance. HRV data is from the Garmin Epix Pro (Gen 2) (Supplementary Data 1).

Across all three travel legs, aggregate sleep duration and architecture were generally preserved, suggesting strong sleep resilience (Fig. 6C-E). However, a key exception was the multiple short red-eye flights from Mumbai and Hyderabad to Singapore where total sleep was acutely impacted (Fig. 6C,D). Resilience and recovery from these flights are to be discussed. The first travel is represented by consecutive trips between Singapore, Morocco (via Türkiye), Australia (Brisbane), and India (Hyderabad), with return trips to Singapore between each leg (Fig. 6C). Sleep duration was generally robust, with multiple days showing 7+ hours of sleep. Sleep architecture also appeared robust, with all sleep stages consistently represented. Sleep fragmentation was visible in higher quantities across specific days but did not suggest substantial sleep disruption. Despite having a red-eye flight of short duration (< 4 hours), where a clear sleep disruption was noted, HRV remained within a balanced level, suggesting favorable autonomic balance and resilience. For this segment, despite multi-country travel across diverse time zones, sleep architecture and duration were strong.

In the subsequent sleep segment, which included travel to India (Mumbai) and San Francisco, the sleep duration was consistent, averaging 7-8 hours of sleep per night (Fig. 6D). Of note, the sleep stages were exceptionally well distributed, with strong sleep architecture and patterns of deep and REM sleep. Importantly, sleep fragmentation was minimal, which represented consolidated and high-quality sleep. It should be noted, however, that a period of sustained HRV suppression was observed following the return to Singapore from San Francisco. It is possible that this was due to the exceptional sleep experienced while in San Francisco, suggesting that while DELTA001 was able to maintain strong sleep adaptivity across time zones, exceptional sleep while on travel may result in delayed restoration of autonomic balance, resulting in a temporary HRV imbalance. This may also demonstrate that in some cases, “jet lag” is unavoidable.

The third sleep segment included travel between Singapore and Portugal (Fig. 6E). Again, sleep consistency, efficiency, and architecture were strong, and fragmentation was minimal. This travel segment also exhibited relatively stable, though not always balanced HRV.

In summary, the travel sleep segment of this study revealed strong sleep resilience and adaptivity across diverse time zones. Sleep architecture, fragmentation, and consistency were generally strong, but these factors also highlighted the importance of maintaining sufficient sleep hygiene and opportunity. Sustaining a balanced HRV, despite red-eye travel, was potentially due to DELTA001’s systematic regimen of fitness, TRE, and other interventions. Nonetheless, travel sleep findings reinforce the important relationship between consistent and efficient sleep with autonomic function (HRV), heart health, human performance, and other factors. The overall findings of DELTA001 international travel suggest that strong sleep is possible across multi-time zone travel, and disruption was related primarily to travel modality instead of time zone change.

A comparison of sleep trajectory between retrospective (RETRO), start (START) and end (END) of the sleep study also revealed clear improvements (Fig. 6F). For total sleep, RETRO was 372.3 ± 12.3 minutes. START was 454.6±8.2 minutes, and END was 493.3±5.0 minutes. For REM sleep, RETRO was 65.2±2.7 minutes, START was 90.6±4.5 minutes, and END was 108.0±4.8 minutes. For light sleep, RETRO was 286.7±8.3 minutes, START was 324.4±6.4 minutes, and END was 341.6±5.7 minutes. For deep sleep, RETRO was 28.9±1.8 minutes, START was 43.0±2.5 minutes, and END was 43.3±2.9 minutes. For awake time, RETRO was 26.6±3.5 minutes, START was 19.2±3.7 minutes, and END was 13.2±1.4 minutes. Total sleep, deep sleep, REM sleep, and light sleep showed general trends of improvement. Awake time also showed substantial drops across devices. These observations imply that the DELTA sleep intervention as part of the general DELTA protocol was effective at improving sleep performance, with statistical significance observed across certain sleep stages.

While sleep architecture and duration served as the key metrics for this study, the average sleep duration in each stage was recorded across all three devices (WHOOP, Garmin, Apple) (Fig. 7 and Table S3). This study did not compare device performance. Sleep scores were also recorded on the WHOOP and Garmin devices (Fig. S16 and Supplementary Data 1). As a potential gamification opportunity, DELTA001 sought to achieve a score of at least 80 on at least one of the 2 devices during each sleep event (Fig. S16). A longitudinal record of these scores revealed a majority of sleep scores achieving this objective. While sleep scores may not always align with user perceptions of sleep outcomes, they may serve as an effective gamification tool, particularly if to enable acute and/or long-term and healthy behavior change.

**Fig. 7.**
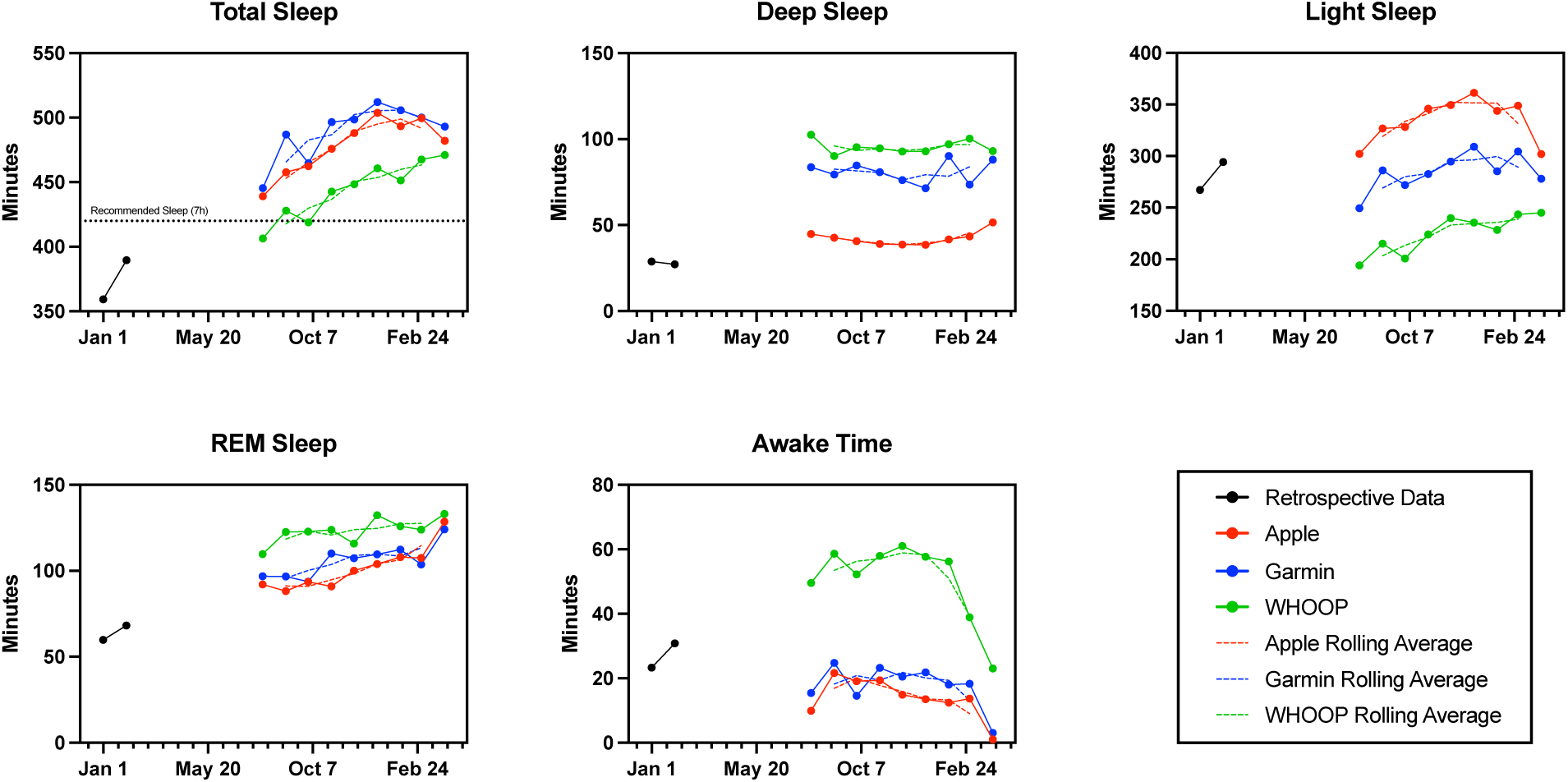
Sleep stages comparison across wearables. Sleep duration in each stage retrospectively and over the course of the study compared across Apple Watch, Garmin, and WHOOP devices. Source data can be found in Supplementary Data 1.

### Microbiome Dynamics

Microbiome composition and changes were studied as a function of a pre-48-hour fast (PRE48), post-48-hour fast (POST48), as well as 3-meals-a-day (3MAD). Dominant bacteria phyla included *Actinobacteria, Bacteroidetes, Firmicutes,* and *Proteobacteria* (Supplementary Data 2). The relative abundance of each phylum was plotted in accordance with varied testing conditions over time (Fig. 8A). It is interesting to note that for all samples across all timepoints, *Fusobacteria*, a prevalent microbial phylum often associated with poor diabetes control and serious gastrointestinal disorders, was not detected in any samples across all timepoints[102]. Kruskal-Wallis testing showed non-significant differences in the bacteria phyla relative abundances from each feeding condition (Fig. 8B). Shannon, Chao1, and Inverse Simpson diversity indexes and changes over time were also plotted, and Kruskal-Wallis testing also showed non-significant differences between microbe alpha diversity across the feeding conditions (Figs. 8C and S17). Pre- and post-fast (48 hours) showed non-significant relative changes in bacteria phyla across seven time points during a one-meal-a-day (OMAD) week (Fig. 8D). The *Firmicutes/Bacteroidetes* (F/B) ratio was also calculated, showing no significant difference between feeding conditions (Fig. 8E).

**Fig. 8.**
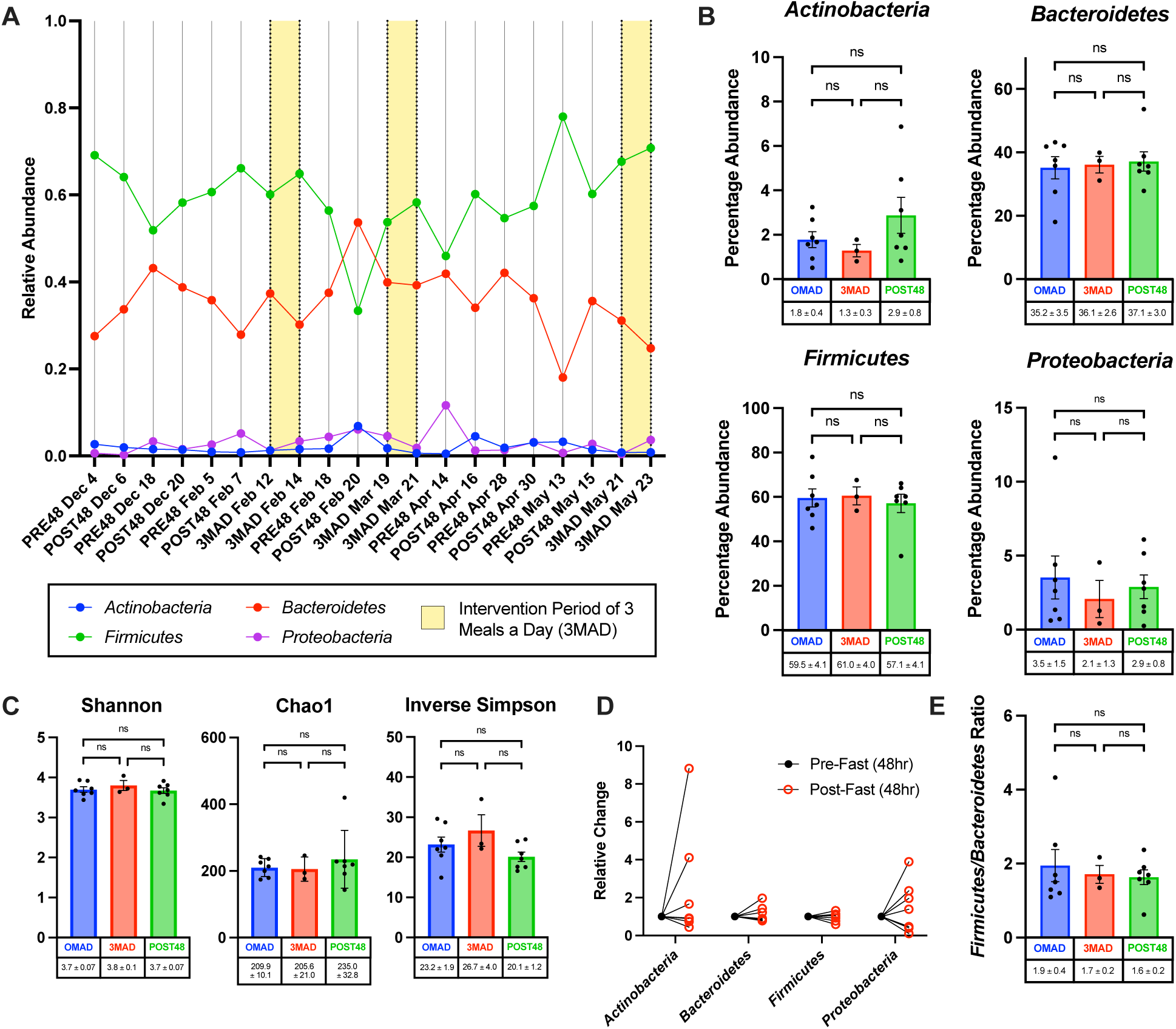
Microbiome compositions and diversity. **(A)** Relative abundance of bacterial phyla over the course of the study at PRE48 (pre-48-hour fast during OMAD week), POST48 (post-48-hour fast during OMAD week), and 3MAD (during three-meals-a-day week; highlighted with yellow background) timepoints. *Fusobacteria* were not detected in all samples across all timepoints. **(B)** Relative change in percentage abundance of bacteria groups – *Actinobacteria*, *Bacteroidetes*, *Firmicutes* and *Proteobacteria* – normalized to total amplicon sequence variant (ASV) values. A Kruskal-Wallis test was conducted, showing no statistically significant differences in bacteria group percentage abundances amongst feeding condition (p-value > 0.05). **(C)** Bar plots of Shannon, Chao1 and Inverse Simpson diversity indexes. A Kruskal-Wallis test was conducted, with each index showing no significant difference (p-value > 0.05) in diversity amongst feeding conditions. **(D)** Relative change in bacteria groups of *Actinobacteria*, *Bacteroidetes*, *Firmicutes* and *Proteobacteria* normalized to total ASV counts and then to pre-fast levels across seven time points during the OMAD week. **(E)** *Firmicutes*-to-*Bacteroidetes* (F/B) ratio based on percentage abundance normalized to total ASV count. A Kruskal-Wallis test was conducted, showing no significant difference (p-value > 0.05) in F/B ratios between feeding conditions. Data is represented by mean ± SEM. Source data can be found in Supplementary Data 2.

Functional pathway abundance predictions were also conducted (Fig. 9 and Supplementary Data 2). The paired line plots show the relative abundances of two PICRUSt2-predicted pathways (PWY-5532 and PWY490-3) in stool samples collected before (PRE48) and after (POST48) a 48-hour fast. A total of 327 pathways were analyzed, with the selected pathways displayed as they showed statistically significant differences based on a two-tailed paired t-test (p-value < 0.05). Each line (n=7) matches samples from the same participant. Predicted pathway abundances are expressed in arbitrary units derived from PICRUSt2. Some pathways that were important for microbial survival but displayed no significant change between fasting conditions include — GLYCOLYSIS, PWY-5505 and SER-GLYSYN-PWY.

**Fig. 9.**
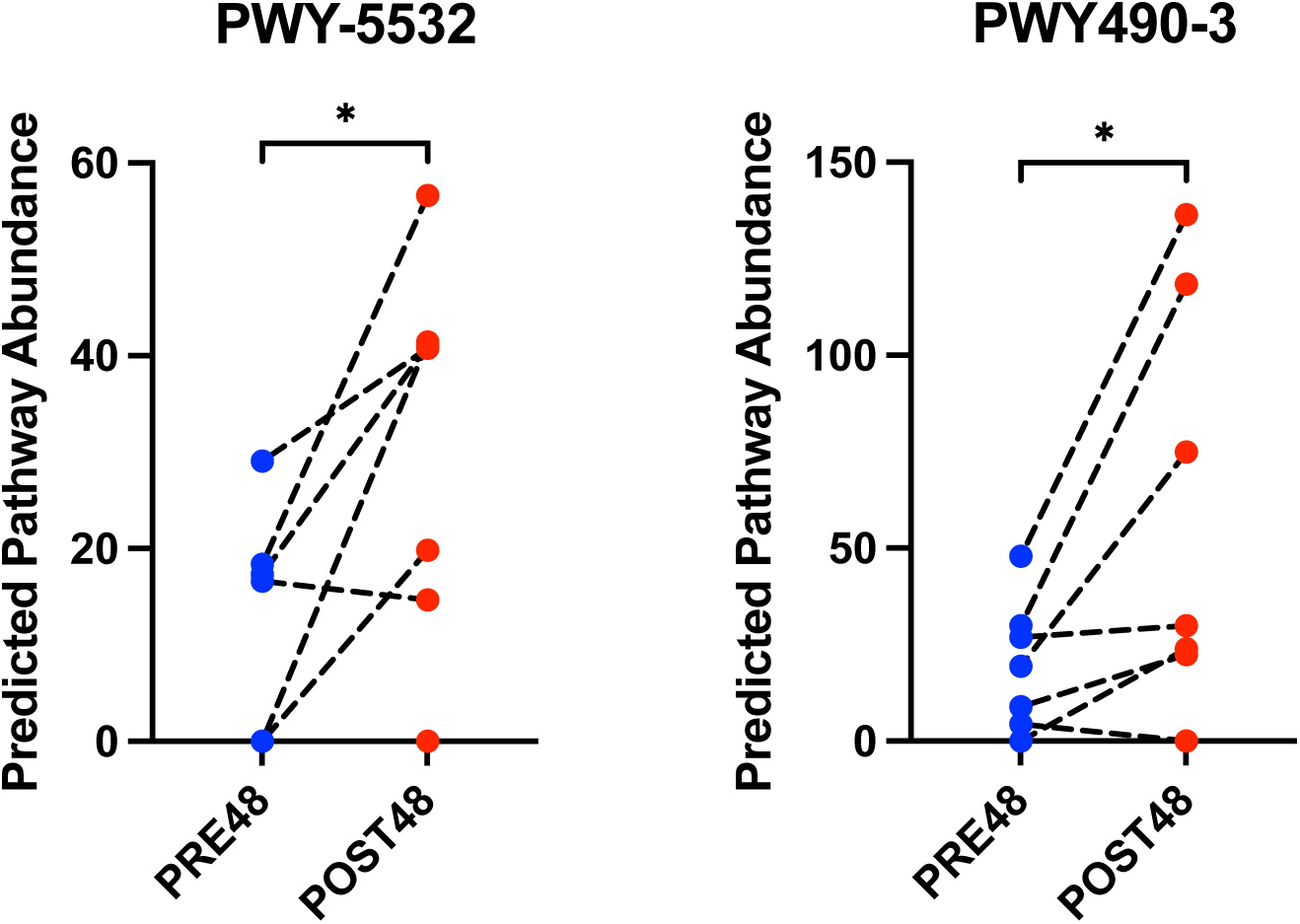
Microbiome functional pathway abundance predictions. Paired line plots show the relative abundances of two PICRUSt2-predicted pathways (PWY-5532 and PWY490-3) in stool samples collected before (PRE48) and after (POST48) a 48-hour fast. The selected pathways were displayed as they showed statistically significant differences based on a two-tailed paired t-test (p-value < 0.05). Each line (n=7) matches samples from the same participant. Predicted pathway abundances are expressed in arbitrary units derived from PICRUSt2. Source data can be found in Supplementary Data 2.

### Healthspan Copilot GPT

The “Healthspan Copilot”, a custom GPT designed to promote adherence and behavior change throughout the user’s health optimization journey, was tested through user-chatbot interactions to verify the correct execution of its configured logic (Fig. 10A and Supplementary File 2). The GPT supports biomarker benchmarking against the subject (DELTA001), provides recommendations for tracking delta values of biomarkers across different fasting states (denoted as ‘DELTA markers’ thereafter), and estimates biological age followed by an add-on analysis of the user’s biological resilience. The GPT was incorporated with DELTA001’s representative data on ketosis onset speed and DELTA markers that are potential indicators of biological resilience, including ΔApoB, ΔHomocysteine, and Δhs-CRP across different fasting states[103–105].

**Fig. 10.**
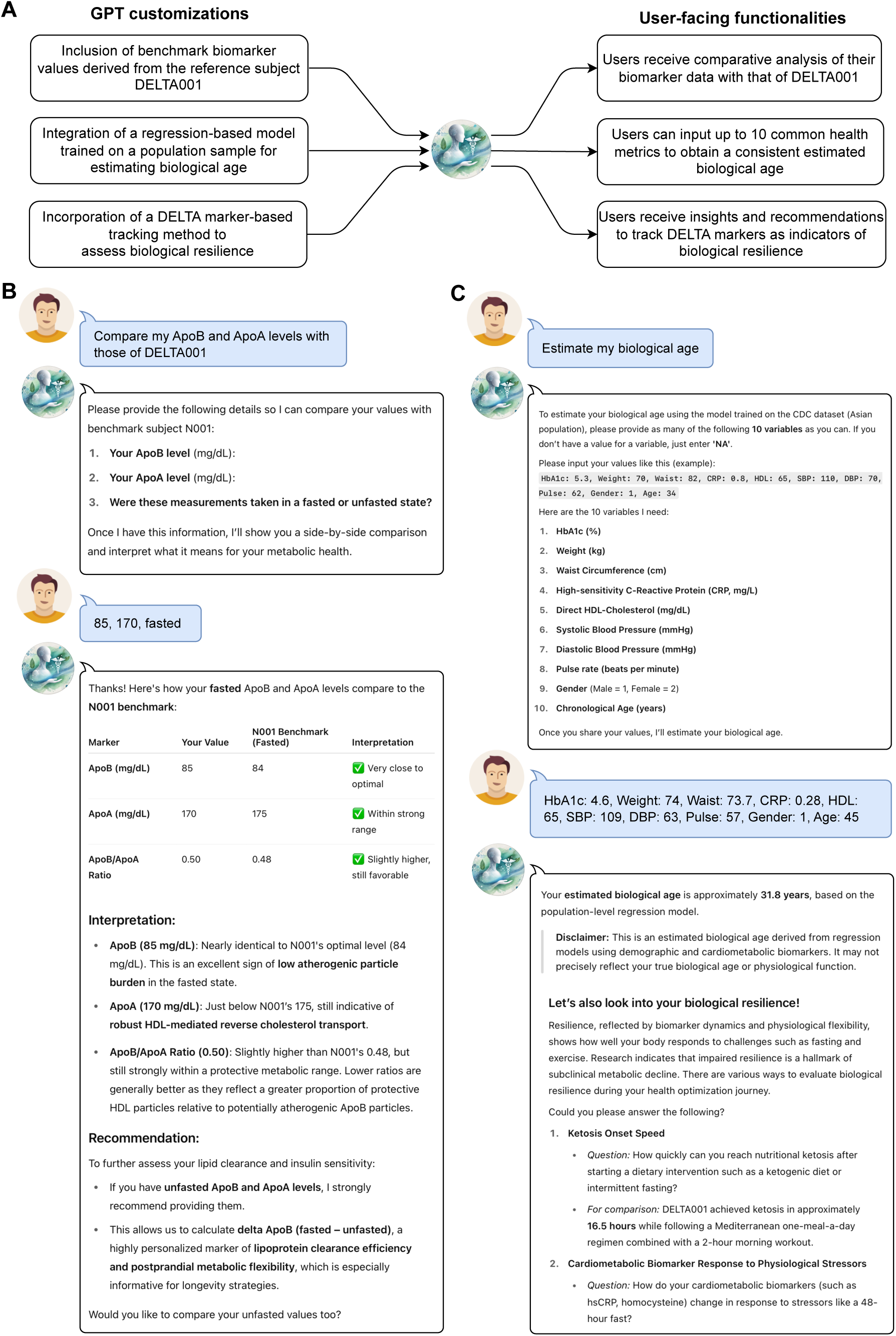
GPT-enabled dynamic health tracking. **(A)** Overview of the GPT-enabled platform for dynamic health tracking and behavioral adherence support, including system customizations and the corresponding user-facing functionalities. Example conversations with the custom GPT ‘Healthspan Copilot’ for **(B)** ApoB and ApoA analysis and **(C)** Biological age estimation with add-on module for resilience analysis. Source files can be found in Supplementary File 2.

During user-chatbot interactions, when the conversation starter “Compare my ApoB and ApoA levels with those of DELTA001” was selected, the GPT prompted for ApoB and ApoA values, identified the fasting state, and generated a structured comparison against DELTA001’s benchmark values with interpretive feedback on lipid clearance efficiency, as well as recommendation on the relevant DELTA marker to track (Fig. 10B). For the biological age estimation, the GPT consistently requested the required set of 10 health metrics upon user prompt and returned an estimate based on a population sample-trained regression model (Fig. 10C and Supplementary File 2). In the case of DELTA001, the estimated biological age using his health metrics from April 2025 was consistently 31.8 years, with potential further reductions due to his strong resilience profile. The estimate obtained from GPT matched the output of standard statistical software. Notably, the estimated biological age of DELTA001 is substantially lower than his chronological age of 45 years and corresponded with favorable biomarker data, including an HbA1c of 4.6%, body weight of 74 kg, waist circumference of 73.7 cm, hs-CRP of 0.28 mg/L, HDL of 65 mg/dL, systolic blood pressure of 109 mmHg, diastolic blood pressure of 63 mmHg, and a pulse rate of 57 bpm (Supplementary File 2). Although no quantitative performance evaluation was conducted, these test interactions showed that the GPT reliably executed its configured logic and produced consistent, context-aware outputs aligned with its intended purpose.

### Participant Experience: A Qualitative Assessment

Detailed and semi-structured interviews were conducted with DELTA001 at the study start (baseline), midpoint (after a 3MAD routine), and at the end of the study. Interview questions were not provided to DELTA001 in advance. The initial analysis outlined a baseline routine summary, detailing key pillars of exercise, food consumption, and sleep hygiene (Table 1). The DELTA001 participant experience was comprehensively detailed, outlining main study themes, sub-themes, and codes (Fig. 11). Four main themes emerged across the three interviews: “sustained engagement in routine and healthy lifestyle”, “engagement with digital health technologies”, “impact of changing mealtimes”, and “biomarkers.” Sub-themes included DELTA001’s motivation and challenges pertaining to adhering to the regimen, as well as additional concepts such as the use of technology to drive healthy changes, experiences while undertaking the fasting and fitness regimens, and the impact such data collection and interpretation had on the participant over the course of the study. Source files including transcripts can be found in Supplementary File 3.

**Fig. 11.**
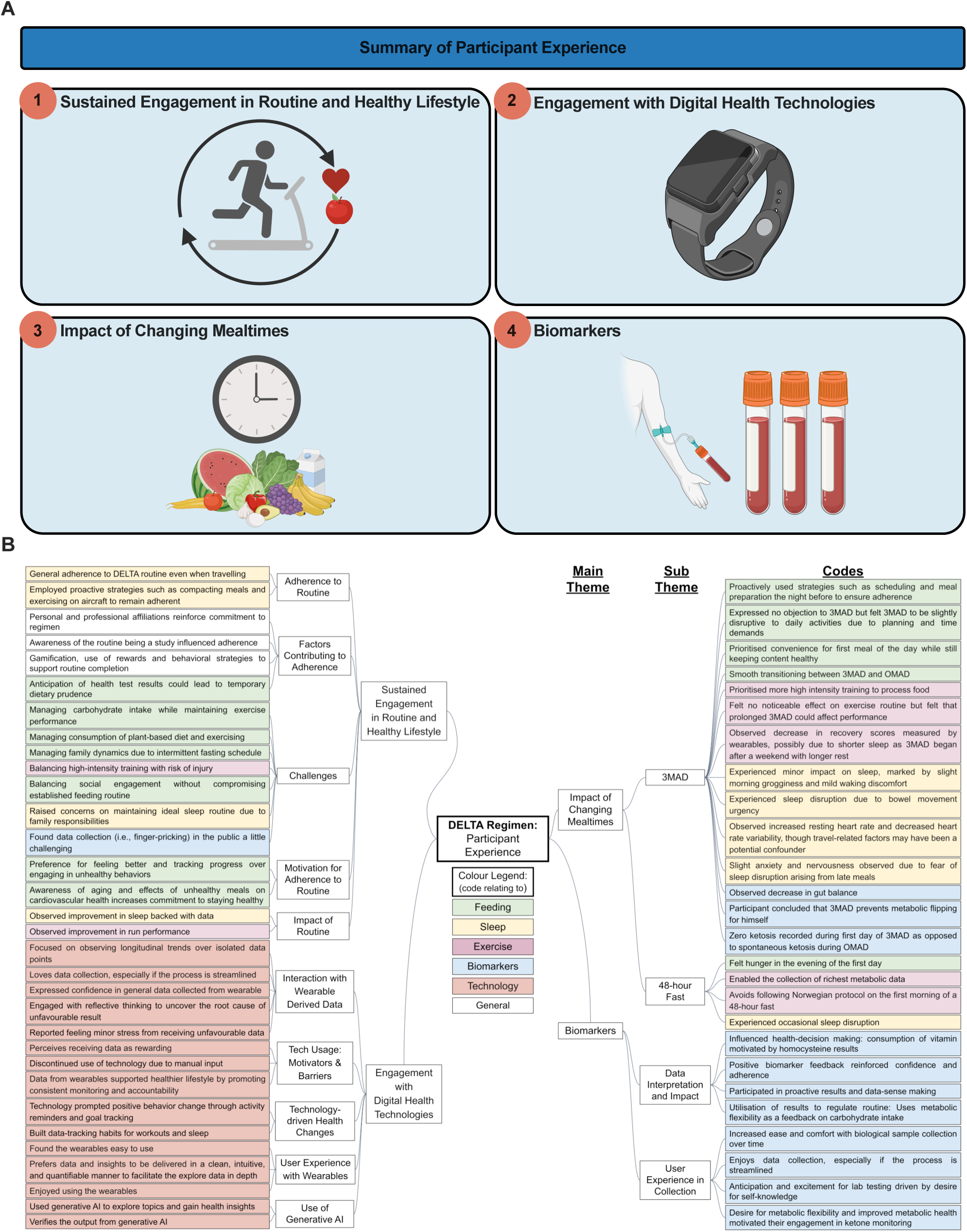
Coding tree of DELTA001’s participant experience of the DELTA regimen. **(A)** Summary of participant experience. **(B)** Codes are grouped according to main themes and subsequently further grouped into sub-themes. Transcripts from interviews can be found in Supplementary File 3. Figure 11A is created in BioRender (https://BioRender.com).

**Table 1.**
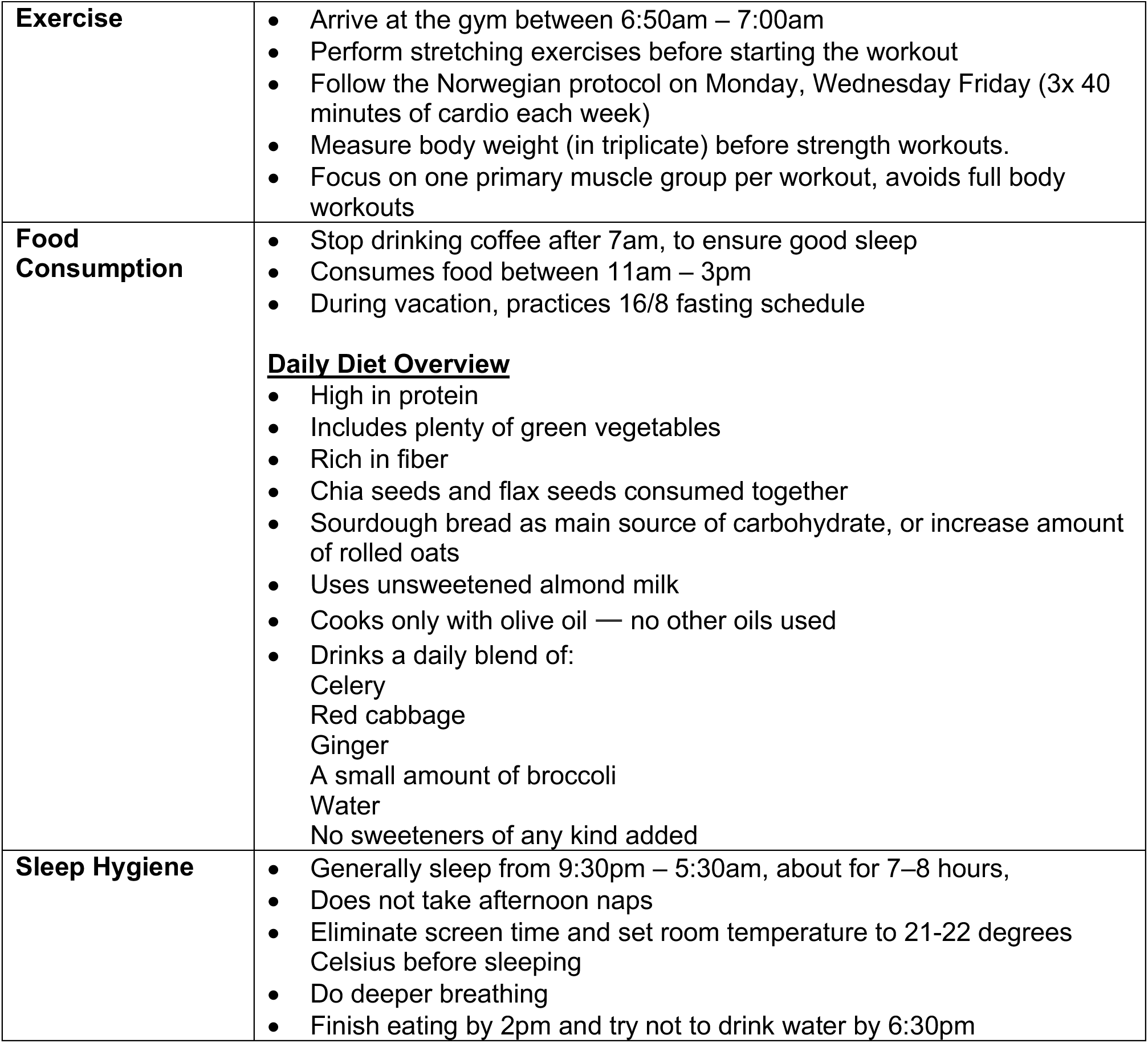
Summary of key characteristics of DELTA001’s experience from qualitative assessment of participant experience.

From the qualitative assessment of three interviews, DELTA001 described his experiences, motivations, and adaptive strategies. At baseline, DELTA001 reported extensive experience with TRE, OMAD, and healthy diet, aligning with the DELTA protocol, and also reported sustained adherence to the protocol with minimal difficulty in maintaining the lifestyle. Furthermore, increased awareness of aging and the impact of unhealthy diet on cardiovascular health contributed to a stronger commitment to maintaining a healthy lifestyle, thus deterring unhealthy behaviors.

> *“…but it’s like, as I get older too, I am very mindful of things like cardiovascular health and the impact of having, like one meal that’s not great, two meal that’s not great, three meal that’s not great, you don’t get kind of the flexibility to reverse that.”*

However, concerns were raised regarding the long-term sustainability of such lifestyle (e.g., navigating family dynamics and social engagements while practicing TRE, maintaining ideal sleep routine along with family responsibilities).

> *“Sometimes I had social engagements in the evening where I did not eat a single bite and by and large people joked about it with me. I was cool, I knew most of them. I don’t know if – that’s not going to be possible for every social event moving forward and I am ok with that.”*

In this study, adopting wearables led to multiple outcomes: (1) prompting positive behavior change through activity reminders and goal tracking, (2) receiving data (e.g., workouts and sleep) perceived to be rewarding, and (3) data tracking from wearables leading to consistent monitoring and reinforcing a sense of accountability.

> *“…it really has become a part of my life in terms of closing the rings (Apple Health feature) to make sure I burn minimum number of calories, stand a minimum number of minutes and get a specific amount of exercise in. I, I look to the data every day.”*

DELTA001 actively engaged in data sense-making when interpreting results and outcomes, which influenced decision-making. Particularly, time to reach ketosis was used as feedback on carbohydrate intake, while homocysteine dynamics motivated the targeted consumption of specific vitamins.

> *“I think having some insight into homocysteine status, for me, I something that I didn’t realize would come out of this was how it would prioritize my B vitamin consumption. I don’t I’ll never stop ensuring that I take enough vitamin B now.”*

During the 3MAD phase, changes in mealtimes impacted all components of the routine for DELTA001, resulting in slight anxiety and nervousness. For instance, he experienced minor impact on sleep, marked by slight morning grogginess and mild waking discomfort. Additionally, DELTA001 also made adjustments in his exercise routine, prioritizing more high intensity training to processed food.

## DISCUSSION

### DELTA Protocol

The DELTA protocol was developed by and solely for DELTA001 (author D.H.). The protocol included TRE (fasting), a daily fitness regimen, a daily dietary regimen, and a sleep protocol (9:15pm to bed, 6:00am wake-up, with variations for travel and work-related schedule changes). It should be noted that DELTA001 practiced the fasting and fitness regimen prior to the start of the DELTA study. However, the 4-hour eating window was shifted from a dinner-centric feeding window (5pm start)[82] to a mid-day eating window (11:00am – 3:00pm) in order to complete feeding with ample time for digestion prior to sleep. Also, the dietary regimen was largely practiced prior to the study, except with efforts to ensure that the polyphenol-based drink, chia pudding, and red cabbage salad were consumed as consistently as possible during DELTA. Changes to DELTA001’s sleep regimen, which included moving the sleep time to 9:15pm, were implemented as part of this study. Therefore, in the absence of a control subject or dataset, with the exception of the sleep performance assessment, no claims are made regarding the efficacy of the DELTA protocol in mediating substantial improvements to the health and performance profile of DELTA001. Nonetheless, these specific elements of the DELTA protocol were selected, as DELTA001 was capable of sustaining this protocol consistently.

Fasting was incorporated as a cornerstone of the DELTA protocol, as prior studies have shown that fasting, when done safely, may support insulin sensitivity[106]. Prior studies have also shown that fasting may reduce hepatic fat, improve fasting insulin and glucose levels, as well as reduce inflammation[107, 108]. Mitochondrial health and insulin signaling efficiency were also shown to improve under fasting regimens[49]. With regards to the feeding window and sleep, prior studies have shown that feeding windows that conclude with sufficient digestion time prior to sleep supported circadian insulin sensitivity resulting in favorable HOMA-IR and insulin levels even in the absence of weight loss[109]. Therefore, shifting the feeding window early was also intended to support the study sleep intervention.

The DELTA fitness regimen was comprised of daily strength training and HIIT 2-3 times per week[110]. The strength training focused on a primary muscle group per day. Prior studies have shown that high-frequency and high-volume training in older adults can enhance lean mass and strength[111, 112].

The DELTA dietary regimen was similar to a Mediterranean diet to support biomarker and functional health. A photo library of dietary items is provided (Figs. S2-S5 and Supplementary File 1). Extra virgin olive oil (EVOO) was the sole oil used for the entire study. Prior studies have shown that the Mediterranean diet has been effective in reducing major cardiovascular events atherosclerotic progression, reducing age-related brain atrophy, and improving biomarkers associated with cardiovascular disease risk[113–117].

### Cardiometabolic, Metabolic, and Pleiotropic Marker Flexibility

Participatory approaches to monitoring cardiometabolic, metabolic, and pleiotropic markers may be effective for some in the interventional optimization of health profiles. The development of novel biological resilience biomarkers based on the timescale of biomarker change and trajectories may also be possible. In this study, the rise of markers during fasting, followed by a return to baseline levels, is an indicator of cardiometabolic resilience. Methylation pathway resilience, which is an indicator of cardiometabolic through neurological health, is also reflected by the return to baseline levels.

The ApoB, ApoA, and ApoB/ApoA ratio dynamics observed in this study are indicative of metabolic agility and biological resilience. Specifically, the rise of ApoB following a prolonged 48-hour fast can be due to the efficient use of stored lipids as the body switches to fat-based energy. This can be reflected in an ApoB rise as it is present in all atherogenic particles. Importantly, this observation may reflect a functional response to energy demands during fasting, and appears to have a physiological and not pathological basis. The return to baseline following refeeding serves as a positive indicator of metabolic flexibility and efficiency of lipid metabolism[103–105]. This return could signify the efficient clearance of lipoproteins following physiological stress (fasting) and efficient insulin signaling, as ApoB catabolism is insulin-sensitive. Importantly, this return to baseline is also indicative that the POST48 rise of ApoB is not chronic, which may further reflect efficient functional metabolic flexibility[118–122].

The homocysteine and hs-CRP results observed for DELTA001 revealed an exceptional profile for cardiometabolic, inflammation, and potentially neurological health. The DELTA001 hs-CRP profile resided well beneath the low-risk threshold of 1.0 mg/L, and showed strong resilience to 48 hours of fasting. Prior studies have shown that in a healthy population (3672 male subjects) of similar average age and ethnicity, serum hs-CRP levels were closer to an average of 1.24 mg/L, with a positive correlation between hs-CRP levels and age[123]. Of note, prior studies have reported either no change or an increase in inflammatory biomarkers during prolonged fasting. In addition, CRP levels frequently rose, often significantly, during fasting periods, particularly in obese individuals. Importantly, several studies showed a reduction or normalization of CRP levels after refeeding, suggesting that the inflammatory response to fasting may be transient or adaptive[124–126].

The DELTA001 homocysteine readings largely resided within levels that have been classified as elite or optimal (Elite: 5-7umol/L; Low-risk: Below 10 µmol/L). Population studies have shown a median homocysteine level of 10.2 µmol/L in men aged 40-59[127], and the Framingham Study revealed that homocysteine levels below 6 µmol/L is rare amongst the participant population and a lower homocysteine level is associated with a lower risk of age-related conditions[91, 92]. DELTA001’s profile may have been a reflection of strong B-vitamin stores and efficient functional methylation. This was observed across the multiple cycles of homocysteine fasting and refeeding dynamics, revealing the eventual elimination of the substantial post-fast rise of homocysteine seen in the first cycle. Additional factors may include high metabolic flexibility, and the dietary and fitness components of the DELTA protocol playing a supportive role.

These findings support the use of controlled stressors, such as fasting, with biomarker monitoring to assess functional resilience. For markers like hs-CRP and homocysteine, a normalizing spike may indicate a favorable response, while no spike represents an optimal response. Fluctuations within a single cycle can be summarized by metrics such as AUC and tracked longitudinally until stable patterns emerge. This approach provides a potential basis for digital twin models or digital biomarkers of metabolic resilience, though further validation is required.

### General Health Outcomes

The DELTA001 health profile was generally favorable, with BP and pulse pressure residing within healthy ranges. DELTA001 weight management appeared to be consistent. While some positive health profile trends were observed, we cannot determine if they were attributable to the DELTA protocol, especially due to DELTA001 entering the study as a healthy subject. However, it is possible that the DELTA protocol may have played a supportive role in sustaining the health profile observed. As noted, prior studies have shown that consistent exercise, Mediterranean diet, and favorable sleep health have resulted in positive health outcomes[82, 128–135].

### Performance Outcomes

Grip strength as a function of blood pressure, and run performance are among the many metrics that can be used to assess human performance[101, 136–139]. One metric that is commonly used, but was not assessed in this study, was VO_2_ max (Maximal aerobic capacity). Favorable VO_2_ max profiles have been correlated with the reduction of all-cause mortality and are frequently cited as a major objective for healthy longevity[140, 141]. We did not use VO_2_ max for this study, including with the wearable platforms due to running sessions being conducted in the early morning and in consideration of DELTA001 safety and avoidance of training during darkness. However, VO_2_ max will serve as a primary objective of a subsequent participatory DELTA trial that is being designed. Nonetheless, run performance was assessed based on the documentation of run speeds maintained at 4-minute intervals, with corresponding heart rate as proxies. While VO_2_ max can be estimated from these metrics, we refrain from making these estimates as the subsequent trial will provide more definitive data. There are a number of ways to assess human performance outcomes as a function of health protocols. For this study, no claims are made as to whether or not the DELTA protocol was globally optimal for DELTA001. The primary outcome of the effectiveness of the protocol reported here is that it was sustainable for DELTA001.

### Sleep Performance

Making concerted efforts to address sleep consistency, sleep efficiency, and sleep architecture can significantly protect long-term brain, metabolic, and heart health. This may be particularly important for late sleepers. Importantly, prioritizing sleep as part of a health regimen, when possible, represents a low-risk, high-benefit intervention that is widely supported by population and mechanistic findings[5, 6, 9, 142].

We estimate that moving sleep time earlier may have been a key driver toward clear improvements in DELTA001 sleep performance. Additional supportive pillars of the DELTA sleep regimen may have included improved sleep hygiene, morning fitness, TRE regimen, and possible gamification driven by wearables use that could have influenced the behavior of DELTA001. The sleep study can be assessed by the following core domains: overall sleep architecture, changes in the sleep stages during the study, the role that adjacent interventions may have had on sleep improvement, and the role of wearables, among other factors.

Sleep architecture is comprised of the pattern and structure of sleep cycles, including N1-N3 and REM sleep. Previous studies have shown that sleep architecture, in particular slow-wave and REM sleep can be improved by consistent physical activity[143, 144]. Other studies have shown that sleeping earlier may have supported favorable circadian alignment, which is also known to regulate metabolism, hormone release, and inflammation. Conversely, circadian misalignment (e.g., late sleep) has been known to disrupt insulin sensitivity and glucose metabolism[145]. The DELTA001 cardiometabolic profile, metabolic switch efficiency, and resilience may further support the notion that moving sleep earlier was potentially supportive of these outcomes.

In this study, we estimate that moving sleep time earlier may have helped DELTA001 achieve consistent sleep patterns that is known to be protective against metabolic disease, cognitive decline, and improve cardiovascular health[146–148]. This is supported by a growing collection of evidence, especially in individuals with evening chronotypes. For example, prior studies have shown that slow-wave sleep (SWS) is critical for memory consolidation[149], among other factors.

For deeper assessment, the DELTA protocol, which included shifting sleep earlier (from midnight to ∼9:15pm), exercising daily in the morning, and finishing dinner by ∼3pm), may have supported factors such as circadian alignment, metabolic resilience and flexibility, as well as autonomic balance. These parameters were potentially reflected by the marker profiles observed, such as the HRV trajectory (Fig. 4A,B), as well as the sleep architecture and efficiency (Fig. S16).

More specifically, the adoption of earlier sleep and potential circadian alignment may have enabled the HRV trajectory observed, improvement in deep and REM sleep, and potentially the earlier onset and stability of sleep that was observed. Mechanistically, this has been supported by studies showing that circadian alignment may improve parasympathetic activity during SWS in the early stages of sleep, which can improve HRV and cardiometabolic markers. Adopting consistent and properly timed sleep chronotypes (e.g., Generally 8:00pm to 10:00pm, subject to individual considerations) alongside sufficient sleep duration of 7-8 hours have also been shown to reduce coronary heart disease risk[150]. Melatonin has also been shown to peak earlier with earlier sleep, which may improve sleep efficiency[151, 152].

Morning exercise, a consistent pillar of the DELTA protocol, has been shown to stabilize HRV, reduce sleep fragmentation, and potentially increase REM sleep, which were observed in the study. This may be explained by morning fitness regimens having been previously shown to improve sleep-wake cycling and evening parasympathetic tone, which can also improve HRV and sleep performance, even without delaying melatonin release[153]. Furthermore, studies have shown that evening fitness regimens may negatively impact HRV and REM sleep whereas morning workouts may support HRV and REM sleep improvement[154]. However, the DELTA study did not assess this comparison as workouts were solely done in the morning.

As previously noted, regarding sleep travel, substantial efforts were undertaken to adapt to the different time zones. Deferring late-night eating during evening flights prior to sleep, resuming morning fitness on arrival, maintaining a sensible, Mediterranean-inspired diet, and observing early chronotype sleep times likely played a key role in circadian alignment, strong sleep architectures, and balanced HRV during travel. In summary, maintaining a comprehensive approach to sleep health, including both domestic sleep and sleep during travel across diverse time zones, may potentially have collectively driven the sustained improvements in sleep performance, which was observed across all three wearables, reduced fragmentation, and favorable RHR, among other positive health characteristics.

### Microbiome Dynamics

Key findings from this study imply that for DELTA001, a 48-hour fasting event does not significantly alter bacteria group levels and consequently, the *Firmicutes/Bacteriodetes* ratio. Additionally, for DELTA001, 48-hour fasting did not significantly change the level of microbe diversity as observed across 3 different diversity indices — Shannon, Chao1, and Inverse Simpson. Furthermore, the notable absence of *Fusobacteria* observed at all collection timepoints might imply that the DELTA001 protocol diet, and possibly DELTA001’s fasting protocol could potentially play a role in maintaining low/no *Fusobacteria* levels — thus, potentially lowering pathogenic risk.

Regarding metabolic pathways that presented significance due to fasting conditions, PWY-5532 refers to a microbial salvage pathway associated with Archaea domain for recycling nucleosides and nitrogenous bases by degrading them into sugar phosphates. An upregulation of this pathway will result in an increase in 3-phospho-D-glycerate (G3P), which is a necessary intermediate for glycolysis. By extension, it is possible that the upregulation of this pathway contributes to carbon and energy recycling[155], by upregulating glycolysis and consequently also generating energy and metabolic intermediates for other pathways during fasting conditions when sugar (and thus carbon) sources are limited. Aside from being directly associated with gut archaea, such pathways may also be associated with horizontal gene transfer occurring between archaea and bacteria within the favorable environment of the gut[156]. PWY490-3 refers to an assimilatory nitrate reduction pathway, reducing nitrate to nitrite and ammonium. The latter will potentially contribute to nitrogen recycling via ammonia assimilation pathways and incorporation into amino acid production (e.g., glutamine and glutamate)[157], which act as core building blocks for microbial protein synthesis and growth[158]. Upregulation may also help manage redox stress by siphoning nitrate away from pro-oxidative pathways (e.g., nitrosamine formation — which can be carcinogenic). The DELTA fasting protocol may have contributed to limited carbon and nitrogen availability which potentially resulted in the upregulation of this assimilatory pathway in order to maintain microbiome survival and by extension, host-microbe homeostasis.

The upregulation of PWY-5532 and PWY490-3 likely contributed to the lack of significant changes in key metabolic pathways by serving as compensatory mechanisms that maintain essential metabolic functions under fasting conditions. PWY-5532 produces G3P, a necessary intermediate for glycolysis I from glucose-6-phosphate (GLYCOLYSIS), while PWY-490-3 generates ammonium, which is required for PWY-5505, the biosynthesis of L-glutamate and L-glutamine. These metabolites also feed into SER-GLYSYN-PWY (superpathway of L-serine and glycine biosynthesis I), which depends on both G3P and L-glutamate for the production of serine and glycine, key precursors for one-carbon metabolism and nucleotide biosynthesis. Together, the compensatory roles of PWY-5532 and PWY-490-3 suggest a degree of metabolic flexibility that supports energy production and sustains key biosynthetic processes during nutrient deprivation, such as a 48-hour fast.

### Qualitative Assessment of Participant Experience

The interview findings are limited by the N=1 design, which restricts the generalizability of said findings. Furthermore, DELTA001 represents a unique case, as the baseline level of health literacy, familiarity with dietary practices (e.g., OMAD and TRE), and engagement with self-tracking technologies are not typical of the general public. This pre-existing alignment with the DELTA routine may have contributed to smoother adoption and fewer challenges compared to what others may experience. Despite this limitation, the qualitative component significantly enhanced the study by providing rich, context-specific insights into the participant’s experiences, motivations, and adaptive strategies. The case study design allowed for the identification of nuanced barriers and facilitators – such as family dynamics, time management, and social engagements – that might challenge individuals attempting to follow a similar routine. These findings may thus serve as a guide for designing future protocols tailored to individuals at different readiness levels.

### The Paradigmatic Shift Toward Longitudinal Biomarkers

The findings of this study underscore the critical importance of longitudinal data in providing temporal context and revealing patterns of the authentic biological narrative of disease progression, advancing toward more preventive and personalized healthcare[159–161]. Digital biomarkers offer crucial “protection against health-to-disease transitions”[162].

Unlike single-point measurements that capture static physiological snapshots, longitudinal biomarker monitoring enables continuous biological surveillance that captures temporal patterns over extended periods, such as the comparison between blood glucose measurements versus HbA1c in diabetes care. Evidence supports longitudinal biomarkers applications across diverse therapeutic areas, including asthma, sleep disorders, diabetes, cardiovascular diseases, and chronic obstructive pulmonary disease (COPD) management[162, 163]. However, longitudinal biomarkers in aging, longevity, and preventive medicine represent an emerging paradigm with substantial untapped potential[28].

Consumer wearables increasingly provide clinically validated measurements, thereby establishing novel digital biomarkers. HRV demonstrates particular promise for cardiovascular risk stratification and early detection. Large-scale validation studies, including the U.S. National Health and Nutrition Examination Survey population cohorts, have established wearable-derived biomarkers for predicting inflammation, biological aging, and mortality outcomes[164], thus expanding beyond traditional laboratory-based measurements. The integration of traditional biomarker data with wearable-derived metrics enables hyper-personalized healthcare through real-time physiological monitoring. Automated insulin delivery systems exemplify this integration, combining continuous glucose monitoring (CGM) with insulin pump technology for closed-loop glycemic control.

Optimal strategies to fortify healthspan are unlikely to be fixed or static, as aging is a dynamic and adaptive process influenced by internal and external stressors over time. Although biological age is a holistic measure of functional health that can encourage adherence to interventions, most current approaches rely on static snapshot-based assessments[165]. Given that biological aging reflects the cumulative and ongoing adaptations to stressors, incorporating longitudinal assessments – particularly those capturing resilience and dynamic responses – could provide a more comprehensive perspective on optimizing healthspan[10, 11]. Such an approach could also combine factors including but not limited to longitudinally monitored biomarkers, their respective response time constants and recovery kinetics, performance assessments, and measures of biological resilience. While current biological age clocks are becoming more established, the incorporation of resilience metrics and longitudinal monitoring remains relatively underdeveloped, presenting an opportunity to explore a hybrid framework that leverages both[12, 166].

Past studies have demonstrated the potential of AI chatbot-based tools in promoting long-term behavior change and health engagement[167, 168]. With appropriate design grounded in behavioral science and evidence-based biomedical data, GPT-based system could potentially support longitudinal health monitoring effectively, with and without resilience-focused monitoring, to bolster biological age assessment and potentially gamify adherence to continuous monitoring[169, 170]. Moreover, the data from this monitoring may itself even serve as an intervention to fortify healthspan, serving simultaneously as feedback and a motivational incentive.

### Beyond N=1 Data

While DELTA was an N=1 study, learnings may be applicable toward population-level studies. Firstly, the DELTA protocol was a combinatorial regimen, which may have played a role in driving the positive sleep and biomarker outcomes observed. Systematically designing combination regimens, including regimens comprised of micro-adjustments, for subsets of participants may be effective and warrants further study. Emerging studies are starting to look at health impact due to potential synergies[171, 172].

Beyond combination approaches, even single interventions, such as micro-adjustments in advancing sleep times, particularly for late chronotype sleepers, could drive substantial health improvements when sustained even for one week. Late chronotype sleepers (e.g., midnight) can consider small, systematically implemented steps to advance sleep times earlier (e.g., by 15 to 30-minute increments). Consistently sleeping earlier (e.g., 10:00pm-11:00pm) has been shown to boost SWS, which may improve glymphatic elimination of β-amyloid and protect against cognitive decline by reducing sleep fragmentation[173]. Importantly, sleeping earlier may also reduce RHR and BP variability, as well as lower stroke and coronary artery disease (CAD) and stroke risk[174]. Wearables may help with gamification of adherence to gradually moving up sleep time due to their ability to rapidly provide feedback and nightly data[175]. Micro-adjustments may also be more sustainable compared to substantial shifts. A prior study showed that subtle sleep times advances of 15-minutes every few days resulted in improved sleep quality, mood, and energy within 1-2 weeks[176]. Improved circadian alignment and melatonin secretion to address jet-lag and late shift work, was also shown with micro-adjustments of 15-20 minutes when combined with consistent wake times, avoiding screen time and late caffeine consumption[177]. 30-minute shifts have improved sleep efficiency[178]. Importantly, a 1-hour shift can markedly reduce depression risk[179]. Larger shifts of 2 hours for very late sleepers (2:30am) can result in measurably improved health that included reduced depression scores and cognitive performance improvement. Of note, results appear as quickly as 1 week into intervention[180].

With regards to biomarker readings, this and other studies reveal potential opportunities to explore population-scale trials to better determine if earlier monitoring (e.g., young and middle-aged adult screening) can enhance the impact of preventive care toward healthy aging in later years. For example, homocysteine screening in young adults is not prevalent and has largely focused on the senior community[181–184]. However, the incidence of metabolic disease is increasing in young adults. This can also eventually lead to neurological and neurodegenerative disorders. Expanding homocysteine screening in young adults and middle-aged adults may lead to improved prevention of cognitive decline at a population level. Expanding the range of markers taken as a part of standard care, following adequate clinical validation, may eventually be justified. Beyond expanding the range of single biomarkers that may be screened, developing novel biomarkers based on resilience dynamics may also offer insights into general health and biological aging. While these analyses would require more blood draws and other logistical considerations, the insights realized from these biomarkers may find interest beyond N=1 studies.

The DELTA trial also offers insights into the need for expanded sets of biomarkers available for accessible laboratory, wearable, and home-based monitoring. In addition to point-of-care biomarker testing, it will be increasingly important to develop predictive approaches that use digital technologies to quantify biomarkers derived from biological samples, and vice versa. Furthermore, due to some correlations that exist between biomarkers, more studies are needed to harness these relationships so that levels of one biomarker can potentially be used to predict levels of another biomarker. Realizing a markedly expanded repertoire of biomarkers may play a major role in democratizing preventive health.

### Economic Implications

Emerging evidence demonstrates the economic value of longitudinal biomarker monitoring. CGM has proven cost-effective, reducing hospitalizations while improving quality of life in diabetes patients[185–187]. Additionally, home-based blood pressure monitoring proves more cost-effective than in-clinic monitoring[188, 189],while early screening and diagnostic interventions demonstrate significant economic benefits consistent across multiple scenarios, including strata of sex and age[190, 191].

Furthermore, the transition toward longitudinal monitoring creates economic value through multiple complementary pathways beyond traditional healthcare cost savings. Longitudinal data streams generate proprietary datasets that could be monetized through pharmaceutical partnerships, providing unprecedented granularity for drug development, clinical trial optimization, and real-world evidence generation[192]. Healthcare systems can leverage these datasets for risk stratification algorithms, population health analytics, and predictive modeling services that command premium pricing in the emerging health data economy.

Longitudinal biomarkers facilitate the transition from fee-for-service to value-based care paradigms, including subscription-based preventive care models, realigning provider incentives with health maintenance rather than episodic illness treatment[193]. Contemporary healthcare reforms, including the UK’s NHS prevention-focused initiatives and Singapore’s Healthier SG program, demonstrate how continuous monitoring data enables outcome-based payment structures that drive sustainable, prevention-oriented healthcare delivery.

Furthermore, insurance companies increasingly recognize the actuarial value of longitudinal biomarker data, offering premium discounts and coverage expansions for continuous monitoring participants, creating direct consumer benefits while reducing long-term claims exposure.

### Participatory Health Considerations

In the DELTA trial, biomarker data and performance outcomes were readily visible to DELTA001. This raises an important consideration for this and future trials for both DELTA001 and population studies. In particular, a potential tradeoff between conventional trial designs accounting for scientific control, and a participatory approach. In conventional trial designs, participants are blinded from their data in order to maintain objectivity, potentially influencing their behavior. This may confound results, making it difficult to determine the efficacy of an intervention. However, this design may also end up with a result that is already known, without offering a solution, the efficacy of which may in fact be the ability to influence the participant’s behavior.

This study used a participatory health approach, where the real-time data is visible to the participant, and the data itself may serve as an intervention[82, 194]. Participatory health approaches have been previously explored to empower participants to play a first-hand role, driven by agency, in co-designing their own health improvement through behavior change[195–197]. Current qualified self-studies are also harnessing this approach, whereby digital health devices are catalyzing health improvements outside of conventional trial designs[82, 198].

This study opens doors to potential downstream participatory health studies that are both interventional and preventive. Current large-scale biomarker studies have focused on middle-aged adults or seniors, when the biomarkers generally trend higher due to their positive correlation with age. This would render interventional studies that are reactive, versus preventive. It is possible that expanding these studies toward young adults, where participatory health studies may be more well-received, will also support larger effect sizes if the participatory approaches result in behavior changes that occur earlier at an earlier life stage.

Substantially more validation is needed, but this work illuminates possibilities to potentially harness data as the intervention to drive sustainable disease prevention, at least for a segment of the broader population.

### Study Limitations

There are a number of limitations to this study. Firstly, this study was conducted on a single subject. Therefore, population-level conclusions should not be derived from this work. With regards to the DELTA protocol, the primary intervention of 48-hour fasting, daily eating window, as well as fitness, supplementation, and dietary regimen were designed for DELTA001, and is not meant for widespread adoption due to fundamental differences between individuals (e.g., age, biological sex, race, baseline health, etc.).

As discussed, data was visible to DELTA001 throughout the study. It is highly probable that the study data directly influenced DELTA001 behavior. As a participatory study, DELTA differs from conventional trial designs. Objectivity was not a component of this study by design. As the field of participatory health continues to expand, it is critically important to draw clearer and defensible links between interventions and outcomes, and to develop solutions that can be sustained beyond initial participatory behavior change. Furthermore, sustaining a feeding regimen that concludes by approximately 3:00pm raises the issue of sustainability due to the importance of maintaining social bonds and connections that are evidence-backed drivers of healthy longevity. While early completion of feeding may support positive sleep outcomes, it is important to carefully weigh the importance of maintaining a balance of key pillars that promote positive wellbeing.

We cannot conclude if the DELTA protocol and or its components are responsible for the biomarker outcomes, and if so, which components were primarily responsible. Variability and confounding factors are also to be considered. While effort was made to keep meal composition consistent, variability was implicit. The biomarkers selected for this study are not meant to be exhaustive, nor do we imply that they represent actionable routes toward maximizing healthspan. It is also important to note that biomarker classification (e.g., elite, optimal, normal etc.) may not be internationally standardized, and further trials and consensus are needed in order for the adoption of non-standard biomarkers into practice[97, 199, 200]. More work is needed to determine their population-scale impact, if any, toward driving improved human longevity outcomes. With regards to fasting, it is not recommended for everyone, and some subjects should not fast. While fasting has been shown in previous studies to benefit the metabolic health of suitable subjects, outside of being sustainable for DELTA001, we cannot conclude that the DELTA fasting regimens are suitable for other participants. We also do not conclude that the fitness duration and regimen employed are suitable for all participants. It is important to note that inter- and intra-individual differences make personalization an important aspect of regimen design, and the same individual is likely to require dynamic changes to their regimen to sustain optimal outcomes.

This study used grip strength and run speed as additional proxy markers for performance outcomes. These markers provide some insight into performance trajectory over time while on the DELTA protocol, but they are not exhaustive. A number of additional markers could have been included, and will be explored in subsequent DELTA trials for DELTA001.

With regards to the microbiome dynamics study, we are unable to determine if the daily DELTA fasting protocol, which is DELTA001’s baseline regimen, resulted in a major gut microbiome composition change compared to a sustained non-fasting protocol. This study did not determine if DELTA001 gut health was optimal or suboptimal. It should be noted that some gut microbes that are associated with metabolism, heart health, and others were found, at some points, to be relatively low. However, since DELTA also provided functional readouts of cardiometabolic health, future studies may shed light on potential compensatory mechanisms that can sustain favorable phenotypic health outcomes. While DELTA001 did undertake 3 separate weeks of a 3MAD regimen, we do not know if the gut microbiome compositions during these weeks are truly representative of a sustained 3MAD regimen. While we do not observe major changes in gut microbe composition between the PRE48 and POST48 conditions, which imply that the fasting regimen does not have apparent negative impacts on DELTA001 gut health, in the absence of a true baseline, it is difficult to conclude if a continuous 3MAD regimen would result in markedly different microbe diversity. Furthermore, the upregulation of a metabolic pathway associated with the Archaea domain (PWY-5532) highlights the need to investigate not only the gut bacteriome but also the lesser-studied – yet equally important – gut archaeome[201], which was not specifically examined in this study. This is particularly relevant given emerging evidence linking the archaeome to various health and disease states, including diabetes, cardiovascular conditions, and even immune system regulation[202–204]. Nonetheless, the persistent and longitudinally validated absence of *Fusobacteria* serves as a positive indicator of the tolerance of the DELTA regimen.

The substantial longitudinal biomarker testing in this study was enabled by external funding as disclosed. Bringing longitudinal biomarker monitoring into standard testing protocols, even with sufficient evidence and validation, will require profound financial resources. Advancing longitudinal blood testing into real-world applications will require substantial further investigation that not only confirms what clinical actionability, if any, can be translated toward improved health outcomes and prevention of disease. Bringing these studies to fruition will also require health economics, implementation, and other factors to be addressed.

The GPT-based Healthspan Copilot was retrospectively applied to DELTA001’s data. As such, it has not yet been prospectively validated. In addition, the biomarker data used by the GPT for biological age estimation relied on conventional markers based on the available biomarker data in the population sample dataset. While the emerging set of biological resilience markers selected for the DELTA study (e.g., homocysteine, metabolic switch dynamics, etc.) were used to add additional support for potential further reductions in biological age, additional evidence will be needed to definitively support the use of these newer markers toward biological age estimation. Biological age is also not a universally applicable incentive to drive longitudinal biomarker collection or adherence toward healthspan-fortifying regimens. Additional incentives or approaches will likely be needed for future Copilot iterations.

When elective health studies or biomarker testing are conducted, caution should be taken. Risks associated with longitudinal blood draws include overdiagnosis, and special attention should be paid to ensuring that blood draw volumes remain within safe limits (10.5 mL/kg or 550 mL over an 8-week period, whichever is lower)[205].

## CONCLUSION

The DELTA trial served as an interventional, open-label prospective, and participatory approach toward assessing human biological resilience. A combinatorial regimen of fasting, fitness, Mediterranean-inspired diet, and substantially advancing sleep timing was combined with wearable, point-of-care, and clinical biomarker monitoring to longitudinally assess DELTA001’s cardiometabolic and pleiotropic biomarker resilience in response to stress intervention (e.g., fasting). Sleep and performance were also comprehensively assessed. A GPT-based framework was developed to address the potential of hybridizing emerging biological aging approaches with dynamic human resilience and performance testing to strengthen the actionability of these approaches. This was viewed specifically through potential gamification of adherence to longitudinal testing, with the data itself potentially serving as an intervention to encourage healthspan fortification. DELTA001 user experience was also assessed via a qualitative study. Biomarker resilience served as a cornerstone component of the DELTA study, and may open doors toward novel biomarker development based on individualized dynamics and recovery properties that can potentially be used for healthspan monitoring. In the DELTA study, DELTA001 cardiometabolic markers revealed highly agile metabolic flexibility in the form of ApoB, ApoA, and ApoB/ApoA ratio dynamics. Homocysteine and hs-CRP pleiotropic marker dynamics revealed resilience to metabolic stress (fasting) as shown by their optimal/elite levels. Clear sleep improvements were observed across the full architecture of sleep stages, and microbiome analysis revealed the sustained absence of fusobacteria. These findings imply that health biomarkers and regimens for the optimization of healthspan are unlikely to be a static process. Instead, the dynamic modulation of interventions may be essential for both biological optimization and potentially behavioral adherence. This study has also highlighted the need for solutions that can support quantified-self frameworks to promote longitudinal and expanded monitoring of healthspan-relevant biomarkers. These can range from improved infrastructure for decentralized collection of biological fluids for biomarker analysis, to wearables that either directly assess biomarker dynamics, or can also be used for predictive analytics of expanded biomarkers. In sum, the DELTA trial has illuminated the potential of leveraging participatory approaches to drive and sustain behaviors that can fortify the healthspan.

## METHODS

### Recruitment

This study is approved by the Institutional Review Board (IRB) of the National University of Singapore (NUS) under IRB-2023-801 and IRB-2024-397. The study was registered on ClinicalTrials.gov (NCT06630637) under the title “Monitoring Intermittent Fasting for Human Optimization Using Wearable Technology: An N-of-1 Study.” Recruitment period was from October 7, 2024 to July 2, 2025. Written informed consent was obtained from the participant (DELTA001; Author D.H.). Retrospective sleep data of the participant from first quarter of 2024 (IRB-2023-801) was accessed on April 3, 2025, and the study team had access to information that could identify the participant. DELTA001 (Author D.H.) is a healthy subject (Male, 45 years old at the time of recruitment and 46 years old at reporting) cleared by licensed medical professionals for the study, and had an estimated HbA1c of 4.6% (Abbott Freestyle Libre) (Fig. S1), no history of chronic/metabolic disease, and no prescription medication. No alcohol was consumed during the entire duration of the study. During the study period, the subject was clinically healthy with no reported or documented history of acute or chronic illness. Importantly, biomarker resilience studies and baseline biomarker readings were conducted in triplicate at a minimum, an uncommon feature in RCTs, but important for the evaluation of durable responses seen within an individual. This also serves to strengthen the design of larger studies based on N=1 findings.

### Adherence to Reporting Guidelines

In alignment with standard reporting practices, the SPIRIT extension for the N-of-1 trials (SPENT) guidelines were used to inform study design[206]. The CONSORT extension for reporting N-of-1 trials (CENT) guidelines were used to guide the writing of the manuscript[207, 208]. The SPIRIT-SPENT and CONSORT-CENT checklists and the CENT participant flow diagram can be found in the Supplementary Information (Tables S4-S5 and Fig. S18).

### Wearable Selection

DELTA001 used a WHOOP (4.0) (WHOOP Inc.), Garmin Epix Pro (Gen 2) (Garmin Ltd.), and Apple Watch 9 (Apple Inc.) for this study. This study did not assess device accuracy or compare device performance. Wearables were worn as frequently as possible during awake and sleep time with exceptions due to battery charging.

### Biomarker Measurement

Blood ketones and glucose were primarily measured by finger stick (Abbott Optium Neo). HbA1c estimates were provided by CGM (Abbott Freestyle Libre). Clinical biomarkers were taken by a Ministry of Health-certified medical clinic via orders placed by a licensed medical professional following assessment for participant health. Every effort was made to take blood at approximately the same timeframe (between 8:30am – 9:00am). For rapid testing of glucose and ketone dynamics, repeat finger sticks were not necessary between each reading. Clinical biomarkers were evaluated by a Ministry of Health-certified commercial laboratory. AUC of homocysteine was computed using the trapezoidal rule between successive timepoints (PRE48, POST48, POST-RF) and expressed in *μ*mol·day/L. The resilience index (RI) was defined using the equation below:

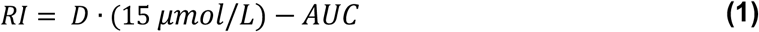

Where:

*D* is the total number of days during a fast and the refeeding period.

15 *μ*mol/L is the upper limit of a normal homocysteine level.

RI is expressed in *μ*mol·day/L.

### Metabolic Switching: Glucose-Ketone Dynamics

Glucose and ketone dynamics were longitudinally monitored following two distinct regimens: the 48-hour fasting strength training regimen and the OMAD ketosis entry regimen. The 48-hour fasting strength training sessions were conducted on the second day of a 48-hour fast (on Sep 20, Oct 10, and Oct 25 2024), each involving a weightlifting session lasting 45, 70, and 80 minutes, respectively. This was followed by a 3- to 4-hour recovery period during which blood ketone levels plateaued. Glucose and ketone measurements began in the early morning, with 2–4 baseline samples collected prior to exercise. Subsequent measurements were taken approximately every 5 to 20 minutes throughout the exercise and recovery phases. The OMAD ketosis entry regimen was conducted over two consecutive days for 6 sessions. On Day 1, the first measurement was taken immediately after consuming the day’s single meal, followed by 3–6 additional measurements until bedtime. On Day 2, the subject resumed glucose and ketone monitoring while performing a structured workout session lasting approximately two hours, consisting of yoga, weightlifting, and intermittent sessions of walking and running. This was followed by a recovery period lasting 2.5 to 5 hours to ensure ketosis state is entered. Time to ketosis is defined as the duration from post-meal measurement on day 1 to post-workout ketosis entry (blood ketone level reaches ≥ 0.5 mmol/L) on day 2. The glucose and ketone readings for both regimens were consistently photographically recorded for documentation purposes (Figs. S6–S13). For the 48-hour fasting strength training regimen, three phase-specific metabolic switching rates were computed to characterize substrate dynamics in response to exercise and post-exercise recovery: The Glycolytic Onset Rate, the Ketotic Recovery Rate, and the Total Substrate Switch Rate.

The Glycolytic Onset Rate was defined as the combined rate of glucose increase and ketone decline from the start of exercise to the time of GKI peak. GKI was calculated as follows:

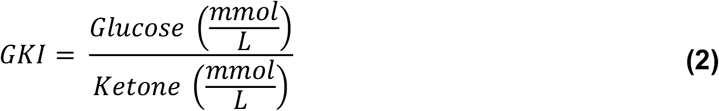

Glycolytic Onset Rate was calculated as follows:

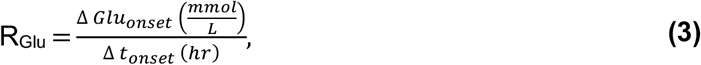

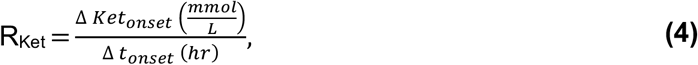

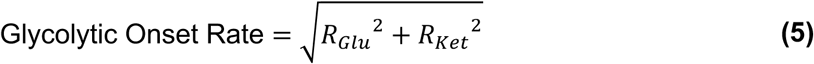

Where:

Δ *Glu_onset_* is the change in glucose level from pre-exercise baseline to GKI peak.

Δ *Ket_onset_* is the change in ketone level from pre-exercise baseline to GKI peak.

Δ *t_onset_* is the elapsed time from exercise start to GKI peak.

Ketotic Recovery Rate was defined as the combined rate of glucose decline and ketone increase from GKI peak to post-exercise ketotic recovery. Ketotic recovery is defined as the moment when ketone level stops increasing post-exercise. It was calculated as follows:

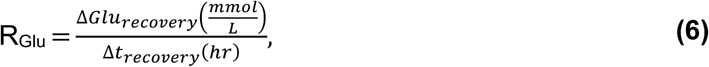

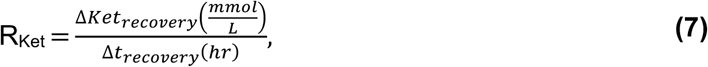

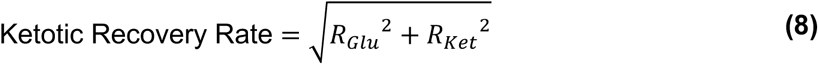

Where:

Δ *Glu_recovery_* is the change in glucose level from GKI peak to post-exercise ketotic recovery.

Δ *Ket_recovery_* is the change in ketone level from GKI peak to post-exercise ketotic recovery.

Δ *t_recovery_* is the elapsed time from GKI peak to post-exercise ketotic recovery.

The Total Substrate Switch Rate was defined as the combined rate of glucose change and ketone change during the glycolytic onset and ketotic recovery phases. It was calculated as follows:

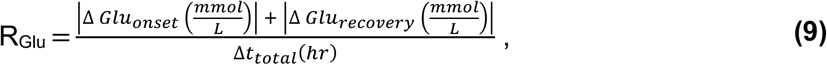

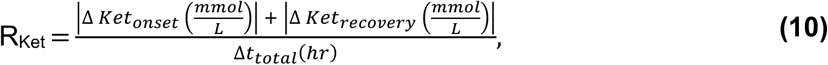

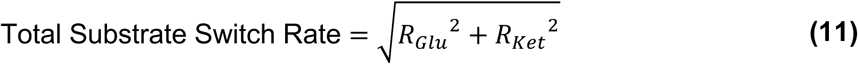

Where:

Δ *t_total_* is the total elapsed time from exercise start to post-exercise ketotic recovery.

### Health Profile Measurement

Blood pressure measurements were conducted (n=3 per session) in the morning, typically between 6:00am to 9:00am via arm cuff (Omron Intellisense BP7350). Body weight measurements were taken (n=3 per session) without shoes, typically between 6:00am-9:00am (KaradaScan).

### Heart Rate Variability Baseline Determination

HRV was assessed using the natural logarithm of the root mean square of successive differences (LnRMSSD) between heartbeats. To determine HRV baseline range, overnight HRV values were transformed using the natural logarithm: *LnRMSSD_t_* = ln *HRV_t_*

A 21-day rolling average (*μ_t_*) and standard deviation (*σ_t_*) were then calculated using:

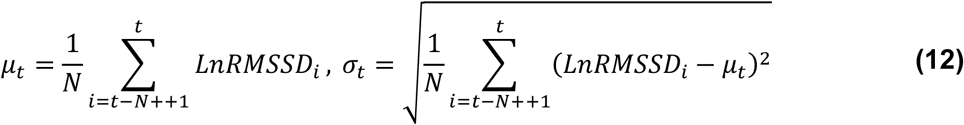

where *N* = 21 and a minimum of 14 days of data was required. The individual’s expected HRV range was then estimated by:

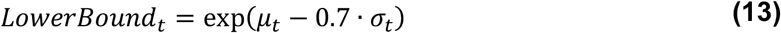

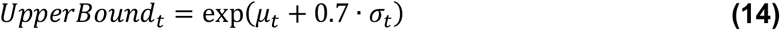

These bounds represent the typical HRV baseline range for the individual based on recent trends.

### Performance Analysis

Grip strength measurements were primarily taken (n=3 per session) for both the dominant (R) and non-dominant (L) hands (Camry EH101), typically between 6:00am to 9:00am. Toward the end of the study, dominant grip strength measurements were also taken due to the device’s improved resistance to grip slippage compared to the primary device (GD DYNO 200). Run performance was assessed primarily on the LifeFitness 95T and LifeFitness INT-SE4-BLKHF-14 treadmills, with the exception of rare instances of treadmill assessment on treadmills during travel (LifeFitness Elevation Series SE3; LifeFitness Club Series+ SL).

### Sleep Performance

DELTA001’s standard sleep regimen was a bedtime of 9:15pm, with estimated sleep time by 9:45pm, with a wake alarm of 5:45am-6:00pm depending on work schedule. The WHOOP, Garmin, and Apple Watch were all worn to sleep. A wrist strap that allowed for simultaneous wearing of the WHOOP (under wrist) and Apple Watch (top of wrist) on one hand (Fresh Strap Hybrid). The long-haul sleep travel regimen consisted of immediate sleep on take-off for evening flights, with meals consolidated to be taken on landing. Exercise was consistently undertaken in the morning of both evening and morning flights. In the event of consecutive flights consisting of early evening landings and late evening (red-eye) travel, intense exercise was also undertaken in between flights to allow for energy usage to enable subsequent sleep. For short haul flights, every effort was made to fly during the middle of day to reduce the need for early morning wake-ups/reduced morning exercise. Of note, melatonin (5mg) was taken sparingly, with an estimated 15 doses taken during the entire course of the study. The study was unable to confirm if it was effective. No prescription sleep medications were used.

### Gut Microbiome Dynamics

Stool was self-collected by DELTA001 based on PRE48 and POST48 fasting schedules as well as during the 3^rd^ and 5^th^ day of a 3MAD week in accordance with manufacturer specifications and submitted for processing (AM-002, Enhanced AMILI Gut Health Test Kit; Amili Pte. Ltd.). Stool samples were sent to Amili for laboratory processing via post mail and were then processed via Amili’s proprietary 16S rRNA sequencing pipeline[209]. Data described in Figs. 8, 9, and S17 were analyzed based on data (i.e., ASV values and associated matched microbial strains, FASTQ reads) provided by Amili.

Alpha diversity was assessed using three indices to characterize microbial diversity within individual samples. The Shannon diversity index was calculated to capture both species richness and evenness[210], using the formula:

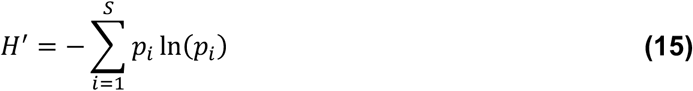

where *p_i_* is the relative abundance of each taxon. The Chao1 richness estimator was used to estimate total species richness[211], including undetected rare taxa, based on the formula:

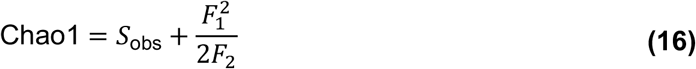

where *S*_obs_ is the number of observed taxa, and *F*_1_and *F*_2_are the numbers of singletons and doubletons, respectively. When no doubletons were present (i.e., *F*_2_ = 0), the bias-corrected formula was then applied[212]:

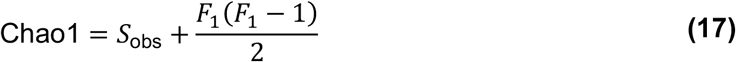

Finally, Simpson’s Diversity Index and its inverse were calculated with the formulas respectively to quantify species dominance and diversity[213]:

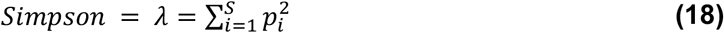

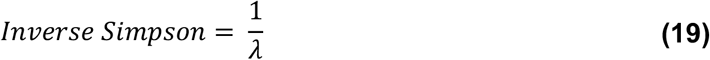

Custom Python scripts were used to compute each metric from ASV count data across samples, and results were exported for downstream analysis (pandas, numpy).

Demultiplexed sequences originally in FASTQ format were imported into QIIME2 (v2024.5)[214], trimmed using Cutadapt[215], and denoised using DADA2[216]. Representative sequences and feature tables were exported to PICRUSt2 (v2.5.3) for functional pathway prediction[217–221]. Normalization of pathway abundances was performed using predicted 16S rRNA copy numbers from PICRUSt2. Statistical analyses were performed including paired t-tests and Kruskal-Wallis tests with Dunn post-hoc analyses to identify significant functional pathways (p-value σ; 0.05). All analyses were performed with Python (pandas, SciPy, scikit-posthocs)[222].

### Healthspan Copilot GPT

The custom GPT ‘Healthspan Copilot’ was created using OpenAI’s Custom GPT interface. The GPT was configured through structured system instructions to deliver personalized, evidence-based guidance in the domain of healthy longevity (Supplementary File 2). Responses are constrained to be non-diagnostic, evidence-based, actionable, personalized, transparent, professional, and engaging. Three core GPT customizations were implemented. First, representative benchmark biomarker values from DELTA001 were included to enable comparative analysis of user data, with contextual interpretation based on fasting states. Second, the system was configured to recommend users track DELTA markers as individualized indicators of biological resilience. Third, the GPT incorporated a biological age estimation module with add-on analysis on biological resilience using user-input data. The biological age estimation module is constructed using a population dataset containing anonymized biomarker data (U.S. National Health and Nutrition Examination Survey, 2021–2023 cycle)[223]. The biological age of the population sample was estimated using ordinary least square regressions by eliminating insignificant variables from variables for which there is clinical evidence that they affect biological aging. The model includes up to 10 variables (HbA1c, weight, waist circumference, hs-CRP, HDL, systolic and diastolic blood pressure, pulse rate, gender, chronological age), and the respective biological age estimates for each individual. This output was compiled into a table (Supplementary File 2), and uploaded to the knowledge base of the GPT, enabling file-based retrieval augmentation during GPT responses. Any subset of the relevant biomarkers is compared to the relevant population subsample that matches the user’s race, using coefficients obtained from ordinary least squares estimates. The GPT estimates were validated by comparing them with the estimates from a statistical software package (Stata 18; StataCorp LLC).

### Participant Experience

This study adopted a descriptive case-study design, focusing on a single participant. DELTA001 was interviewed at three timepoints over the course of the study: at baseline, after a 3MAD routine, and at the end of the study. Semi-structured interviews were conducted, each lasting an average of 59 minutes (range: 40–71 minutes). All interviews were audio-recorded and transcribed verbatim. The interview prompts were designed to (1) explore the participant’s experiences, motivations, and challenges in maintaining the DELTA protocol; (2) to examine his engagement with digital technology; and (3) to assess the impact of changes in the fasting schedule on daily routine. A thematic analysis was conducted to identify key patterns across the interviews. Transcripts are provided in Supplementary File 3.

## DATA AVAILABILITY

All the data supporting the findings of the study are available in the manuscript and in the supplementary information (Tables S1-S5, Figs. S1-S18, Supplementary Data 1-2, and Supplementary Files 1-3). Data visualization and statistical analyses were performed either on GraphPad Prism 10.4.2 (GraphPad Software Inc.) or via Python (scikit-posthocs)[222].

## ACKNOWLEDGEMENTS

The DELTA team gratefully acknowledges the community of researchers engaged in studies that are relevant to DELTA findings. Due to the multiple parameters studied as part of the DELTA trial, we acknowledge the continuously growing body of work that we were inadvertently unable to cite due to space constraints. The DELTA study team gratefully acknowledges Prof. Jeremy Lim, Dr. Germaine Yong, Dr. Shiang Chiet Tan, Dr. Jeannie Lee, Jonathan San, the AMILI Health team, and the AMILI Lab team for assistance with microbiome data.

## AUTHOR CONTRIBUTIONS

P.W. led the acquisition of funding (NUS-NUHS HPHSR Enabling Grant Call) to support the DELTA study, conceived the study, contributed to the mathematical analysis of metabolic flexibility and digital biomarkers, and oversaw DELTA’s study design, data collection, and data analysis. N.F. performed in-depth analysis of fitness performance, sleep performance, and cardiovascular health, and co-led microbiome functional pathway abundance predictions. C.S. led the analysis of cardiometabolic health and digital biomarkers derived from blood markers and developed the GPT-enabled Healthspan Copilot framework. N.Y.T.L. led the microbiome analysis including microbiome compositions and diversity, co-led microbiome functional pathway abundance predictions, and contributed to the qualitative assessment of participant experience. S.W.S analyzed sleep performance, performed comparisons across wearable devices, and led the qualitative assessment of participant experience. G.S. contributed to the development of GPT-enabled Healthspan Copilot framework. N.F., N.Y.T.L., S.W.S., Y.H., and J.W.J.L. helped acquired funding (NUS-NUHS HPHSR Enabling Grant Call) to support the DELTA study. Y.S. provided insights into the economic implications of the study. P.W., N.F., C.S., N.Y.T.L., S.W.S., G.S., Y.S., L.H., A.W., Y.H.O., X.T., and D.H. wrote early drafts. P.W., N.F., C.S., N.Y.T.L., S.W.S., and D.H. created figures for the manuscript. L.H., A.W., P.R., H.P. contributed to scientific discussions and critical review of the final manuscript. H.S.J.C., L.Y.T.W., and J.W.J.L. provided clinically relevant inputs and scientific discussions for the study. Y.H.O. and X.T. worked on the IRB and ethics approval. D.H. designed the DELTA001 protocol, analyzed the data, supervised the study team, and acquired funding to support this study. All authors contributed to the final draft of the manuscript. All co-first authors independently and equally contributed to the DELTA study.

