## Supplementary Information for "DELTA: Fortifying Human Biological Resilience with an N=1 Digital Health and Dynamic Biomarker Protocol"

This Supplementary Information includes:

- Tables S1-S5
- Figs. S1-S18
- Supplementary Data 1-2
- Supplementary Files 1-3

**Table S1**. DELTA001's supplement regimen. The compounds and their dosing schedules are summarized below.

| Compound | Dose/Unit | Regimen | Brand |
|---|---|---|---|
| Creatine Monohydrate | 500mg/capsule | 3g/day (6 capsules) | Life Extension |
| Omega-3 | 1000mg/softgel (540mg EPA/360mg DHA) | 3000mg/day (3 Gels) | Now |
| Vitamin B Complex | Per 2 capsules. Thiamine (Vitamin B1) (as thiamine HCl) — 100 mg<br>Riboflavin (Vitamin B2) (as riboflavin and riboflavin 5'-phosphate) — 75 mg<br>Niacin (as niacinamide and niacin) — 100 mg<br>Vitamin B6 (as pyridoxine HCl and pyridoxal 5'-phosphate) — 100 mg<br>Folate (as L-5-methyltetrahydrofolate, calcium salt) — 680 mcg DFE<br>Vitamin B12 (as methylcobalamin) — 300 mcg<br>Biotin — 1000 mcg<br>Pantothenic acid (as D-calcium pantothenate) — 500 mg<br>Calcium (as D-calcium pantothenate, dicalcium phosphate) — 50 mg<br>Inositol — 100 mg<br>PABA (para-aminobenzoic acid) — 50 mg | 2 capsules/day | Life Extension |
| Multivitamin (One a Day) | Vitamin A (100% as beta-carotene), 900 mcg<br>Vitamin C, 99 mg<br>Vitamin D, 25 mcg (1000 IU)<br>Vitamin E, 15 mg<br>Thiamin (B1), 1.32 mg<br>Riboflavin (B2), 1.43 mg<br>Niacin, 17.6 mg<br>Vitamin B6, 2.17 mg<br>Folate (240 mcg folic acid), 400 mcg DFE<br>Vitamin B12, 6.24 mcg<br>Biotin, 43 mcg<br>Pantothenic Acid, 15.5 mg<br>Calcium, 210 mg<br>Iron, 0 mg<br>Iodine, 150 mcg<br>Magnesium, 120 mg<br>Zinc, 11 mg<br>Selenium, 55 mcg<br>Copper, 0.9 mg<br>Manganese, 2.3 mg<br>Chromium, 35 mcg<br>Lycopene, 300 mcg | 1 tablet/day | Bayer (One a Day) |
| Vitamin D3 | 1000IU/gel | 2000 IU/day (2 gels) | Now |
| Plant Sterols | 400mg/gel | 2000 mg/day (5 gels) | Swansons |

| | | | |
|---|---|---|---|
| Curcumin C3 Complex | 500mg/capsule | 500 mg/day (1 capsule) | California Gold Nutrition - Strict adherence to recommended dose |
| Ubiquinol | 200mg/gel | 200 mg/day (1 gel) | Now |
| Quercetin/Bromelain | Per 2 capsules Quercetin 800mg<br>Bromeliad (2400 GDU/g). 165mg | 4 capsules/day Quercetin 1600mg Bromeliad (2400 GDU/g). 330mg | Now |
| Vitamin K2 (MK-7) | 100mcg/capsule | 100mcg/day (1 capsule) | Now |
| EGCg | 400mg/capsule | 400 mg/day (1 capsule) | Now - Strict adherence to recommended dose |
| Probiotic | Per 1 capsule<br>Lactobacillus acidophilus (La-14)<br>Bifidobacterium lactis (Bl-04)<br>Lactobacillus plantarum (Lp-115)<br>Lactobacillus casei (Lc-11)<br>Lactobacillus rhamnosus (Lr-32)<br>Lactobacillus paracasei (Lpc-37)<br>Bifidobacterium breve (Bb-03)<br>Streptococcus thermophilus (St-21)<br>Lactobacillus salivarius (Ls-33)<br>Bifidobacterium longum (Bl-05) | 100 billion CFU/day (1 capsule) | Now |

**Table S2.** Summary of biomarkers measured in the DELTA protocol.

| Biomarker | Purpose | Collection Method |
|---|---|---|
| Heart Rate Variability (HRV) | Stress resilience and recovery | Wearables |
| Resting Heart Rate (RHR) | Heart fitness and cardiovascular health | Wearables |
| Blood Pressure | Heart health and risks for cardiovascular diseases | Omron IntelliSense BP7350 |
| Grip Strength | Physical health (e.g., muscle function, frailty) and longevity | Camry EH101 |
| Blood glucose | Carbohydrate metabolism and blood sugar regulation | Finger stick (Abbott Optium Neo) |
| Blood ketones | Fat metabolism and ketosis | Finger stick (Abbott Optium Neo) |
| HbA1c | Indicator of diabetes risk | Abbott Libre |
| Apolipoprotein B (ApoB) | Cardiovascular risk, a better indicator of 'bad' cholesterol than LDL | Blood test |
| Apolipoprotein A (ApoA) | Cardiovascular protection, a better indicator of 'good' cholesterol than HDL | Blood test |
| High-sensitivity C-reactive protein (hs-CRP) | Systemic inflammation | Blood test |
| Homocysteine | Methylation and B-vitamin status | Blood test |
| Gut Microbiome | Composition of gut microbiome as indicator of overall health | 16S rRNA gene sequencing (Amili Pte. Ltd.) |

**Table S3.** Duration of sleep stages recorded with Garmin, Apple Watch, and WHOOP devices.

| Devices | Deep Sleep (Mins) | Light Sleep (Mins) | REM Sleep (Mins) | Awake (Mins) |
|---|---|---|---|---|
| Garmin | 79.98 | 287.7 | 104.3 | 20.03 |
| Apple | 40.92 | 342.3 | 99.91 | 16.34 |
| WHOOP | 93.55 | 222.5 | 120.8 | 53.88 |

**Table S4. SPIRIT extension and elaboration of N-of-1 trials (SPENT) checklist for N-of-1 protocol reporting guidelines**

| SECTION / TOPIC | ITEM NO. | SPIRIT 2013 | ITEM NO. | SPENT 2019 | ON PAGE |
|---|---|---|---|---|---|
| **SECTION 1: ADMINISTRATIVE DATA** | | | | | |
| Title | 1 | Descriptive title identifying the study design, population, interventions, and, if applicable, trial acronym. | 1a | Descriptive title, including "N-of-1 trial" and "protocol". ***For series:*** Descriptive title, including "a series of N-of-1 trials" and "protocol". | Pg.1 |
| | | — | 1b | For specific guidance on abstracts, see SPENT Guidance for Abstracts. (Appendix table 2) | N/A |
| Trial Registration | 2a | Trial identifier and registry name. If not yet registered, name of intended registry. | 2a | (no change) | Pg.20 |
| | 2b | All items from the World Health Organization Trial Registration Data Set. (WHOTRDS) | 2b | (no change) | N/A |
| Protocol version | 3 | Date and version identifier. | 3 | (no change) | N/A |
| Funding | 4 | Sources and types of financial, material, and other support. | 4 | (no change) | Pg.24 |
| Roles and responsibilities | 5a | Names, affiliations, and roles of protocol contributors. | 5a | (no change) | Pg.24-25 |
| | 5b | Name and contact information for the trial sponsor. | 5b | (no change) | N/A |
| | 5c | Role of study sponsor and funders, if any, in study design; collection, management, analysis, and interpretation of data; writing of the report; and the decision to submit the report for publication, including whether they will have ultimate authority over any of these activities. | 5c | (no change) | N/A |
| | 5d | Composition, roles, and responsibilities of the coordinating center, steering committee, end point adjudication committee, data management team, and other individuals or groups overseeing the trial, if applicable. (see Item 21a for DMC) | 5d | (no change) | N/A |
| **SECTION 2: INTRODUCTION** | | | | | |
| Background and rationale | 6a | Description of research question and justification for undertaking the trial, including summary of relevant studies (published and unpublished) examining benefits and harms for each intervention. | 6a | Description of research question and justification for undertaking the trial, including summary of relevant studies (published and unpublished) examining benefits and harms for each intervention, and rationale for using N-of-1. | Pg.2-4 |

| | | | | |
|---|---|---|---|---|
| | 6b | Explanation for choice of comparators. | 6b | (no change, SPENT commentary) |
| Objectives | 7 | Specific objectives or hypotheses. | 7 | (no change) |
| Trial design | 8 | Description of trial design, including type of trial (eg, parallel group, crossover, factorial, single group), allocation ratio, and framework (eg, superiority, equivalence, non-inferiority, exploratory). | 8 | Description of the trial design, including N-of-1 trial or series of trials, and framework (eg, superiority, equivalence, non-inferiority, exploratory). ***In addition for series:*** Explanation of the series design including whether the design will be tailored to each participant. |
| **SECTION 3: METHODS** | | | | |
| ***Participants, interventions, and outcomes*** | | | | |
| Study Setting | 9 | Description of study settings (eg, community clinic, academic hospital) and list of countries where data will be collected. Reference to where list of study sites can be obtained. | 9 | (no change) |
| Eligibility criteria | 10 | Inclusion and exclusion criteria for participants. If applicable, eligibility criteria for study centers and individuals who will perform the interventions (eg, surgeons, psychotherapists). | 10 | (in addition) Diagnosis/disorder, diagnostic criteria, co-morbid conditions and concurrent therapies. ***For series:*** Same as SPIRIT item 10. |
| Interventions | 11a | Interventions for each group with sufficient detail to allow replication, including how and when they will be administered. | 11a | Intervention(s) for each period with sufficient detail to allow replication, including how and when they will be administered, planned number of periods, and duration of each period (including run-in and washout, if applicable). ***In addition for series:*** How the design will be tailored to each participant, if applicable. |
| | 11b | Criteria for discontinuing or modifying allocated interventions for a given trial participant (eg, drug dose change in response to harms, participant request, or improving/ worsening disease). | 11b | (no change, SPENT commentary) |
| | 11c | Strategies to improve adherence to intervention protocols, and any procedures for monitoring adherence (eg, drug tablet return, laboratory tests). | 11c | (no change) |
| | 11d | Relevant concomitant care and interventions that are permitted or prohibited during the trial. | 11d | (no change) |
| Outcomes | 12 | Primary, secondary, and other outcomes, including the specific measurement variable (eg, systolic blood pressure), analysis metric (eg, change from baseline, final value, time to event), | 12 | (no change) |

| | | | | |
|---|---|---|---|---|
| | | method of aggregation (eg, median, proportion), and time point for each outcome. Explanation of the clinical relevance of chosen efficacy and harm outcomes is strongly recommended. | | |
| Participant timeline | 13 | Time schedule of enrollment, interventions (including any run-ins and washouts), assessments, and visits for participants. A schematic diagram is highly recommended (Figure). | 13 | (no change) |
| Sample size | 14 | Estimated number of participants needed to achieve study objectives and how it was determined, including clinical and statistical assumptions supporting any sample size calculations. | 14 | Estimated number of intervention periods and measurements/observations needed to achieve study objectives within an individual N-of-1 trial. ***In addition for series:*** Estimated number of participants needed to achieve study objectives. How these numbers were determined, including clinical and statistical assumptions supporting any sample size calculations. |
| Recruitment | 15 | Strategies for achieving adequate participant enrollment to reach target sample size. | 15 | ***For series:*** Strategies for achieving adequate participant enrollment to reach target sample size. |
| ***Assignment of interventions (for controlled trials)*** | | | | |
| *Allocation* sequence generation | 16a | Method of generating the allocation sequence (eg, computer-generated random numbers), and list of any factors for stratification. To reduce predictability of a random sequence, details of any planned restriction (eg, blocking) should be provided in a separate document that is unavailable to those who enrol participants or assign interventions. | 16a | Method of generating the allocation sequence (eg, computer-generated random numbers), and list of any factors for stratification. To reduce predictability, details of any restrictions (eg, pairs, blocking) should be provided in a separate document that is unavailable to those who enrol participants or assign interventions. ***In addition for series:*** List of any factors for stratification. |
| concealment mechanism | 16b | Mechanism of implementing the allocation sequence (eg, central telephone; sequentially numbered, opaque, sealed envelopes), describing any steps to conceal the sequence until interventions are assigned. | 16b | (no change, SPENT commentary) |
| implementation | 16c | Who will generate the allocation sequence, who will enroll participants, and who will assign participants to interventions. | 16c | (no change) |
| Blinding (masking) | 17a | Who will be blinded after assignment to interventions (eg, trial participants, care providers, outcome assessors, data analysts), and how. | 17a | (no change) |

| | | | | |
|---|---|---|---|---|
| | 17b | If blinded, circumstances under which unblinding is permissible, and procedure for revealing a participant's allocated intervention during the trial. | 17b | (no change) |
| ***Data collection, management, and analysis*** | | | | |
| Data collection methods | 18a | Plans for assessment and collection of outcome, baseline, and other trial data, including any related processes to promote data quality (eg, duplicate measurements, training of assessors) and a description of study instruments (eg, questionnaires, laboratory tests) along with their reliability and validity, if known. Reference to where data collection forms can be found, if not in the protocol. | 18a | (no change) |
| | 18b | Plans to promote participant retention and complete follow-up, including list of any outcome data to be collected for participants who discontinue or deviate from intervention protocols. | 18b | (no change) |
| Data management | 19 | Plans for data entry, coding, security, and storage, including any related processes to promote data quality (eg, double data entry; range checks for data values). Reference to where details of data management procedures can be found, if not in the protocol. | 19 | (no change) |
| Statistical methods | 20a | Statistical methods for analyzing primary and secondary outcomes. Reference to where other details of the statistical analysis plan can be found, if not in the protocol. | 20a1 | Statistical methods for analyzing primary and secondary outcomes for each individual. Reference to where other details of the statistical analysis plan can be found, if not in the protocol. ***In addition for series:*** if planned, proposed methods of quantitative synthesis of individual trial data, and how heterogeneity between participants will be assessed |
| | | | 20a2 | Statistical methods to account for correlation introduced by the repeated measures and crossover design of N-of-1 studies. |
| | 20b | Methods for any additional analyses (eg, subgroup and adjusted analyses). | 20b | ***For series:*** Methods for any additional analyses (eg, subgroup and adjusted analyses). |
| | 20c | Definition of analysis population relating to protocol non-adherence (eg, as-randomized | 20c | Statistical methods to handle missing data (eg, multiple imputation, modelling). ***In addition for series:*** Definition of analysis population relating |

| | | | | |
|---|---|---|---|---|
| | | analysis), and any statistical methods to handle missing data (eg, multiple imputation). | | to protocol non-adherence (eg, as-randomized analysis). |
| **Monitoring** | | | | |
| Data monitoring | 21a | Composition of Data Monitoring Committee (DMC); summary of its role and reporting structure; statement of whether it is independent from the sponsor and competing interests; and reference to where further details about its charter can be found, if not in the protocol. Alternatively, an explanation of why a DMC is not needed. | 21a | (no change, SPENT commentary) |
| | 21b | Description of any interim analyses and stopping guidelines, including who will have access to these interim results and make the final decision to terminate the trial. | 21b | (no change, SPENT commentary) |
| Harms | 22 | Plans for collecting, assessing, reporting, and managing solicited and spontaneously reported adverse events and other unintended effects of trial interventions or trial conduct. | 22 | (no change, SPENT commentary) |
| Auditing | 23 | Frequency and procedures for auditing trial conduct, if any, and whether the process will be independent from investigators and the sponsor. | 23 | (no change) |
| **SECTION 4: ETHICS & DISSEMINATION** | | | | |
| Research ethics approval | 24 | Plans for seeking REC/IRB approval. | 24 | (no change, SPENT commentary) |
| Protocol amendments | 25 | Plans for communicating important protocol modifications (eg, changes to eligibility criteria, outcomes, analyses) to relevant parties (eg, investigators, RECs/IRBs, trial participants, trial registries, journals, regulators). | 25 | (no change) |
| Consent or assent | 26a | Who will obtain informed consent or assent from potential trial participants or authorized surrogates, and how (see item 32). | 26a | (no change) |
| | 26b | Additional consent provisions for collection and use of participant data and biological specimens in ancillary studies, if applicable. | 26b | (no change) |
| Confidentiality | 27 | How personal information about potential and enrolled participants will be collected, shared, | 27 | (no change, SPENT commentary) |

| | | | | |
|---|---|---|---|---|
| | | and maintained in order to protect confidentiality before, during, and after the trial. | | |
| Declaration of interests | 28 | Financial and other competing interests for principal investigators for the overall trial and each study site. | 28 | (no change) |
| Access to data | 29 | Statement of who will have access to the final trial data set, and disclosure of contractual agreements that limit such access for investigators. | 29 | (no change) |
| Ancillary and post-trial care | 30 | Provision, if any, for ancillary and post-trial care, and for compensation to those who suffer harm from trial participation. | 30 | (no change) |
| Dissemination policy | 31a | Plans for investigators and sponsor to communicate trial results to participants, health care professionals, the public, and other relevant groups (eg, via publication, reporting in results databases, or other data-sharing arrangements), including any publication restrictions. | 31a | Plans for investigators to communicate each individual's results to the participant. Plans for investigators and sponsor to communicate trial results to participants, health care professionals, the public, and other relevant groups (eg, via publication, reporting in results databases, or other data-sharing arrangements), including any publication restrictions. |
| | 31b | Authorship eligibility guidelines and any intended use of professional writers. | 31b | (no change) |
| | 31c | Plans, if any, for granting public access to the full protocol, participant-level data set, and statistical code. | 31c | (no change) |
| **SECTION 5: APPENDICES** | | | | |
| Informed consent materials | 32 | Model consent form and other related documentation given to participants and authorized surrogates. | 32 | (no change) |
| Biological specimens | 33 | Plans for collection, laboratory evaluation, and storage of biological specimens for genetic or molecular analysis in the current trial and for future use in ancillary studies, if applicable. | 33 | (no change) |

**Table S5.** CENT 2015 checklist*; CONSORT 2010 checklist items with modifications or additions for individual or series of N-of-1 trials; empty items in the CENT 2015 column indicate no modification from the CONSORT 2010 item

| Section/ Topic | CONSORT 2010 No | CONSORT 2010 Item | CENT 2015 No | CENT 2015 Item | On Page |
|---|---|---|---|---|---|
| **Title and abstract** | | | | | |
| | 1a | Identification as a randomised trial in the title | 1a | Identify as an "N-of-1 trial" in the title<br>***For series:*** Identify as "a series of N-of-1 trials" in the title | Pg. 1 |
| | 1b | Structured summary of trial design, methods, results, and conclusions (for specific guidance see CONSORT for abstracts) | 1b | *For specific guidance, see CENT guidance for abstracts (table 2)* | N/A |
| **Introduction** | | | | | |
| Background and objectives | 2a | Scientific background and explanation of rationale | 2a.1 | | Pg. 2-4 |
| | | | 2a.2 | Rationale for using N-of-1 approach | Pg. 3 |
| | 2b | Specific objectives or hypotheses | 2b | | Pg. 3-4 |
| **Methods** | | | | | |
| Trial design | 3a | Description of trial design (such as parallel, factorial) including allocation ratio | 3a | Describe trial design, planned number of periods, and duration of each period (including run-in and wash out, if applicable)<br>***In addition for series:*** Whether and how the design was individualized to each participant, and explain the series design | Pg. 20-24 |
| | 3b | Important changes to methods after trial start (such as eligibility criteria), with reasons | 3b | | N/A |
| Participant(s) | 4a | Eligibility criteria for participants | 4a | Diagnosis or disorder, diagnostic criteria, comorbid conditions, and concurrent therapies.<br>***For series:*** Same as CONSORT item 4a | Pg. 20 |

| | | | | | |
|---|---|---|---|---|---|
| | 4b | Settings and locations where the data were collected | 4b | | Pg. 20 |
| | | | 4c | Whether the trial(s) represents a research study and if so, whether institutional ethics approval was obtained | Pg. 20 |
| Interventions | 5 | The interventions for each group with sufficient details to allow replication, including how and when they were actually administered | 5 | The interventions for each period with sufficient details to allow replication, including how and when they were actually administered | Pg. 20-24 |
| Outcomes | 6a | Completely defined pre-specified primary and secondary outcome measures, including how and when they were assessed | 6a.1 | | Pg. 2-4 |
| | | | 6a.2 | Description and measurement properties (validity and reliability) of outcome assessment tools | Pg. 4 |
| | 6b | Any changes to trial outcomes after the trial commenced, with reasons | 6b | | N/A |
| Sample size | 7a | How sample size was determined | 7a | | N/A |
| | 7b | When applicable, explanation of any interim analyses and stopping guidelines | 7b | | N/A |
| Randomisation: | | | | | |
| Sequence generation | 8a | Method used to generate the random allocation sequence | 8a | Whether the order of treatment periods was randomised, with rationale, and method used to generate allocation sequence | N/A |
| | 8b | Type of randomisation; details of any restriction (such as blocking and block size) | 8b | When applicable, type of randomisation; details of any restrictions (such as pairs, blocking) | N/A |
| | | | 8c | Full, intended sequence of periods | N/A |
| Allocation concealment mechanism | 9 | Mechanism used to implement the random allocation sequence (such as sequentially numbered containers), describing any steps taken to conceal the sequence until interventions were assigned | 9 | | N/A |
| Implementation | 10 | Who generated the random allocation sequence, who enrolled participants, and who assigned participants to interventions | 10 | | N/A |

| | | | | | |
|---|---|---|---|---|---|
| Blinding | 11a | If done, who was blinded after assignment to interventions (for example, participants, care providers, those assessing outcomes) and how | 11a | | N/A |
| | 11b | If relevant, description of the similarity of interventions | 11b | | N/A |
| Statistical methods | 12a | Statistical methods used to compare groups for primary and secondary outcomes | 12a | Methods used to summarize data and compare interventions for primary and secondary outcomes | Pg. 20-24 |
| | 12b | Methods for additional analyses, such as subgroup analyses and adjusted analyses | 12b | ***For series:*** If done, methods of quantitative synthesis of individual trial data, including subgroup analyses, adjusted analyses, and how heterogeneity between participants was assessed, (for specific guidance on reporting syntheses of multiple trials, please consult the PRISMA Statement) | N/A |
| | | | 12c | Statistical methods used to account for carryover effect, period effects, and intra-subject correlation | N/A |
| **Results** | | | | | |
| Participant flow (a diagram is strongly recommended) | 13a | For each group, the numbers of participants who were randomly assigned, received intended treatment, and were analysed for the primary outcome | 13a.1 | Number and sequence of periods completed, and any changes from original plan with reasons | N/A |
| | | | 13a.2 | ***For series:*** The number of participants who were enrolled, assigned to interventions, and analysed for the primary outcome | N/A |
| | 13b | For each group, losses and exclusions after randomisation, together with reasons | 13c | ***For series:*** losses or exclusions of participants after treatment assignment, with reasons, and period in which this occurred, if applicable | N/A |
| Recruitment | 14a | Dates defining the periods of recruitment and follow-up | 14a | | N/A |
| | 14b | Why the trial ended or was stopped | 14b | Whether any periods were stopped early and/or whether trial was stopped early, with reason(s). | N/A |
| Baseline data | 15 | A table showing baseline demographic and clinical characteristics for each group | 15† | | N/A |
| Numbers analysed | 16 | For each group, number of participants (denominator) included in each analysis and | 16 | For each intervention, number of periods analysed. | N/A |

| | | | | | |
|---|---|---|---|---|---|
| | | whether the analysis was by original assigned groups | | ***In addition for series:*** if quantitative synthesis was performed, number of trials for which data were synthesized | |
| Outcomes and estimation | 17a | For each primary and secondary outcome, results for each group, and the estimated effect size and its precision (such as 95% confidence interval) | 17a.1 | For each primary and secondary outcome, results for each period; an accompanying figure displaying the trial data is recommended. | Pg. 4-11, Pg. 36-44 Pg. 46 |
| | | | 17a.2 | For each primary and secondary outcome, the estimated effect size and its precision (such as 95% confidence interval)<br>***In addition for series:*** if quantitative synthesis was performed, group estimates of effect and precision for each primary and secondary outcome | N/A |
| | 17b | For binary outcomes, presentation of both absolute and relative effect sizes is recommended | 17b | | N/A |
| Ancillary analyses | 18 | Results of any other analyses performed, including subgroup analyses and adjusted analyses, distinguishing pre-specified from exploratory | 18 | Results of any other analyses performed, including assessment of carryover effects, period effects, intra-subject correlation<br>***In addition for series:*** If done, results of subgroup or sensitivity analyses | N/A |
| Harms | 19 | All important harms or unintended effects in each group (for specific guidance see CONSORT for harms) | 19 | All harms or unintended effects for each intervention. *(for specific guidance see CONSORT for harms)* | N/A |
| **Discussion** | | | | | |
| Limitations | 20 | Trial limitations, addressing sources of potential bias, imprecision, and, if relevant, multiplicity of analyses | 20 | | Pg. 18-19 |
| Generalisability | 21 | Generalisability (external validity, applicability) of the trial findings | 21 | | Pg. 18-19 |
| Interpretation | 22 | Interpretation consistent with results, balancing benefits and harms, and considering other relevant evidence | 22 | | Pg. 18-19 |
| **Other information** | | | | | |

| | | | | |
|---|---|---|---|---|
| Registration | 23 | Registration number and name of trial registry | 23 | Pg. 20 |
| Protocol | 24 | Where the full trial protocol can be accessed, if available | 24 | N/A |
| Funding | 25 | Sources of funding and other support (such as supply of drugs), role of funders | 25 | Pg. 24 |

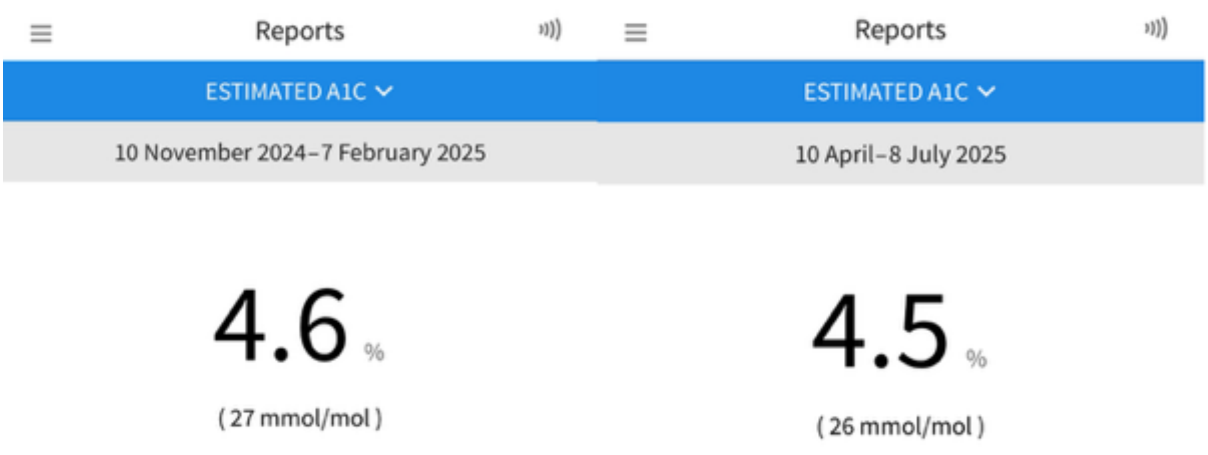

**Fig. S1**. Estimated HbA1C (mmol/mol) from CGM (Abbott Freestyle Libre).

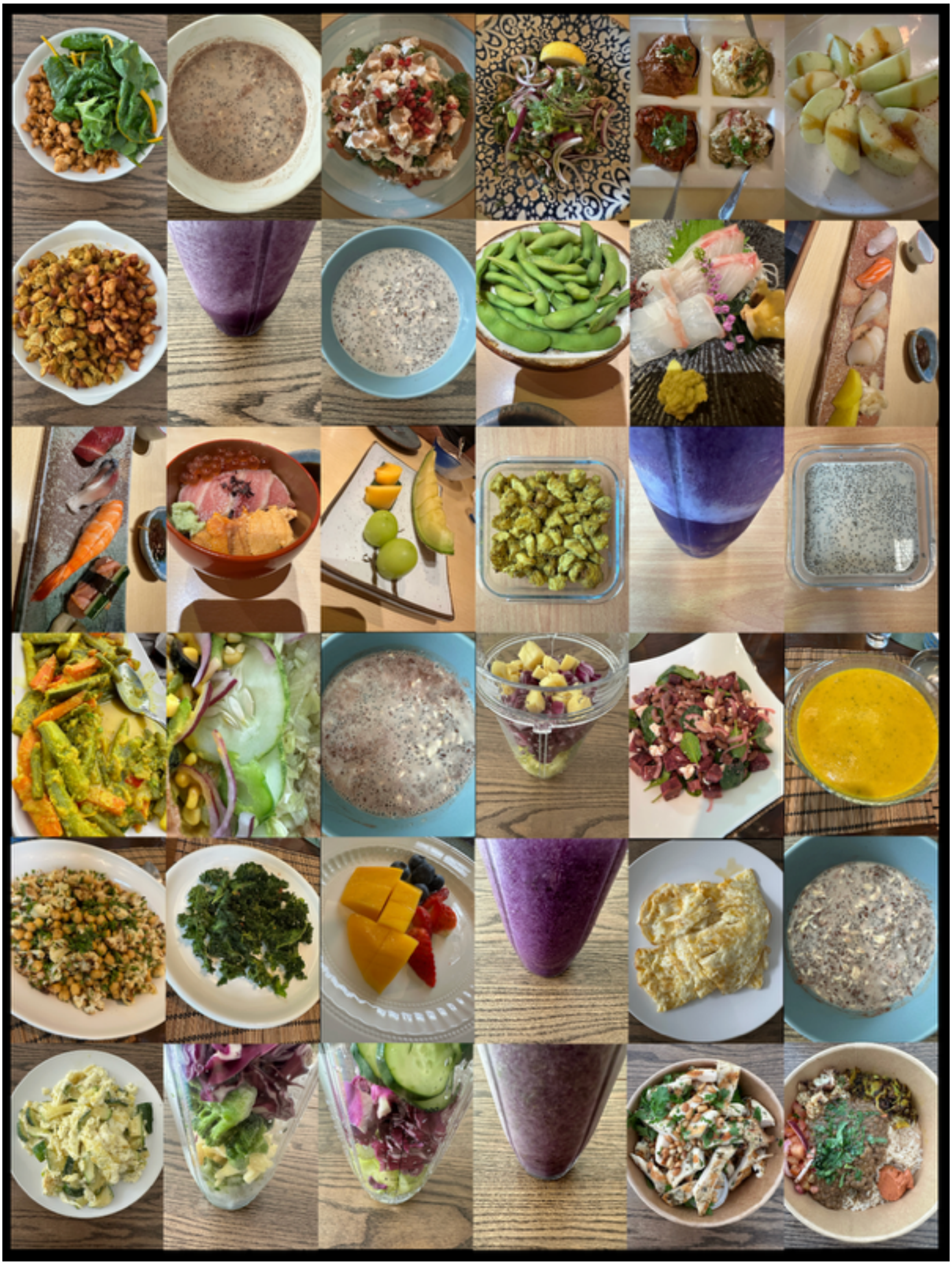

**Fig. S2.** DELTA001's dietary regimen from Oct 1-7 2024.

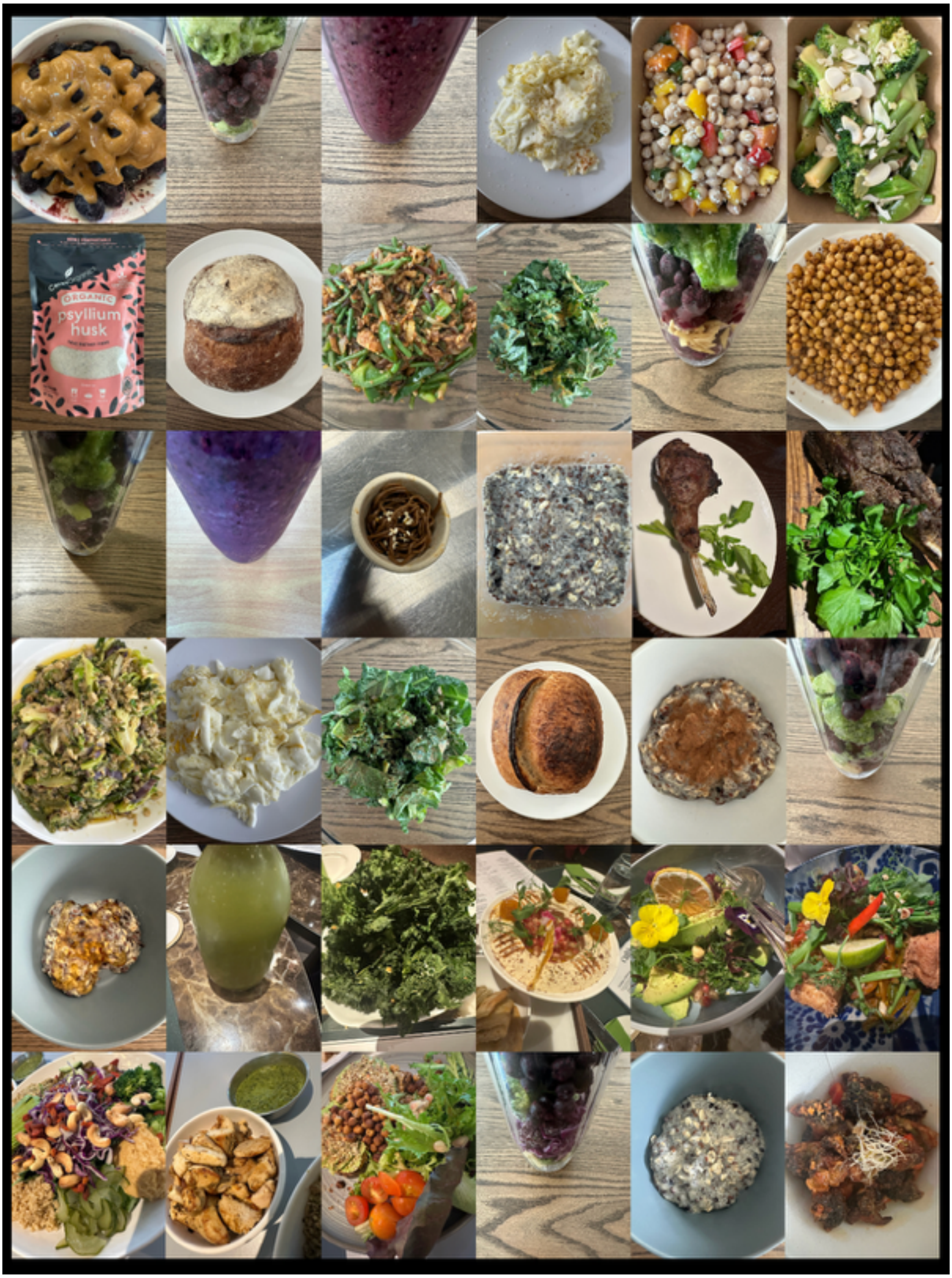

**Fig. S3.** DELTA001's dietary regimen from Dec 1-7 2024.

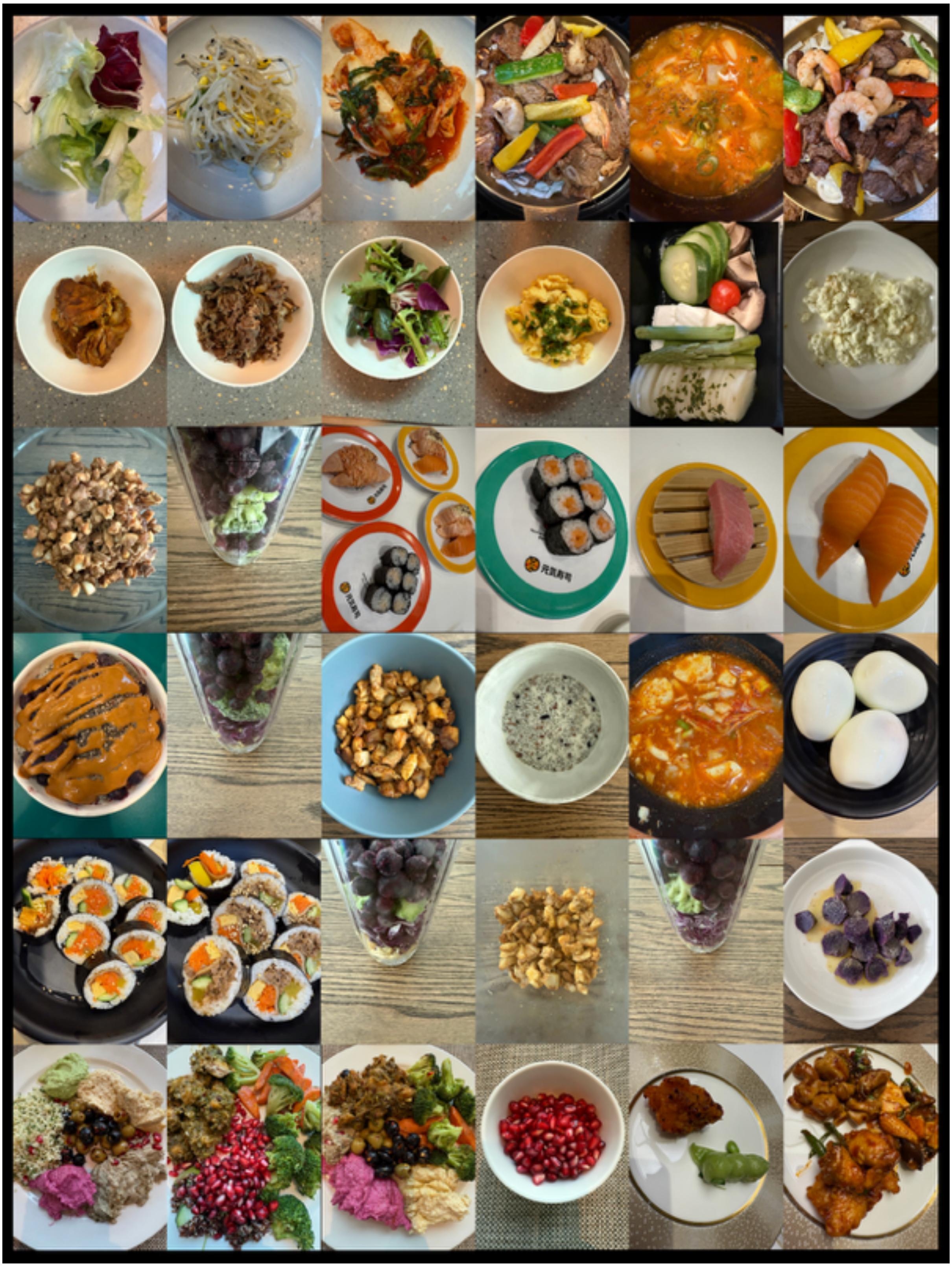

**Fig. S4.** DELTA001's dietary regimen from Jan 1-7 2025.

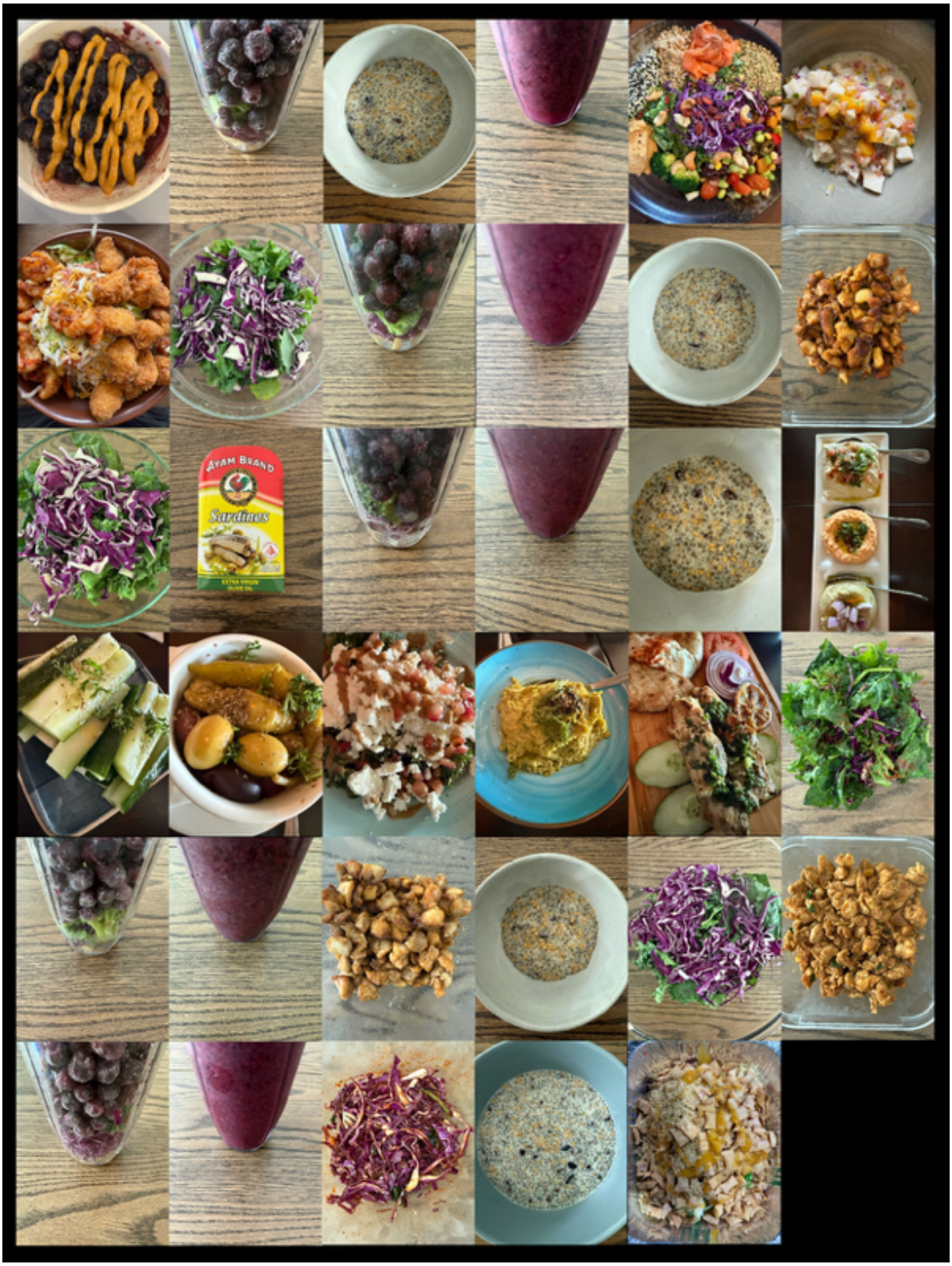

**Fig. S5.** DELTA001’s dietary regimen from Jan 1-7 2025.

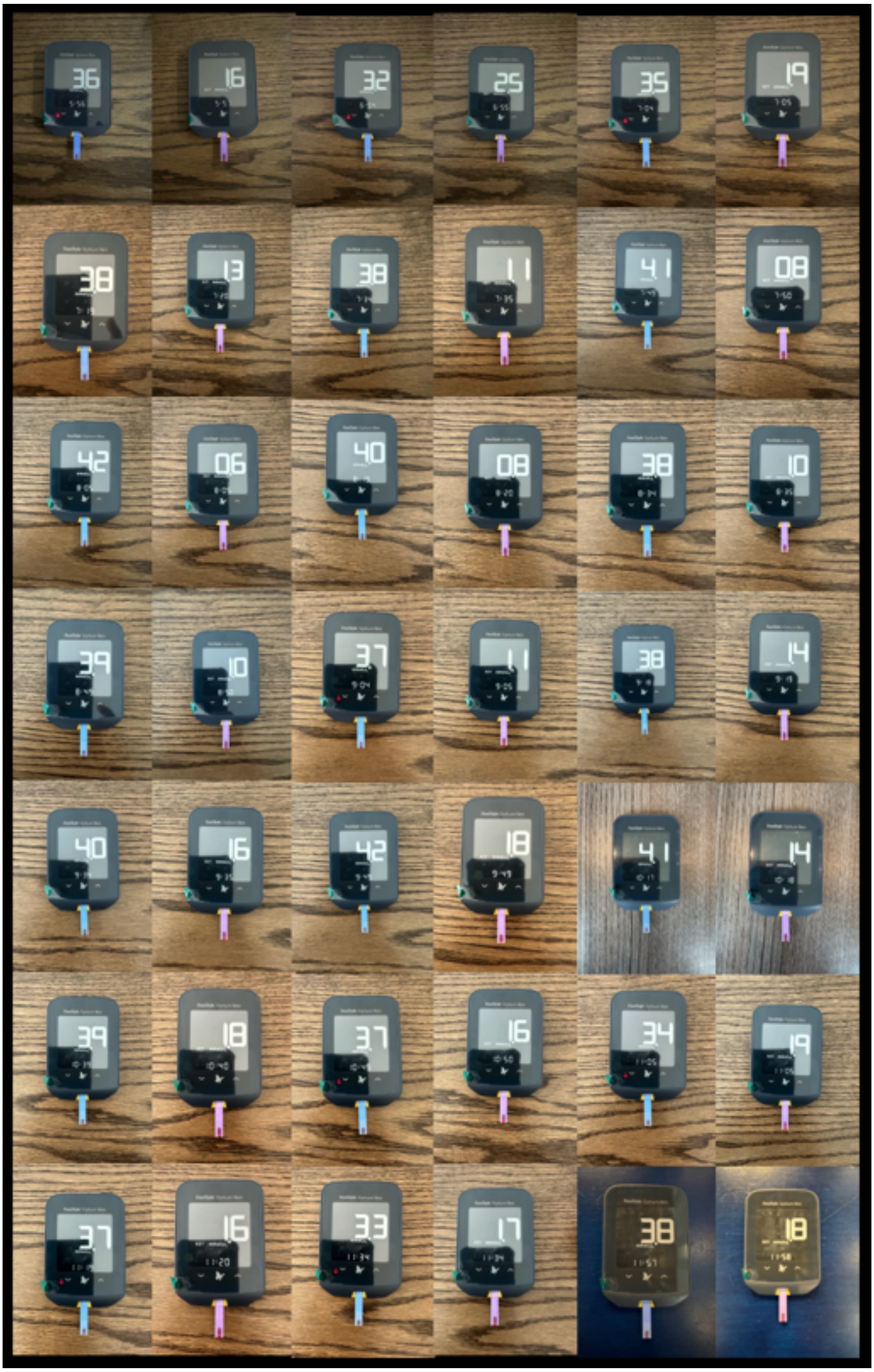

**Fig. S6.** Measured blood ketone and glucose levels for the 48-hour fasting strength training regimen on Sep 20 2024.

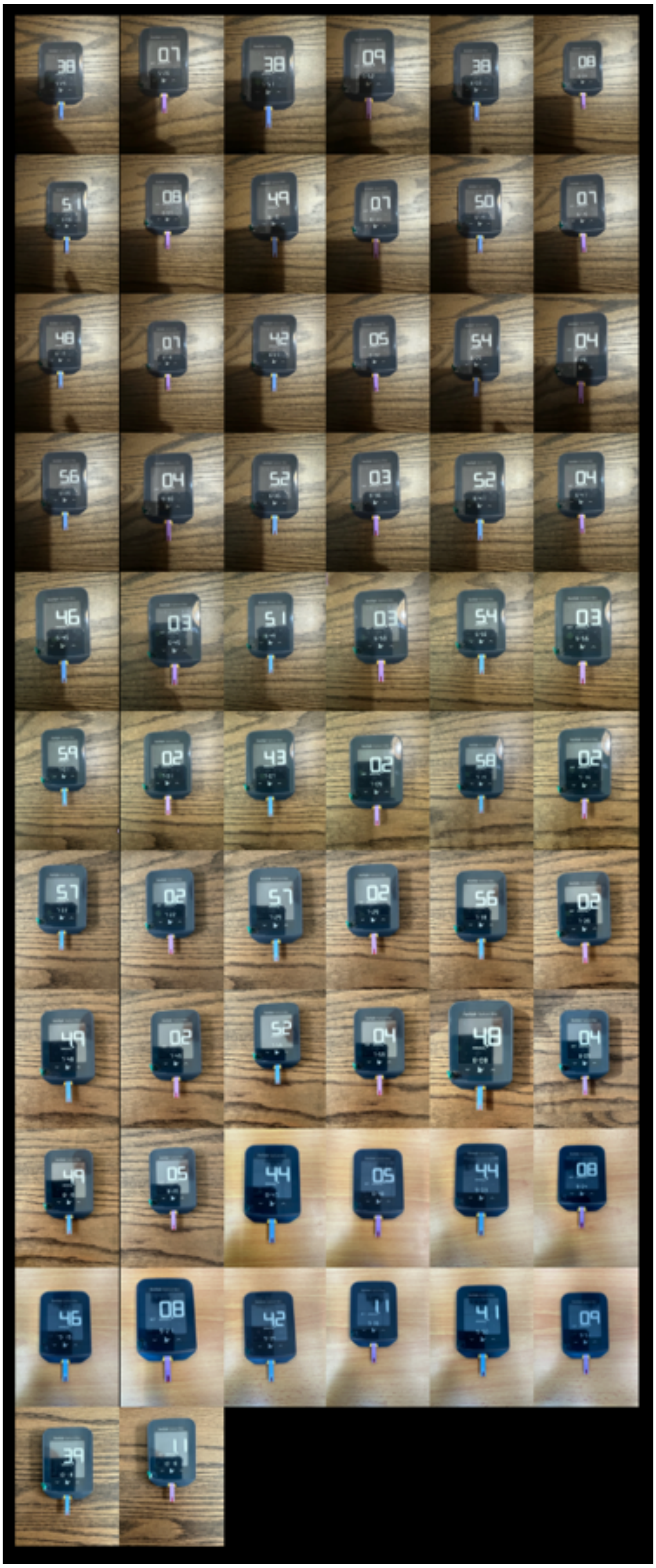

**Fig. S7.** Measured blood ketone and glucose levels for the 48-hour fasting strength training regimen on Oct 10 2024.

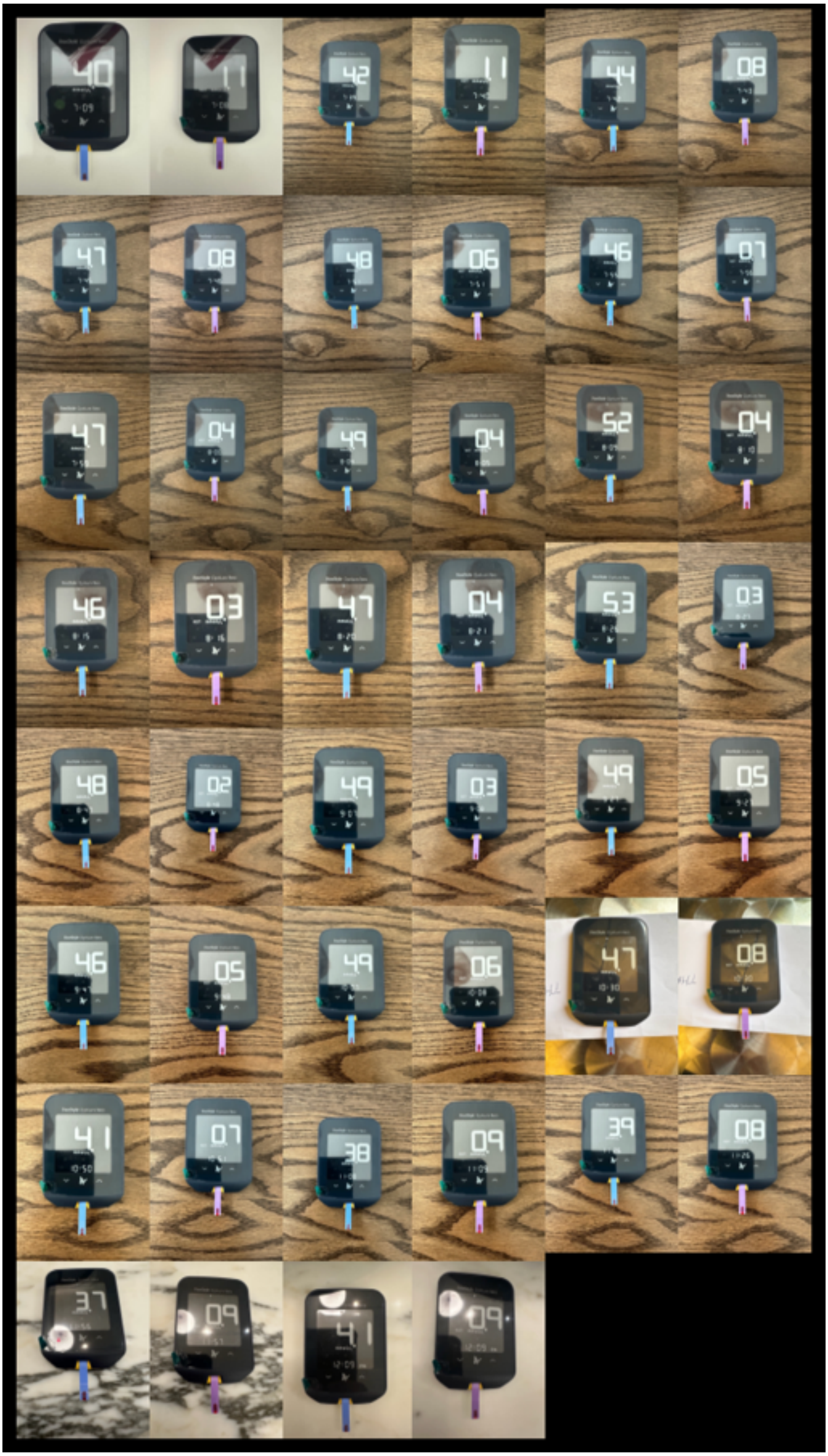

**Fig. S8.** Measured blood ketone and glucose levels for the 48-hour fasting strength training regimen on Oct 25 2024.

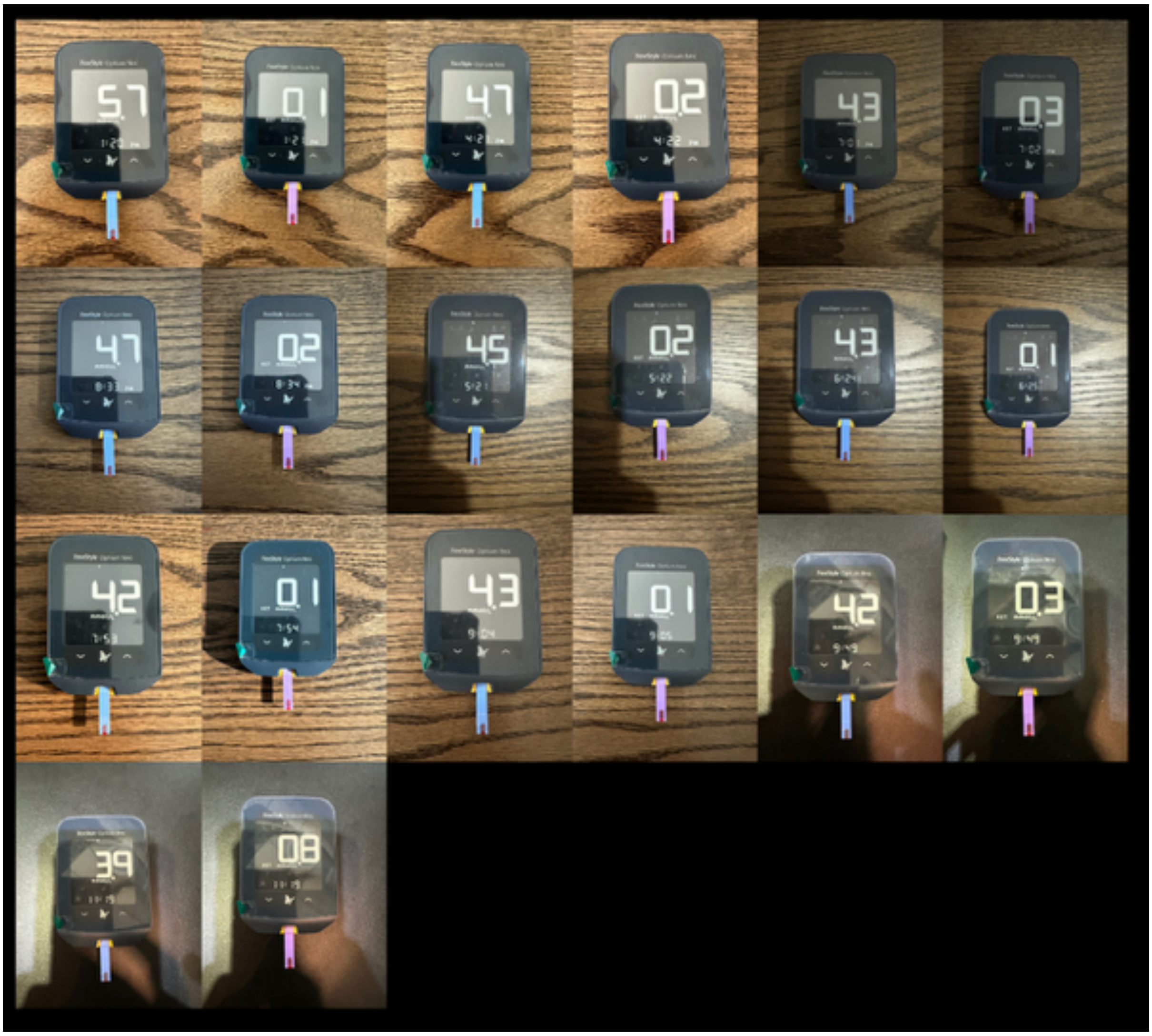

**Fig. S9.** Measured blood ketone and glucose levels for the OMAD ketosis entry regimen from Oct 27-28, 2024.

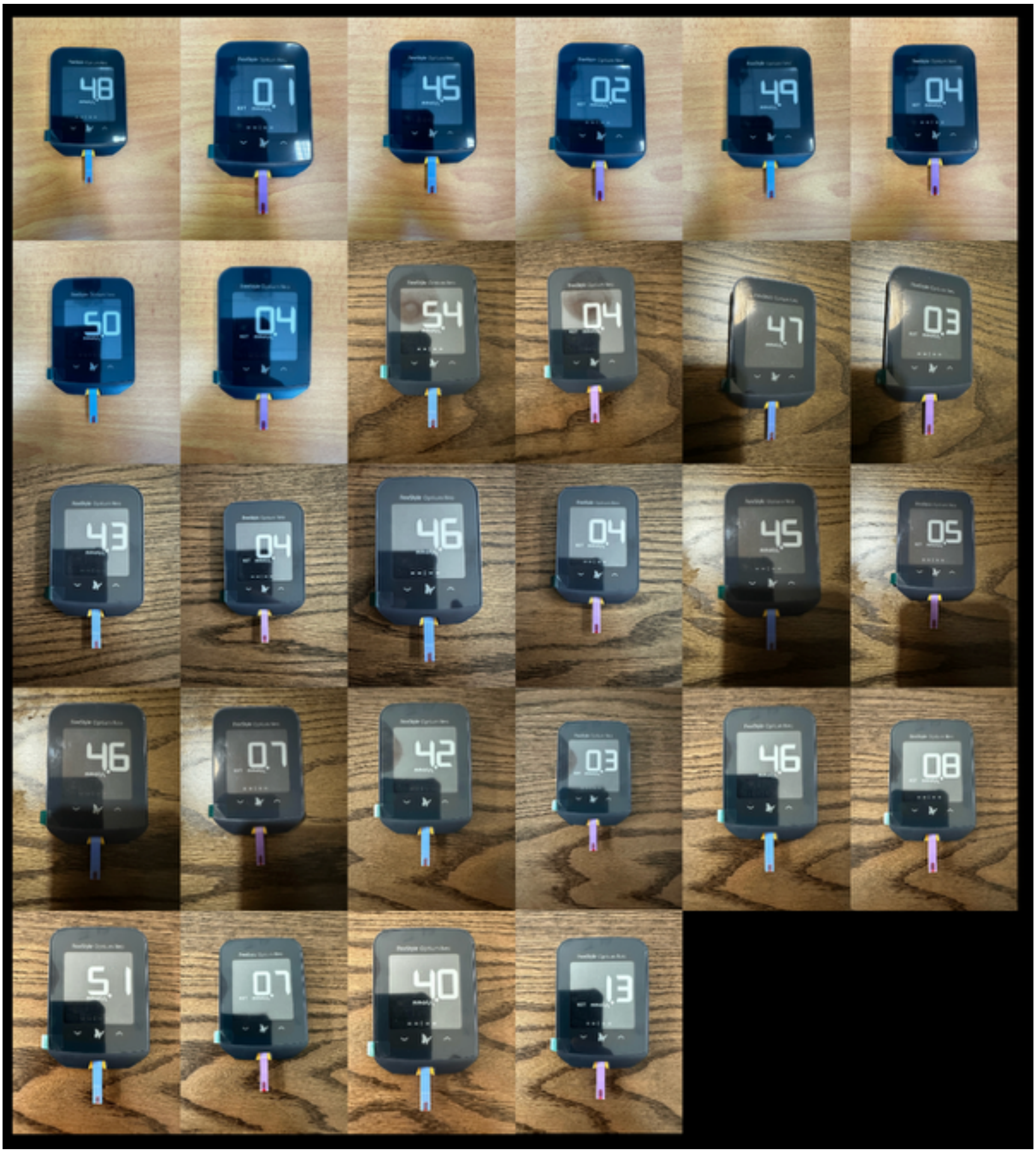

**Fig. S10.** Measured blood ketone and glucose levels for the OMAD ketosis entry regimen from Nov 19-20, 2024.

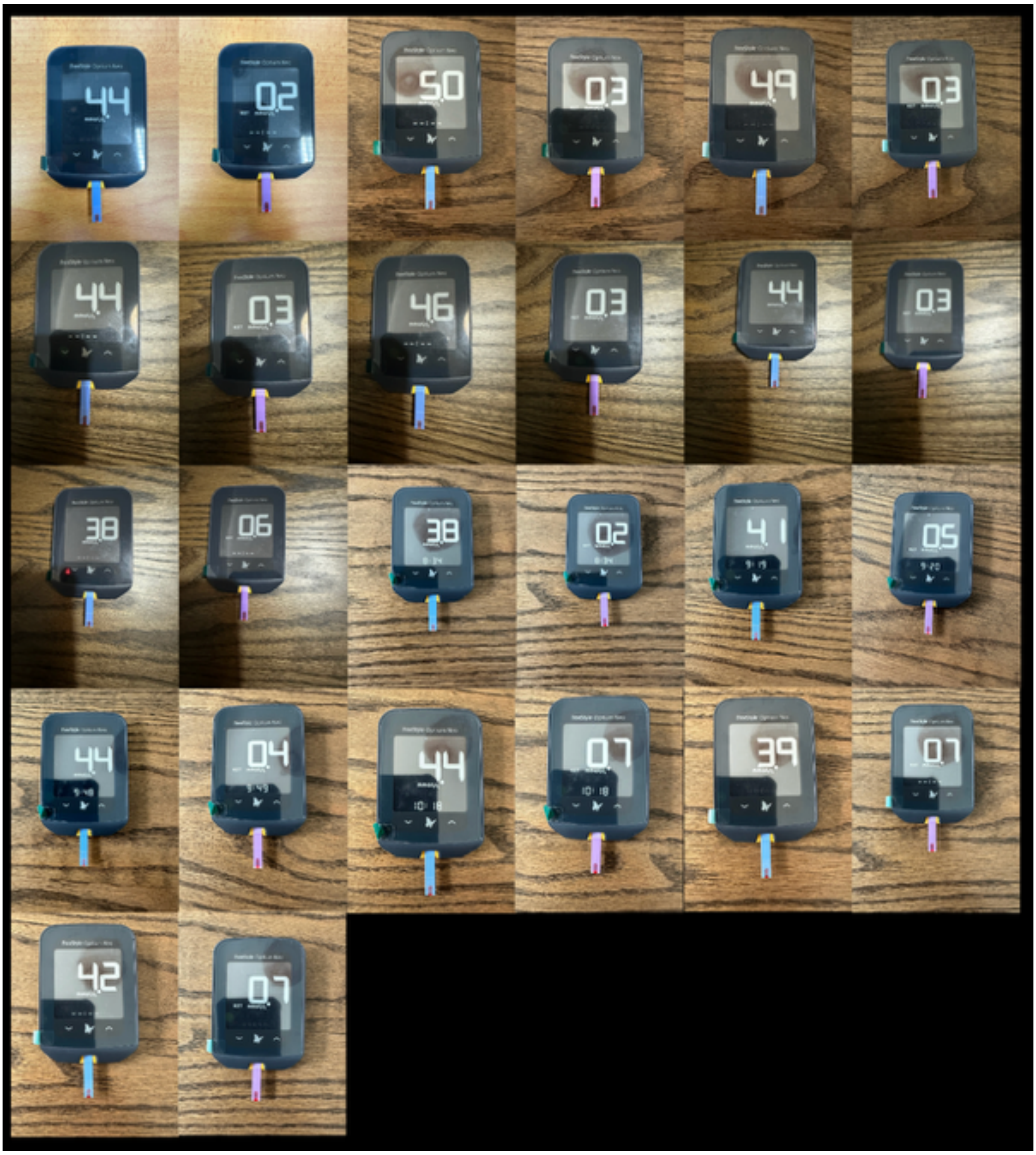

**Fig. S11.** Measured blood ketone and glucose levels for the OMAD ketosis entry regimen from Nov 26-27, 2024.

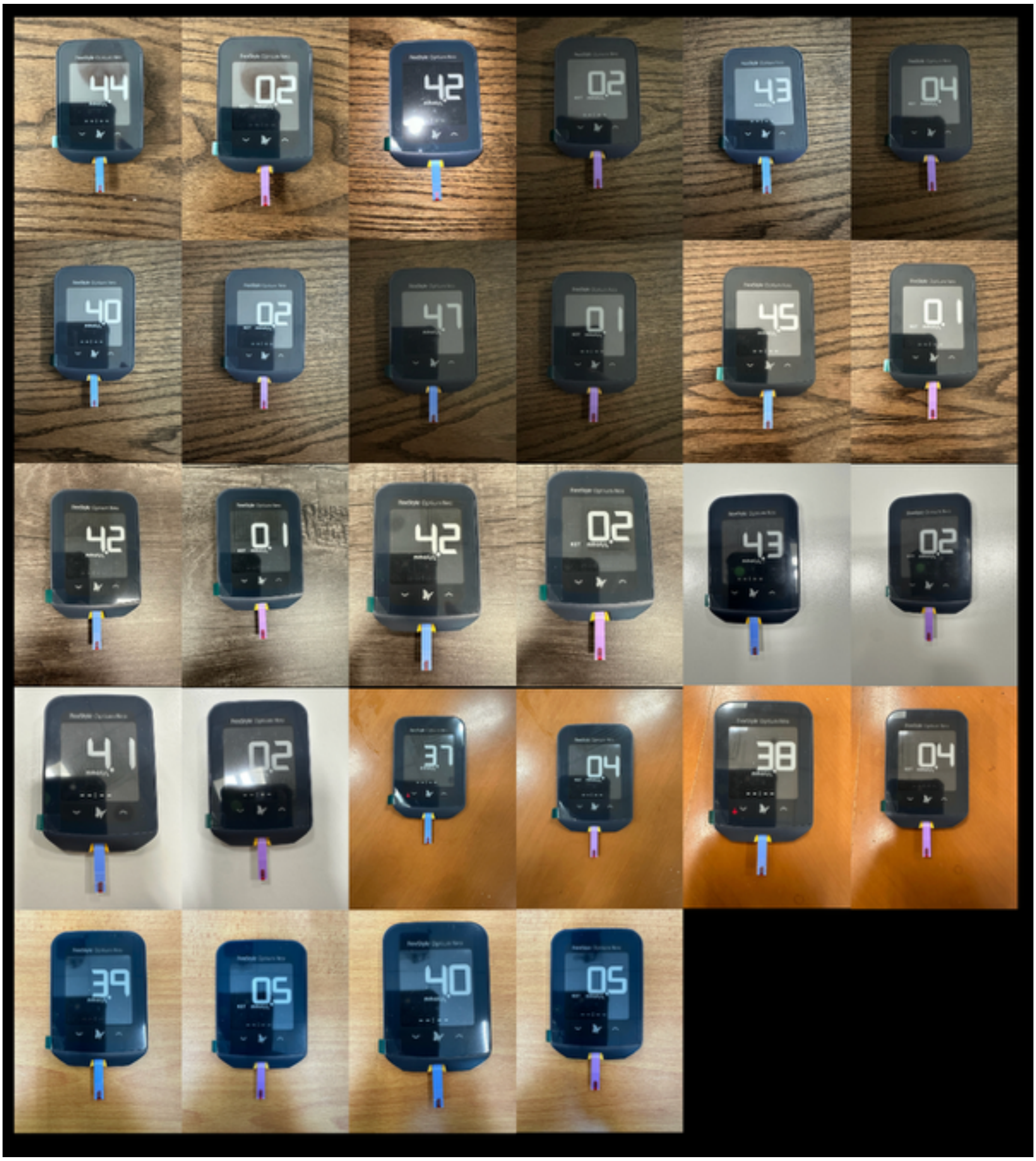

**Fig. S12.** Measured blood ketone and glucose levels for the OMAD ketosis entry regimen from Dec 9-10, 2024.

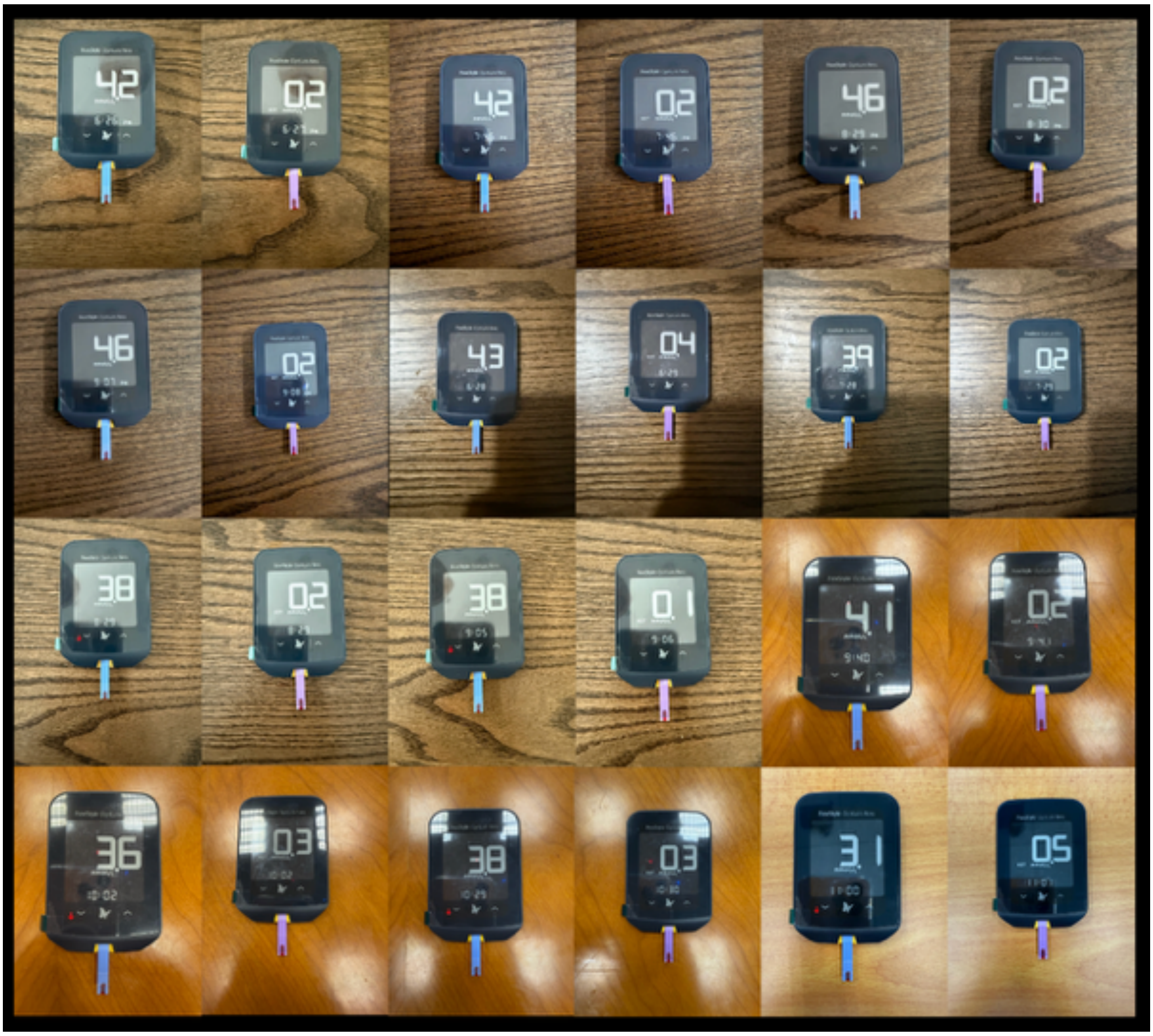

**Fig. S13.** Measured blood ketone and glucose levels for the OMAD ketosis entry regimen from Feb 3-4, 2025.

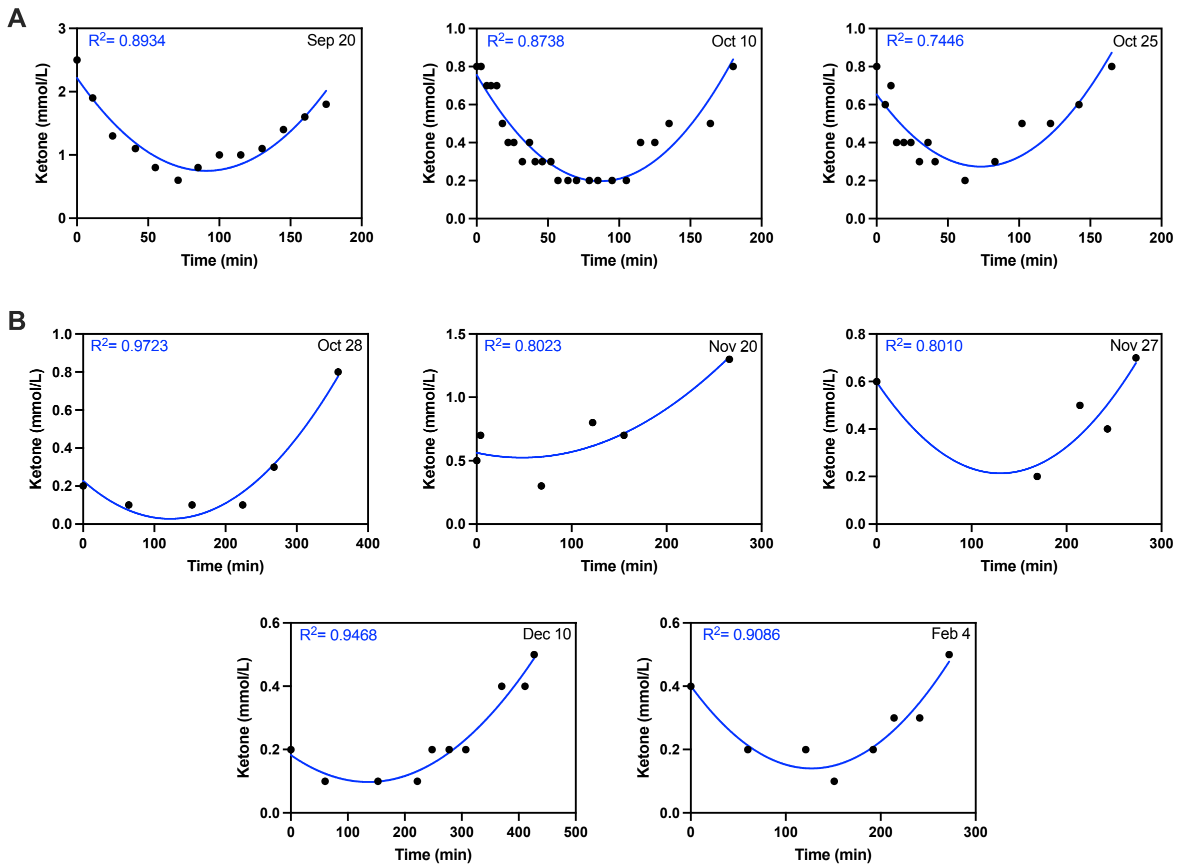

**Fig. S14.** Second-order regression analysis of ketone dynamics. **(A)** The 48-hour fasting strength training regimen. Measurements taken from exercise start to ketotic recovery were used for regression analysis. **(B)** The OMAD ketosis entry regimen. Measurements taken on day 2 of the regimen were used for regression analysis.

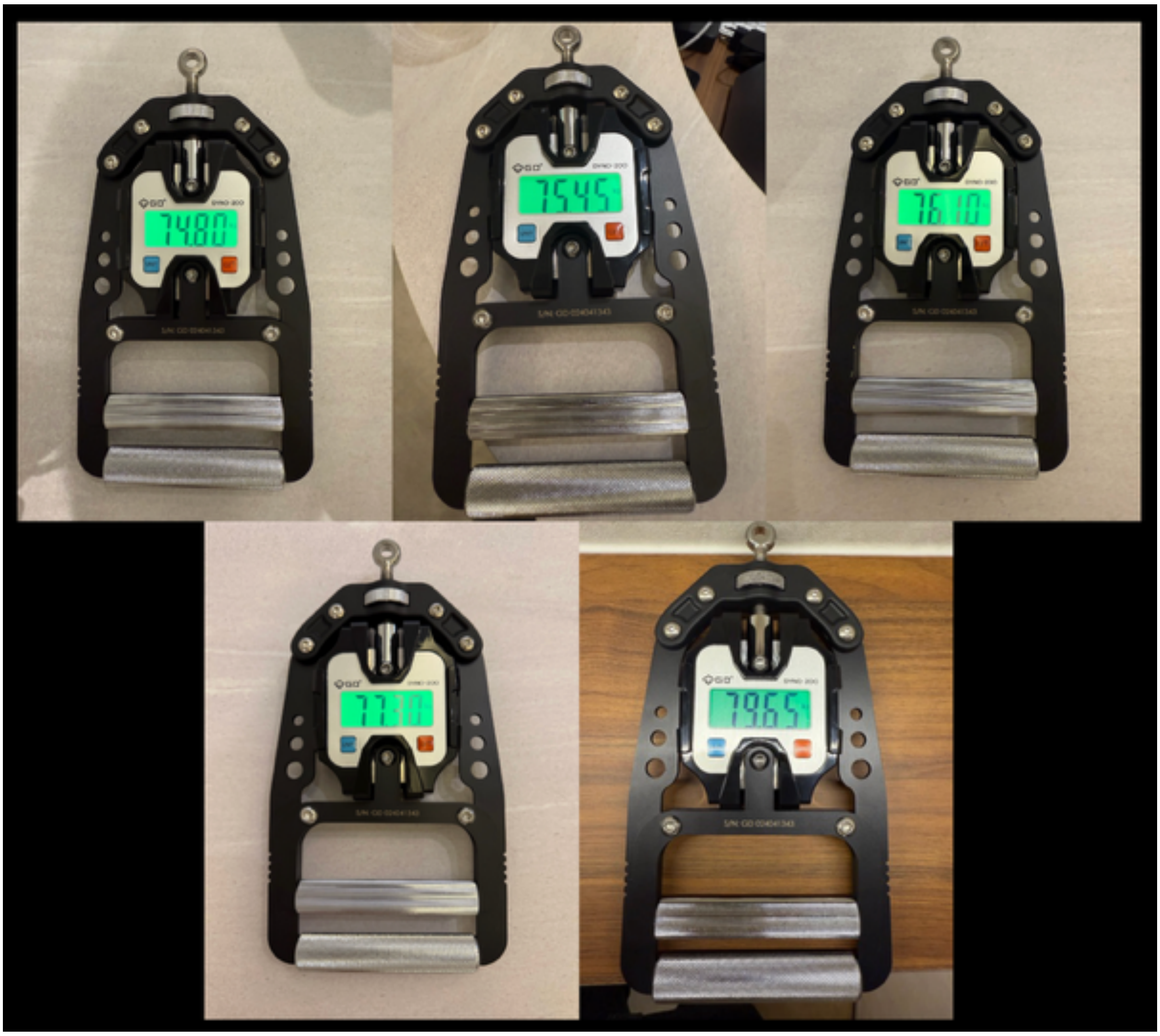

**Fig. S15.** Grip strength (dominant-hand) measured using an industrial grade grip strength meter (GD DYNO 200) on Feb 17, 18, and 28 2025.

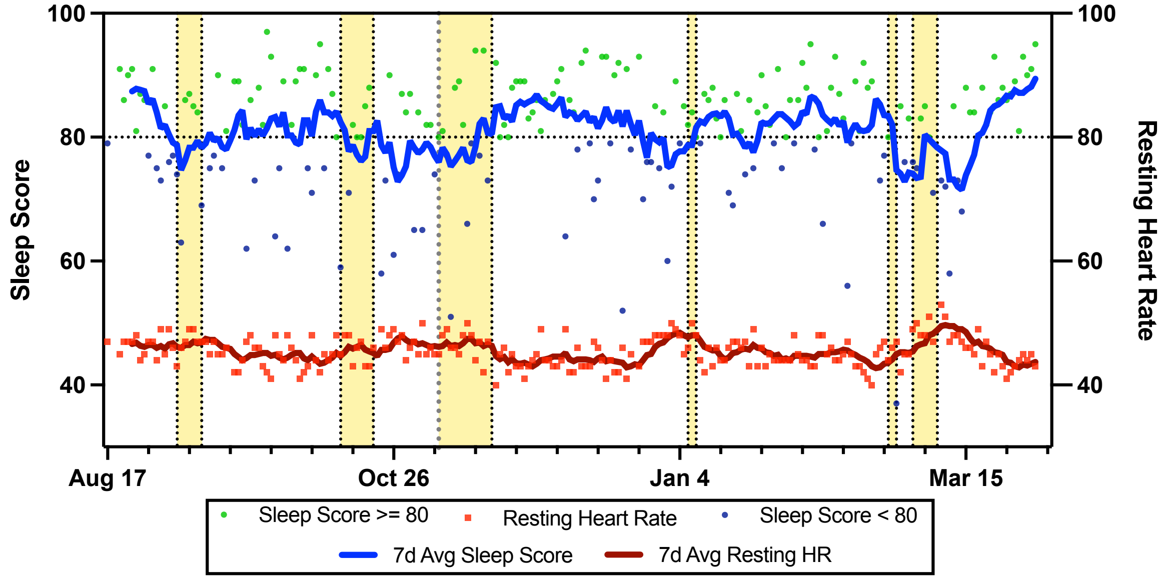

**Fig. S16.** Longitudinal tracking of Garmin sleep score compared to trends in resting heart rate (RHR). The lines indicate a 7-day rolling average of RHR (red) and sleep score (blue). Yellow shaded regions indicate periods of overseas travel. Source data can be found in Supplementary Data 1.

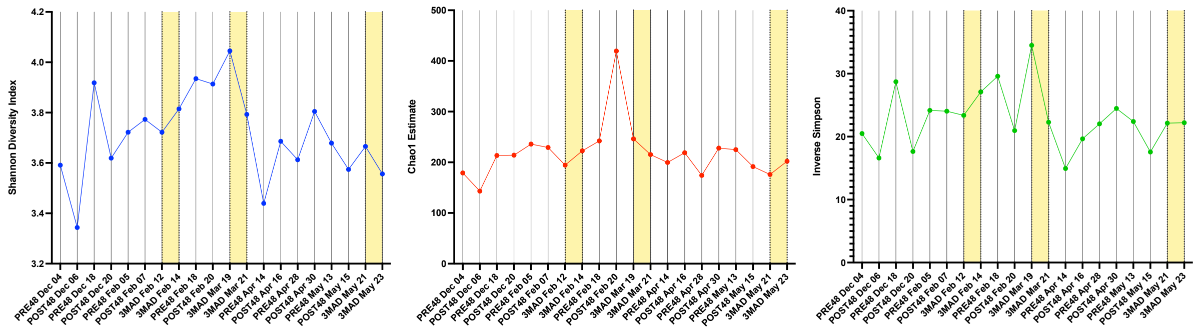

**Fig. S17.** Microbiome diversity measured using Shannon, Chao1 and Inverse Simpson over the course of the study – with PRE48 indicating samples that were taken before a 48-hour fast, POST48 indicating samples that were taken after a 48-hour fast and 3MAD indicating samples taken during the intervention period of a 3MAD week (highlighted in yellow). Source data can be found in Supplementary Data 2.

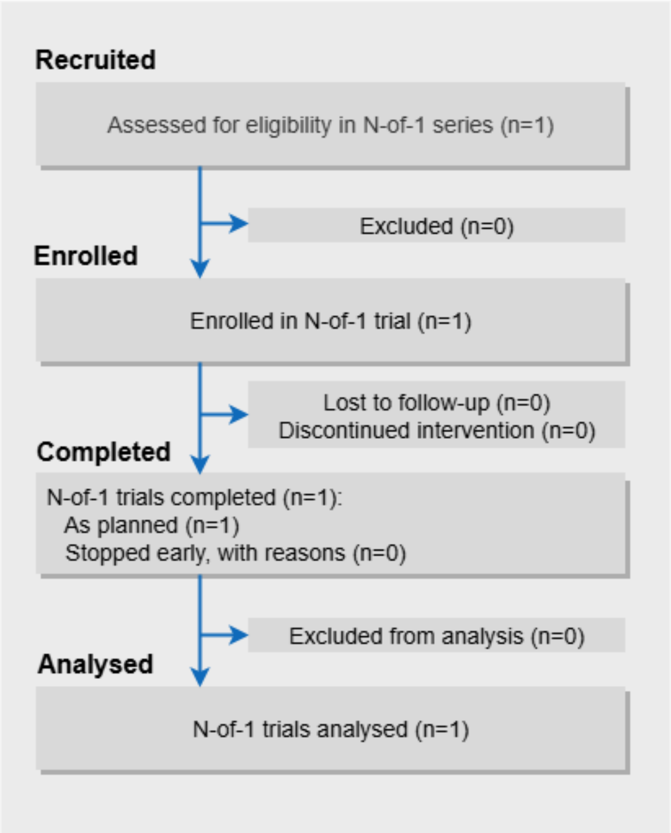

**Fig. S18**. CONSORT extension for reporting N-of-1 trials (CENT) participant flow diagram illustrating flow of participant in N-of-1 trial. There were no dropouts in this study, and the study's sole participant remained in the trial until its completion.

**Supplementary Data 1:** Source data for fitness performance, sleep performance, cardiometabolic health (biomarkers, HRV, blood pressure), and weight.

**Supplementary Data 2:** Source data for biomarker dynamics.

**Supplementary File 1:** DELTA001's dietary regimen throughout the study.

**Supplementary File 2:** Source files for GPT-enabled Healthspan Copilot.

**Supplementary File 3:** Transcripts for participant experience assessment.
