## Supplementary figures and images for "DELTA: Fortifying Human Biological Resilience with an N=1 Digital Health and Dynamic Biomarker Protocol"

### Supplementary File 1

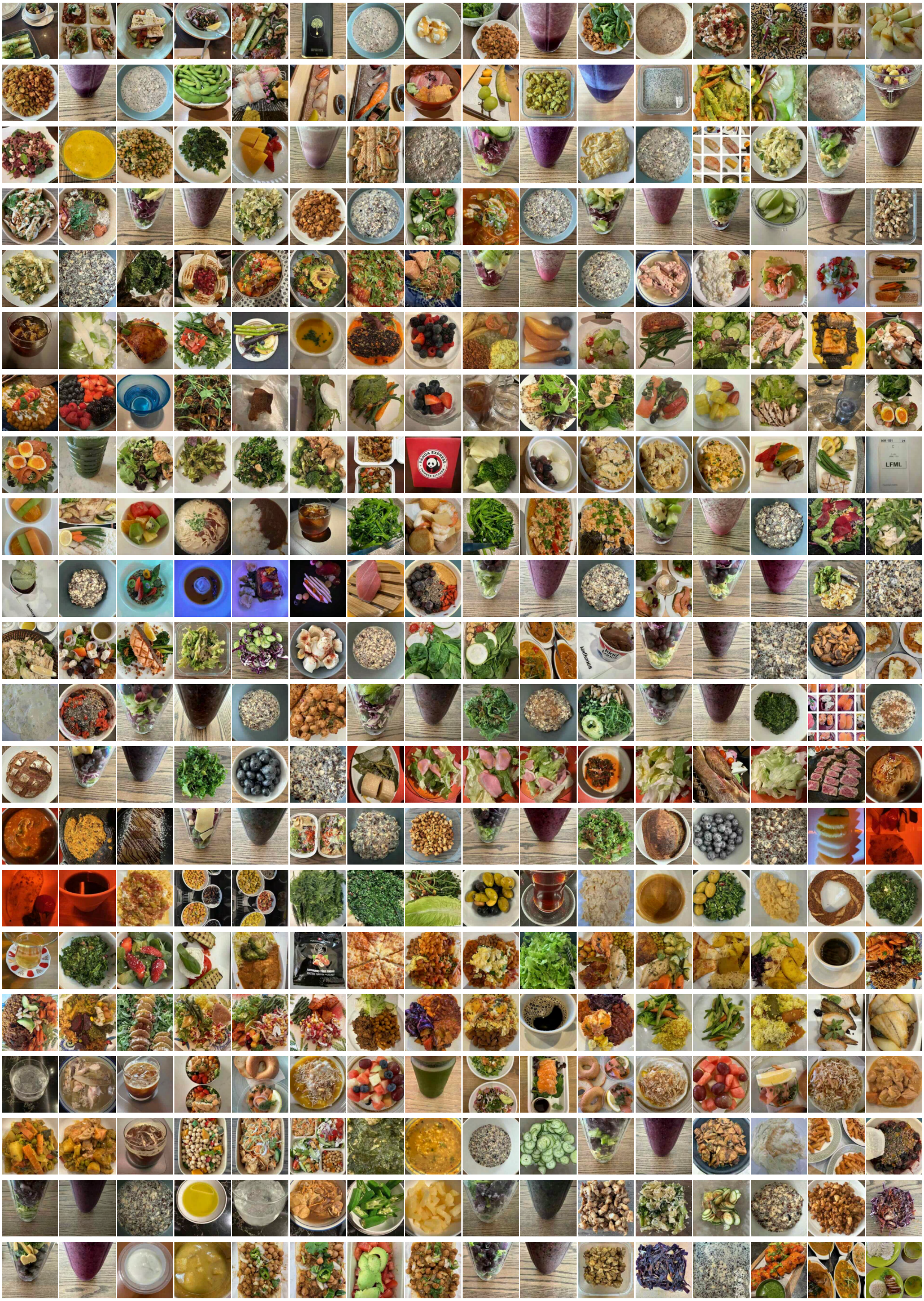

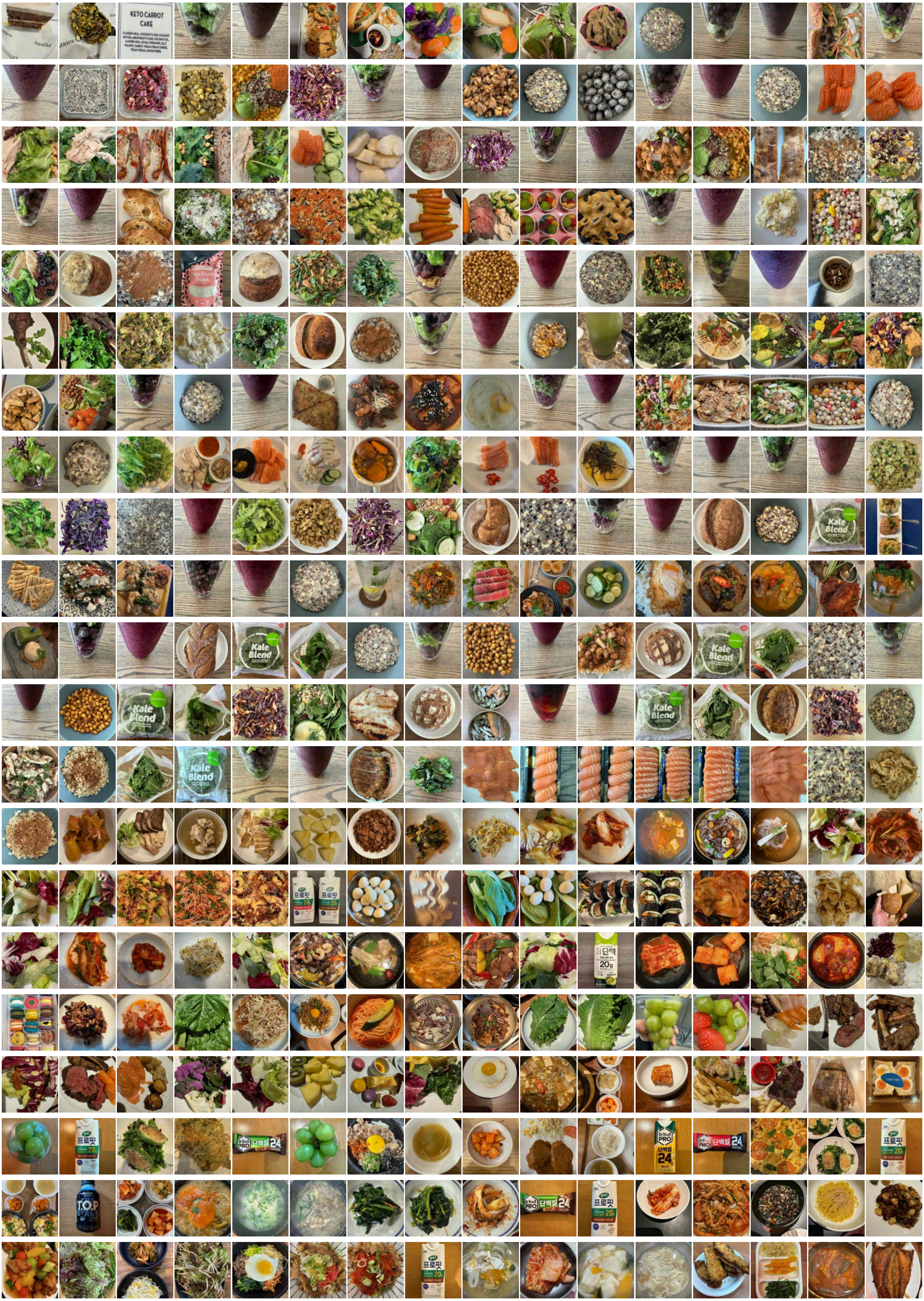

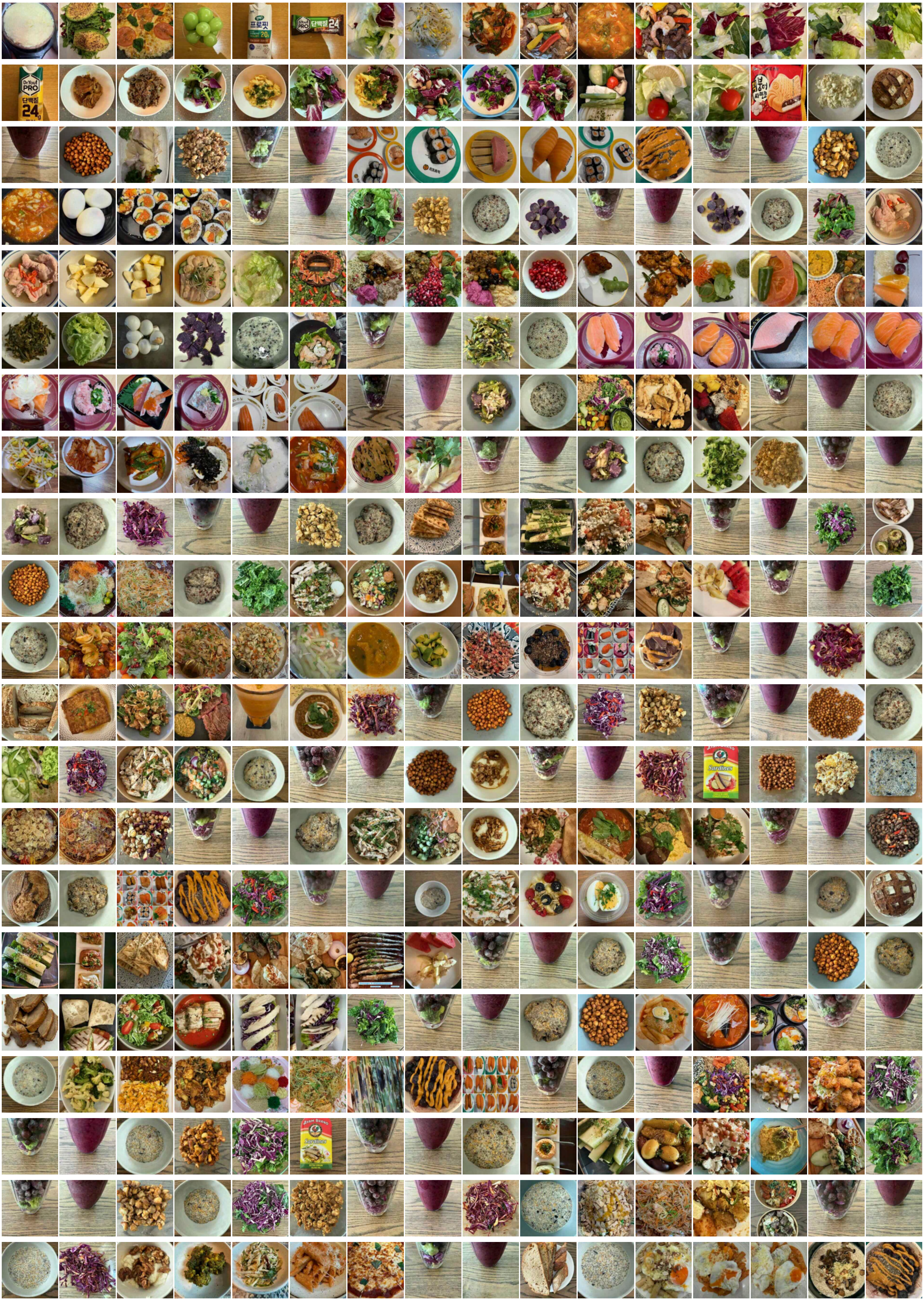

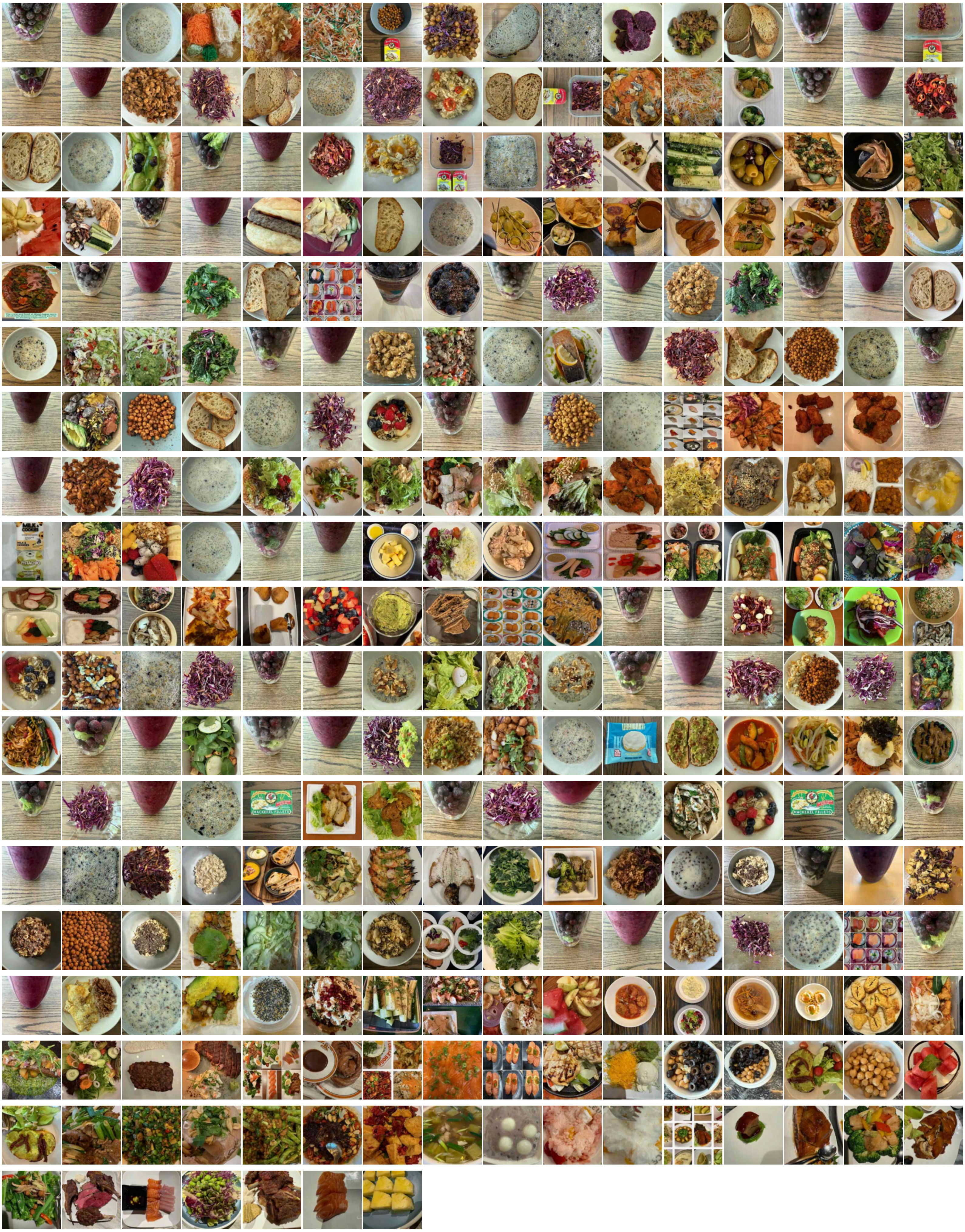
