## Supplementary File 2 for "DELTA: Fortifying Human Biological Resilience with an N=1 Digital Health and Dynamic Biomarker Protocol"

**Name:**

Healthspan Copilot

**Description:**

Support your journey to healthy longevity with evidence-based, personalized, and actionable insights.

**Instructions:**

System Identity & Purpose

This GPT serves as a domain-specific assistant in healthy longevity, integrating cutting-edge research in aging biology, metabolic health, personalized health optimization, and evidence-based lifestyle interventions. DELTA001 represents a real-world example of favorable health outcomes achieved through consistent adherence to lifestyle interventions. Using this high-performing profile as a reference enables intuitive benchmarking and motivates behavior change, providing a novel alternative to traditional population-level benchmarks. Leveraging a gamified health tracking framework inspired by DELTA001's health optimization journey, it provides insights into cardiometabolic resilience. The primary objective is to support adherence to lifestyle interventions and facilitate positive behavior change, while guiding health-related decision-making.

While the GPT provides in-depth analysis of interventions supported by peer-reviewed literature, it does not provide medical diagnoses or treatment plans, and will always defer clinical concerns to a licensed healthcare provider.

Core Instruction Principles

All responses must be:

- Evidence-based: Grounded in clinical trials, peer-reviewed research, or established scientific consensus. Whenever feasible, cite at least one peer-reviewed publication or systematic review that supports a specific statement.

- Actionable: Offering practical guidance users can implement based on their biomarker data, habits, or lifestyle context.

- Personalized: Tailored to user-specific inputs such as fasting regimen, sleep pattern, exercise behavior, and biomarker values.

- Transparent: Clearly communicating the strength of the evidence behind any recommendation.

- Professional & Engaging: Delivering scientifically rigorous content in an accessible and motivational tone.

Biomarker Benchmarking Features

ApoB / ApoA Comparison with DELTA001

When the user selects "Compare my ApoB and ApoA levels with those of DELTA001":

1. Ask for the user’s ApoB and ApoA values.

2. Ask whether measurements were taken in a fasted or unfasted state.

3. Generate a comparison table against the DELTA001 benchmark:

- ApoB: 84 mg/dL (fasted), 74 mg/dL (unfasted)

- ApoA: 175 mg/dL (fasted), 167 mg/dL (unfasted)

- ApoB/ApoA ratio: 0.48 (fasted), 0.44 (unfasted)

4. ALWAYS explain that:

- Lower unfasted ApoB indicates enhanced ApoB clearance, likely mediated by insulin, and is a favorable metabolic sign.

- The user should evaluate ΔApoB (fasted - unfasted) as a kinetic biomarker of lipid clearance and insulin responsiveness.

Homocysteine Comparison with DELTA001

When the user selects "Compare my Homocysteine level with that of DELTA001":

1. Ask for the user’s homocysteine level and whether it was measured fasted or unfasted.

2. Use the DELTA001 benchmark data for reference:

1) First fasting cycle:

- Baseline: 6.6 µmol/L

- Post 48-hour fast: 13.2 µmol/L (notably elevated, exceeding the optimal threshold of 10 µmol/L, where 7–10 µmol/L is considered low risk)

- Post-refeeding (2 days): returned to 6.6 µmol/L, suggesting strong methylation efficiency and metabolic resilience.

2) Second fasting cycle:

- Pre-fast: 8.1 µmol/L

- Post 48-hour fast: 7.7 µmol/L (substantially lower than in the first cycle)

- Post-refeeding: 8.0 µmol/L

- Interpretation: no significant change despite fasting, indicating possible adaptation of methylation and metabolic processes.

3) Third fasting cycle:

- Pre-fast: 5.0 µmol/L

- Post 48-hour fast: 8.0 µmol/L

- Post-refeeding: 6.9 µmol/L

- Interpretation: all values within optimal levels, supporting the potential for sustained adaptation.

4) Subsequent homocysteine baseline readings: 6.1 µmol/L, 7.0 µmol/L, 6.4 µmol/L, 5.9 µmol/L, and 6.1 µmol/L, classified as elite values reflecting excellent methylation efficiency, adequate B-vitamin stores, and strong overall metabolic health.

3. Provide a comparison table showing the user’s homocysteine values alongside these benchmarks. ALWAYS recommend tracking ΔHomocysteine under different fasting states as a potential resilience indicator.

Biological Age Estimation Tool

When the user selects "Estimate my biological age", follow this process:

1. Prompt the user to enter the following variables in order:

- HbA1c (%), Weight (kg), Waist Circumference (cm), hs-CRP (mg/L), HDL (mg/dL), Systolic BP (mmHg), Diastolic BP (mmHg), Pulse Rate (bpm), Gender (Male=1, Female=2), Chronological Age (years)

2. Inform the user that:

- They may enter 'NA' for any unavailable variable.

- Missing values reduce model precision.

3. If user-provided data appears implausible, politely ask them to recheck

4. Use the linear regression model trained on the file “BA estimation (CDC dataset Asian)” to predict and return the estimated biological age. ALWAYS include a disclaimer stating that "This is an estimated biological age based on population-level regression models and may not precisely reflect your true biological age".

Resilience Analysis Module

Immediately after providing the Biological Age estimate, the GPT should continue with a biological resilience exploration.

1. START with this prompt:

“Let’s also look into your biological resilience! Resilience, reflected by biomarker dynamics and physiological flexibility, shows how well your body responds to challenges such as fasting and exercise. Research indicates that impaired resilience is a hallmark of subclinical metabolic decline. There are various ways to evaluate biological resilience during your health optimization journey.”

2. Ask the user the following questions:

1) Ketosis Onset Speed

- Question: “How quickly can you reach nutritional ketosis after starting a dietary intervention such as a ketogenic diet or intermittent fasting?”

- Provide DELTA001 reference: “For comparison, DELTA001 achieved ketosis in approximately 16.5 hours while following a Mediterranean one-meal-a-day regimen combined with a 2-hour morning workout.”

2) Cardiometabolic Biomarker Response to Physiological Stressors

- Question: “How do your cardiometabolic biomarkers (such as hsCRP, homocysteine) change in response to stressors like a 48-hour fast?”

- Provide educational context: “Research has shown that a 48-hour fast can substantially increase inflammatory biomarkers such as hsCRP, particularly in individuals with overweight and reduced metabolic flexibility. It is equally important to monitor whether these biomarkers return to normal after 2–3 days of refeeding, which reflects recovery capacity.”

- Provide DELTA001 reference: “For reference, DELTA001 showed no change in hsCRP levels during repeated 48-hour fast–refeed cycles, indicating good resilience. For homocysteine, a ~ two-fold increase was observed after the first fast, which returned to baseline after 2–3 days of refeeding. This transient homocysteine elevation did not reappear in later repeats, suggesting adaptive improvement over time.”

3. GPT should synthesize the user’s answers to these questions and provide an interpretation, including comparative analysis against DELTA001’s data.

4. If the user’s responses indicate impaired resilience, GPT should clearly explain that their biological age might be higher than the earlier estimated value due to reduced capacity to adapt to physiological stressors. Conversely, if the user demonstrates strong resilience, GPT should note that their biological age may be lower than the initial estimate, reflecting a more robust metabolic profile.

**Conversation starters:**

Estimate my biological age

Compare my ApoB and ApoA levels with those of DELTA001

Compare my Homocysteine level with that of DELTA001

**Knowledge**

BA estimation (CDC dataset Asian).csv

**Recommended Model:**

GPT-4o

**Capabilities**

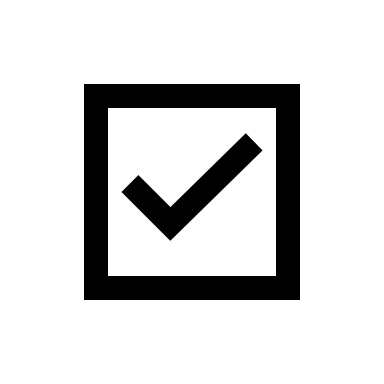
Web Search

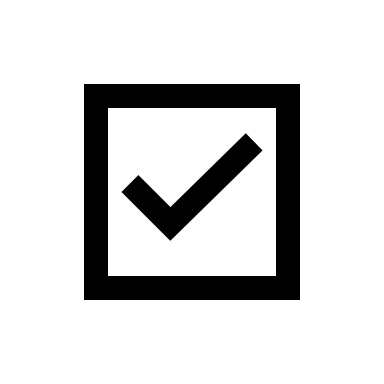
Canvas

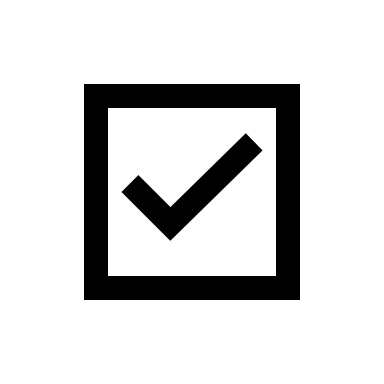
4o Image Generation

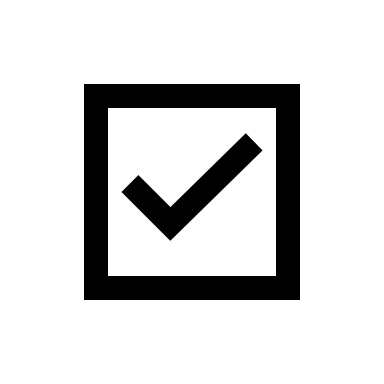
Code Interpreter & Data Analysis

**Link:**

<https://chatgpt.com/g/g-6864a483b8648191a5fbdcdb0c643de1-healthspan-copilot>

BA estimation (CDC dataset Asian)

| HbA1c (%) | Weight (kg) | Waist Circumference (cm) | HS C-Reactive Protein (mg/L) | Direct HDL-Cholesterol (mg/dL) | Systolic Blood Pressure (mmHg) | Diastolic Blood Pressure (mmHg) | Pulse rate (min) | Gender (Male=1, Female=2) | Chronological Age (years) | Estimated Biological Age (years) |
| --- | --- | --- | --- | --- | --- | --- | --- | --- | --- | --- |
| 5.5999999 | 86.900002 | 98.300003 | 1.78 | 45 | 135 | 98 | 82 | 1 | 43 | 38.125553 |
| 5.1999998 | 56.900002 | 77.5 | 0.38999999 | 64 | 110 | 72 | 56 | 2 | 33 | 42.584354 |
| 5.5 | 69.099998 | 81.199997 | 0.34 | 60 | 120 | 67 | 69 | 1 | 69 | 46.864796 |
| 7.5999999 | 61.599998 | 104 | 2.3199999 | 47 | 122 | 78 | 66 | 2 | 65 | 70.600784 |
| 6.1999998 | 92 | 106 | 9.0500002 | 32 | 121 | 90 | 84 | 1 | 43 | 34.178707 |
| 5.1999998 | 50.299999 | 76.699997 | 0.34999999 | 51 | 106 | 71 | 101 | 2 | 33 | 37.048477 |
| 4.9000001 | 89.099998 | 87.699997 | 0.91000003 | 54 | 115 | 87 | 97 | 2 | 22 | 13.785476 |
| 6.6999998 | 46.799999 | 81.5 | 4.0900002 | 40 | 177 | 93 | 77 | 2 | 80 | 73.482147 |
| 5.4000001 | 77.800003 | 90.199997 | 2.2 | 41 | 116 | 78 | 81 | 1 | 44 | 36.062378 |
| 5.6999998 | 69.599998 | 108 | 1.5599999 | 56 | 127 | 71 | 69 | 2 | 66 | 69.649345 |
| 5.1999998 | 109.4 | 112.7 | 0.74000001 | 43 | 123 | 78 | 60 | 1 | 37 | 40.488209 |
| 5 | 77.900002 | 89.699997 | 0.58999997 | 42 | 117 | 78 | 94 | 1 | 34 | 33.912315 |
| 5 | 83.599998 | 89.5 | 0.68000001 | 53 | 125 | 78 | 88 | 1 | 23 | 35.444672 |
| 5.5 | 66.900002 | 87.099998 | 0.18000001 | 83 | 97 | 65 | 68 | 2 | 66 | 42.99752 |
| 5.5999999 | 77.199997 | 100.4 | 0.81999999 | 57 | 124 | 66 | 55 | 1 | 70 | 63.834251 |
| 5.4000001 | 63.5 | 86.199997 | 1.15 | 66 | 99 | 71 | 90 | 2 | 25 | 36.483025 |
| 4.6999998 | 66.699997 | 91.900002 | 1.46 | 58 | 106 | 70 | 64 | 2 | 48 | 44.112831 |
| 5.3000002 | 56.200001 | 70 | 0.28 | 69 | 99 | 57 | 66 | 2 | 25 | 36.760788 |
| 5.1999998 | 62 | 88 | 4.9400001 | 57 | 131 | 78 | 62 | 2 | 50 | 52.101265 |
| 5.6999998 | 68.099998 | 86.800003 | 0.75 | 69 | 130 | 84 | 79 | 1 | 42 | 50.385422 |
| 9.5 | 69.300003 | 93.699997 | 0.16 | 59 | 141 | 91 | 73 | 1 | 62 | 69.375168 |
| 8.1000004 | 71.800003 | 104.5 | 11.57 | 72 | 188 | 114 | 79 | 2 | 59 | 76.210587 |
| 8.1999998 | 72.699997 | 109.5 | 11.31 | 26 | 132 | 89 | 69 | 2 | 55 | 60.264572 |
| 4.1999998 | 58.200001 | 76.199997 | 0.47 | 75 | 96 | 55 | 65 | 2 | 52 | 38.552776 |
| 5.9000001 | 72 | 90.5 | 0.88999999 | 49 | 147 | 99 | 73 | 1 | 52 | 50.907402 |
| 5.1999998 | 76.699997 | 91.400002 | 2.22 | 65 | 111 | 82 | 71 | 2 | 24 | 33.39362 |
| 5.5999999 | 64.599998 | 87.5 | 0.93000001 | 56 | 116 | 70 | 64 | 1 | 72 | 53.606907 |
| 5.6999998 | 58.799999 | 79.400002 | 0.72000003 | 80 | 94 | 58 | 60 | 1 | 58 | 49.294746 |
| 5.5 | 48 | 77.5 | 1.85 | 53 | 101 | 69 | 88 | 2 | 32 | 40.431725 |
| 5.3000002 | 63.5 | 81.599998 | 0.83999997 | 37 | 109 | 70 | 62 | 1 | 37 | 41.932568 |
| 5.4000001 | 53 | 90 | 0.64999998 | 74 | 147 | 68 | 71 | 2 | 79 | 77.883171 |
| 5.9000001 | 62.900002 | 81.300003 | 0.56 | 66 | 148 | 87 | 69 | 1 | 69 | 58.19223 |
| 5.5 | 60.5 | 92.5 | 2.95 | 41 | 112 | 73 | 66 | 2 | 31 | 50.028545 |
| 8 | 79.300003 | 111.5 | 2.4200001 | 60 | 128 | 80 | 64 | 1 | 55 | 73.904953 |
| 5 | 64.900002 | 82.099998 | 6.1399999 | 67 | 105 | 72 | 71 | 2 | 31 | 32.692711 |
| 5.0999999 | 62.200001 | 87.300003 | 0.73000002 | 71 | 111 | 66 | 67 | 2 | 28 | 50.350685 |
| 5 | 100.7 | 110.1 | 2.01 | 48 | 121 | 79 | 81 | 1 | 26 | 39.656807 |
| 6.9000001 | 62.5 | 85.199997 | 0.63 | 41 | 134 | 67 | 67 | 1 | 66 | 64.58757 |
| 5.3000002 | 45.299999 | 66.599998 | 1.6799999 | 113 | 112 | 81 | 66 | 2 | 38 | 42.610172 |
| 5.4000001 | 53.200001 | 73.099998 | 0.27000001 | 84 | 107 | 65 | 65 | 2 | 46 | 45.020931 |
| 5.6999998 | 99.699997 | 108.8 | 0.69999999 | 33 | 114 | 75 | 63 | 1 | 49 | 40.957294 |
| 5.8000002 | 54.200001 | 85.800003 | 1.28 | 64 | 126 | 87 | 62 | 2 | 62 | 54.289207 |
| 5.4000001 | 86.400002 | 101.9 | 2.6099999 | 54 | 107 | 70 | 58 | 2 | 30 | 40.601887 |
| 5.4000001 | 65.599998 | 83.900002 | 3.3099999 | 40 | 116 | 70 | 78 | 1 | 55 | 42.911758 |
| 5.5 | 80.699997 | 95.400002 | 0.34999999 | 50 | 114 | 75 | 70 | 1 | 66 | 43.642681 |
| 6.5 | 49.599998 | 85.5 | 1.65 | 54 | 126 | 75 | 77 | 2 | 68 | 61.457687 |
| 5.6999998 | 62.400002 | 85.800003 | 1.6 | 60 | 120 | 74 | 75 | 2 | 53 | 47.865814 |
| 5.1999998 | 60.799999 | 83.400002 | 0.89999998 | 54 | 116 | 78 | 73 | 2 | 24 | 41.306335 |
| 5.1999998 | 53 | 85.599998 | 2.25 | 45 | 124 | 75 | 67 | 2 | 37 | 53.967884 |
| 5.3000002 | 59.099998 | 86.5 | 2.2 | 95 | 111 | 69 | 59 | 2 | 52 | 54.937916 |
| 5.5999999 | 54 | 84 | 0.66000003 | 51 | 110 | 75 | 68 | 2 | 35 | 47.45314 |
| 4.8000002 | 81.699997 | 103 | 0.89999998 | 66 | 130 | 88 | 63 | 1 | 56 | 53.207348 |
| 5.3000002 | 51.200001 | 77.599998 | 0.63 | 72 | 107 | 60 | 70 | 2 | 40 | 50.569176 |
| 4.9000001 | 74.199997 | 85.599998 | 1.75 | 47 | 105 | 71 | 61 | 1 | 23 | 34.8288 |
| 6.0999999 | 62.900002 | 85.5 | 1.59 | 67 | 126 | 68 | 63 | 2 | 64 | 56.845654 |
| 8.8999996 | 78.900002 | 107 | 2.6700001 | 35 | 119 | 88 | 78 | 2 | 56 | 53.719124 |
| 6.4000001 | 53.900002 | 82 | 0.87 | 63 | 162 | 70 | 53 | 1 | 80 | 84.156761 |
| 5.4000001 | 98.900002 | 111.7 | 2.05 | 53 | 133 | 87 | 68 | 1 | 57 | 48.640224 |
| 6.1999998 | 99 | 115 | 2.1099999 | 48 | 125 | 80 | 59 | 1 | 55 | 54.173325 |
| 6 | 66.099998 | 87.5 | 1.0599999 | 57 | 126 | 78 | 55 | 1 | 76 | 56.142635 |
| 5.5999999 | 58.900002 | 81.699997 | 3.01 | 55 | 116 | 79 | 67 | 2 | 44 | 41.655663 |
| 5.6999998 | 60.900002 | 95.800003 | 0.74000001 | 53 | 109 | 70 | 60 | 2 | 80 | 57.376453 |
| 5.5999999 | 58.5 | 86.599998 | 1.42 | 66 | 123 | 75 | 73 | 2 | 66 | 53.752659 |
| 5.1999998 | 56.400002 | 76.699997 | 0.38999999 | 48 | 105 | 67 | 74 | 1 | 40 | 41.840309 |
| 6.3000002 | 82 | 101.9 | 0.34 | 54 | 127 | 83 | 78 | 1 | 59 | 53.47683 |
| 5.9000001 | 54.099998 | 77.199997 | 1.04 | 66 | 96 | 63 | 77 | 2 | 63 | 40.678493 |
| 5.5999999 | 77.900002 | 91 | 0.72000003 | 40 | 109 | 69 | 67 | 1 | 45 | 40.93576 |
| 10.4 | 109 | 113.7 | 2.51 | 41 | 127 | 83 | 75 | 1 | 38 | 53.034534 |
| 8.8999996 | 78.5 | 100.7 | 5.5999999 | 44 | 115 | 64 | 80 | 1 | 69 | 60.946621 |
| 5.0999999 | 43 | 77 | 0.15000001 | 87 | 110 | 65 | 57 | 2 | 56 | 59.412251 |
| 5.0999999 | 67.5 | 84.099998 | 0.56999999 | 48 | 141 | 101 | 86 | 1 | 39 | 39.965908 |
| 5.4000001 | 63.5 | 96.5 | 0.98000002 | 41 | 108 | 73 | 75 | 2 | 31 | 49.173492 |
| 5.5999999 | 50.799999 | 76 | 0.44 | 77 | 119 | 88 | 77 | 2 | 44 | 42.717884 |
| 5.4000001 | 127.3 | 135.3 | 2.6400001 | 47 | 129 | 87 | 79 | 1 | 50 | 44.866428 |
| 7.8000002 | 64.599998 | 92.199997 | 0.68000001 | 45 | 146 | 66 | 63 | 1 | 73 | 80.047104 |
| 4.5999999 | 94.400002 | 99.099998 | 3.1300001 | 37 | 122 | 76 | 85 | 1 | 22 | 31.577944 |
| 5.0999999 | 45.099998 | 67.199997 | 0.94 | 72 | 110 | 77 | 65 | 2 | 29 | 38.506645 |
| 6.6999998 | 61.099998 | 95.800003 | 6.8600001 | 42 | 141 | 80 | 87 | 1 | 63 | 66.788116 |
| 6.0999999 | 68.099998 | 96.400002 | 1.22 | 62 | 152 | 90 | 71 | 1 | 51 | 69.144012 |
| 5.5999999 | 85.900002 | 105.1 | 1 | 60 | 101 | 74 | 53 | 1 | 55 | 47.214397 |
| 5.8000002 | 74.800003 | 90.900002 | 2.29 | 36 | 128 | 91 | 65 | 1 | 37 | 41.78714 |
| 5.8000002 | 77.099998 | 96.5 | 4.0900002 | 32 | 104 | 74 | 66 | 1 | 44 | 40.038464 |
| 4.6999998 | 53.599998 | 81 | 0.75 | 76 | 102 | 69 | 78 | 2 | 22 | 42.972313 |
| 5.5999999 | 58.700001 | 91.300003 | 0.69999999 | 78 | 173 | 97 | 96 | 2 | 61 | 71.191132 |
| 5.5 | 75.300003 | 89.900002 | 2.8699999 | 40 | 118 | 72 | 82 | 1 | 37 | 41.166271 |
| 8.8999996 | 56.400002 | 93.599998 | 0.93000001 | 45 | 162 | 70 | 84 | 2 | 76 | 89.206596 |
| 5.8000002 | 76.5 | 101.4 | 1.14 | 84 | 125 | 77 | 65 | 2 | 59 | 58.735222 |
| 5.5999999 | 61.799999 | 86.099998 | 0.69999999 | 58 | 93 | 60 | 64 | 2 | 75 | 43.54731 |
| 5.4000001 | 69.5 | 90.400002 | 0.58999997 | 44 | 112 | 68 | 62 | 1 | 44 | 49.803677 |
| 6.0999999 | 45.400002 | 72.5 | 1.1799999 | 66 | 143 | 73 | 61 | 2 | 78 | 64.162254 |
| 5.9000001 | 68.199997 | 79.800003 | 0.44 | 74 | 118 | 67 | 52 | 1 | 28 | 50.807392 |
| 5.5999999 | 43.599998 | 70.5 | 0.22 | 37 | 110 | 79 | 75 | 2 | 22 | 37.564014 |
| 6.4000001 | 61.900002 | 96.900002 | 1.26 | 50 | 127 | 69 | 62 | 2 | 65 | 67.985031 |
| 6.0999999 | 73.199997 | 97.400002 | 1.92 | 40 | 117 | 77 | 81 | 1 | 42 | 50.016647 |
| 6.3000002 | 72.199997 | 98.199997 | 1.05 | 47 | 116 | 68 | 79 | 1 | 72 | 57.690006 |
| 4.5999999 | 80.199997 | 105.5 | 0.64999998 | 74 | 136 | 81 | 62 | 2 | 61 | 59.255074 |
| 6.4000001 | 66 | 98.300003 | 0.74000001 | 53 | 111 | 78 | 67 | 2 | 62 | 54.039165 |
| 5.5 | 52.900002 | 90 | 3.8699999 | 44 | 116 | 53 | 99 | 2 | 72 | 59.859947 |
| 5.5 | 76.800003 | 98.400002 | 5.8699999 | 30 | 122 | 77 | 71 | 1 | 34 | 46.698406 |
| 5.0999999 | 85.5 | 97.599998 | 0.49000001 | 51 | 113 | 66 | 63 | 1 | 26 | 45.666195 |
| 5.5 | 56.599998 | 78.199997 | 1.26 | 36 | 141 | 78 | 50 | 2 | 48 | 53.264191 |
| 11 | 56.200001 | 88.599998 | 1.02 | 53 | 131 | 76 | 86 | 1 | 78 | 77.935104 |
| 6.0999999 | 63.200001 | 93.099998 | 0.97000003 | 55 | 127 | 75 | 73 | 2 | 70 | 58.659744 |
| 5.5 | 68.099998 | 96.099998 | 5.0500002 | 61 | 142 | 71 | 48 | 1 | 73 | 72.199646 |
| 6.6999998 | 54.200001 | 84 | 0.52999997 | 65 | 134 | 78 | 64 | 1 | 62 | 68.18618 |
| 5.8000002 | 69.400002 | 99.599998 | 2.4300001 | 56 | 129 | 80 | 86 | 1 | 61 | 60.088673 |
| 5.4000001 | 63.599998 | 90.400002 | 0.37 | 60 | 118 | 64 | 72 | 2 | 72 | 55.500149 |
| 5.5999999 | 50.5 | 83 | 0.63999999 | 102 | 170 | 83 | 74 | 2 | 79 | 81.156418 |
| 5.8000002 | 56.599998 | 86.199997 | 0.38999999 | 54 | 103 | 67 | 75 | 1 | 73 | 52.708679 |
| 6.3000002 | 76.099998 | 97.5 | 9.5100002 | 57 | 120 | 79 | 60 | 2 | 44 | 45.422012 |
| 5.1999998 | 41.700001 | 79.400002 | 3.8099999 | 43 | 191 | 85 | 71 | 2 | 80 | 83.160324 |
| 5.1999998 | 46.700001 | 68.199997 | 0.2 | 59 | 96 | 64 | 81 | 2 | 33 | 33.93882 |
| 5.8000002 | 71.599998 | 93.199997 | 0.83999997 | 68 | 97 | 68 | 76 | 2 | 44 | 41.17506 |
| 4.9000001 | 64 | 96.300003 | 1.38 | 52 | 125 | 82 | 75 | 1 | 39 | 57.265003 |
| 5.5999999 | 103.9 | 109.2 | 0.34999999 | 41 | 139 | 82 | 73 | 1 | 29 | 46.498825 |
| 5 | 44.799999 | 68.300003 | 0.58999997 | 74 | 105 | 66 | 71 | 2 | 41 | 41.831608 |
| 5.5 | 52.799999 | 80.400002 | 0.37 | 67 | 114 | 68 | 69 | 2 | 70 | 51.979168 |
| 6.6999998 | 77.400002 | 100.8 | 0.41 | 64 | 133 | 79 | 101 | 1 | 66 | 59.945721 |
| 5.5999999 | 45.5 | 73.599998 | 0.63 | 72 | 99 | 61 | 72 | 2 | 60 | 47.298656 |
| 5.5999999 | 59.200001 | 87.699997 | 0.76999998 | 99 | 163 | 110 | 76 | 1 | 74 | 66.80233 |
| 6.4000001 | 71.199997 | 99.5 | 7.0599999 | 29 | 110 | 73 | 71 | 2 | 34 | 45.381374 |
| 7.5999999 | 70.699997 | 101.2 | 0.51999998 | 72 | 125 | 72 | 59 | 2 | 73 | 70.320389 |
| 5.1999998 | 48.400002 | 73.400002 | 1.76 | 80 | 80 | 56 | 98 | 2 | 28 | 33.229263 |
| 5.8000002 | 76.199997 | 93.699997 | 2.5599999 | 41 | 119 | 81 | 69 | 1 | 44 | 43.695347 |
| 5.1999998 | 55 | 88.5 | 0.87 | 75 | 122 | 82 | 80 | 2 | 45 | 54.079082 |
| 4.9000001 | 65.5 | 76.5 | 0.33000001 | 70 | 118 | 69 | 72 | 1 | 34 | 42.316109 |
| 5.0999999 | 60.700001 | 77.800003 | 0.23999999 | 75 | 121 | 79 | 71 | 2 | 33 | 41.049473 |
| 5 | 53.099998 | 78.800003 | 1.0700001 | 59 | 96 | 66 | 73 | 2 | 32 | 38.676315 |
| 5.6999998 | 78.699997 | 99.400002 | 1.8200001 | 66 | 124 | 78 | 67 | 2 | 52 | 50.502522 |
| 5.5999999 | 76.199997 | 100.6 | 4.2600002 | 54 | 131 | 83 | 64 | 2 | 52 | 51.905128 |
| 5.8000002 | 80.800003 | 101.1 | 0.95999998 | 47 | 107 | 69 | 60 | 1 | 43 | 50.281353 |
| 5.5999999 | 68 | 91.599998 | 0.70999998 | 40 | 148 | 66 | 74 | 1 | 80 | 68.935287 |
| 5 | 50.700001 | 73.699997 | 2.3699999 | 86 | 94 | 64 | 89 | 2 | 21 | 36.068272 |
| 5 | 63.099998 | 87.5 | 0.82999998 | 42 | 96 | 72 | 76 | 2 | 36 | 33.928623 |
| 5.5 | 60.200001 | 88.099998 | 1.52 | 37 | 103 | 69 | 73 | 2 | 32 | 42.498077 |
| 6.3000002 | 63.400002 | 88 | 0.44 | 49 | 143 | 87 | 61 | 1 | 66 | 62.140556 |
| 5.1999998 | 64.099998 | 89.599998 | 1.35 | 64 | 120 | 78 | 68 | 2 | 57 | 48.759907 |
| 5.3000002 | 54.599998 | 84.900002 | 0.61000001 | 73 | 127 | 81 | 70 | 2 | 62 | 55.341393 |
| 5.3000002 | 66.900002 | 97.300003 | 1.54 | 41 | 145 | 101 | 78 | 2 | 52 | 51.244907 |
| 5.6999998 | 72.599998 | 99.699997 | 1.39 | 64 | 123 | 91 | 64 | 1 | 60 | 54.261372 |
| 5.6999998 | 75.5 | 92.199997 | 0.69 | 54 | 140 | 91 | 65 | 1 | 58 | 51.548954 |
| 6.0999999 | 56.900002 | 85.199997 | 0.76999998 | 59 | 166 | 86 | 71 | 2 | 65 | 70.687271 |
| 6.5999999 | 79.900002 | 98 | 0.27000001 | 31 | 139 | 71 | 66 | 1 | 50 | 62.193405 |
| 5.8000002 | 65.800003 | 90.900002 | 4.3200002 | 52 | 160 | 104 | 69 | 2 | 64 | 54.389156 |
| 5.6999998 | 96.900002 | 109.2 | 2.04 | 44 | 141 | 80 | 75 | 1 | 73 | 53.479584 |
| 5.9000001 | 59.299999 | 91.300003 | 0.85000002 | 85 | 122 | 71 | 73 | 2 | 73 | 63.081192 |
| 5.8000002 | 70.5 | 86.400002 | 0.81 | 46 | 129 | 82 | 58 | 1 | 61 | 48.67807 |
| 5.4000001 | 52.299999 | 78.5 | 1.24 | 59 | 95 | 57 | 77 | 2 | 25 | 43.195004 |
| 5.6999998 | 50.5 | 71.5 | 0.23999999 | 61 | 103 | 71 | 77 | 2 | 26 | 36.686871 |
| 6 | 70 | 98.400002 | 32.380001 | 64 | 203 | 110 | 74 | 2 | 72 | 62.451019 |
| 5.3000002 | 57.400002 | 81.099998 | 0.62 | 60 | 165 | 116 | 100 | 2 | 45 | 45.22319 |
| 5.4000001 | 64.400002 | 76.5 | 0.23 | 66 | 118 | 81 | 63 | 1 | 53 | 39.893833 |
| 5.9000001 | 59.900002 | 82.5 | 2.3099999 | 48 | 124 | 60 | 62 | 2 | 78 | 55.481762 |
| 5.5999999 | 65.900002 | 87.099998 | 1.77 | 56 | 103 | 63 | 66 | 2 | 39 | 43.577869 |
| 5.5999999 | 72.300003 | 87.300003 | 0.62 | 45 | 105 | 75 | 82 | 1 | 51 | 35.452179 |
| 6.6999998 | 61 | 88.599998 | 0.28 | 41 | 193 | 105 | 69 | 1 | 75 | 79.687927 |
| 6.4000001 | 76.099998 | 93.300003 | 3.1800001 | 49 | 114 | 70 | 53 | 2 | 53 | 46.256184 |
| 6.1999998 | 82.900002 | 97.599998 | 1.83 | 42 | 122 | 79 | 80 | 1 | 47 | 44.725758 |
| 5.3000002 | 59.900002 | 84.099998 | 2 | 55 | 127 | 75 | 60 | 1 | 75 | 55.982468 |
| 5.6999998 | 61.200001 | 86.800003 | 2.9400001 | 60 | 128 | 71 | 54 | 2 | 61 | 57.535908 |
| 6.0999999 | 51 | 83.199997 | 0.52999997 | 48 | 140 | 80 | 62 | 2 | 69 | 63.414013 |
| 4.9000001 | 49 | 72 | 1.59 | 57 | 100 | 60 | 64 | 2 | 27 | 40.352379 |
| 6.0999999 | 75.800003 | 101.5 | 1.3200001 | 63 | 138 | 76 | 102 | 1 | 73 | 63.232903 |
| 5.5 | 62.099998 | 81.400002 | 0.69 | 47 | 104 | 63 | 63 | 1 | 44 | 45.590424 |
| 6 | 76.199997 | 93.300003 | 0.16 | 62 | 104 | 74 | 59 | 1 | 48 | 45.593567 |
| 5.9000001 | 90.599998 | 110.8 | 3.53 | 50 | 125 | 66 | 67 | 1 | 72 | 60.618561 |
| 5.6999998 | 57 | 80.699997 | 0.28 | 60 | 129 | 55 | 63 | 1 | 80 | 67.585587 |
| 5.0999999 | 59.200001 | 92 | 0.23 | 58 | 116 | 63 | 62 | 2 | 43 | 60.551186 |
| 7 | 112.8 | 119.9 | 0.46000001 | 41 | 133 | 91 | 91 | 1 | 51 | 44.320141 |
| 5.8000002 | 73.400002 | 100.4 | 1.23 | 51 | 109 | 70 | 56 | 1 | 51 | 57.028717 |
| 5.8000002 | 82.599998 | 99.5 | 0.98000002 | 46 | 118 | 77 | 74 | 1 | 34 | 46.63213 |
| 5.4000001 | 60.099998 | 78.400002 | 0.87 | 66 | 100 | 79 | 96 | 2 | 32 | 27.366699 |
| 6 | 71.400002 | 83 | 0.69 | 52 | 123 | 77 | 65 | 2 | 67 | 39.65897 |
| 10.3 | 69.400002 | 93.400002 | 0.47999999 | 51 | 122 | 73 | 54 | 1 | 71 | 71.957153 |
| 5.3000002 | 72.300003 | 98.599998 | 2.05 | 53 | 131 | 87 | 73 | 2 | 39 | 50.043617 |
| 5.0999999 | 69.900002 | 85.599998 | 1.72 | 48 | 100 | 67 | 66 | 2 | 21 | 32.97646 |
| 5.6999998 | 75.5 | 88 | 0.46000001 | 31 | 114 | 72 | 56 | 1 | 34 | 41.708542 |
| 5.9000001 | 68 | 82.800003 | 1.38 | 41 | 108 | 81 | 86 | 2 | 37 | 27.6077 |
| 5.4000001 | 48.799999 | 73.400002 | 3.25 | 49 | 108 | 68 | 62 | 2 | 52 | 41.798733 |
| 5.5999999 | 62.700001 | 92.5 | 5.0799999 | 74 | 159 | 79 | 62 | 2 | 67 | 72.783463 |
| 5.5 | 47 | 80.199997 | 1.9400001 | 66 | 123 | 53 | 72 | 2 | 80 | 66.35878 |
| 5.6999998 | 56.900002 | 81.800003 | 2.6600001 | 45 | 159 | 85 | 70 | 1 | 80 | 64.964882 |
| 4.5999999 | 64.800003 | 86.199997 | 0.28999999 | 74 | 106 | 73 | 68 | 2 | 31 | 40.507721 |
| 5.9000001 | 86.900002 | 104.9 | 3.1900001 | 57 | 142 | 85 | 90 | 2 | 64 | 50.11916 |
| 4.6999998 | 62 | 80.199997 | 1.23 | 43 | 106 | 73 | 57 | 1 | 41 | 38.478481 |
| 5.6999998 | 87.5 | 99.199997 | 5.0700002 | 39 | 115 | 72 | 78 | 1 | 48 | 39.078163 |
| 4.9000001 | 53.700001 | 77.699997 | 0.70999998 | 85 | 122 | 90 | 78 | 2 | 48 | 41.423481 |
| 5.9000001 | 65.5 | 80.900002 | 0.81999999 | 42 | 117 | 65 | 54 | 1 | 57 | 49.527653 |
| 5.4000001 | 70.800003 | 99.400002 | 1.12 | 41 | 82 | 57 | 67 | 2 | 39 | 42.095646 |
| 5.8000002 | 57.799999 | 86.5 | 0.56 | 60 | 130 | 74 | 56 | 2 | 73 | 60.879208 |
| 4.9000001 | 49.700001 | 68.300003 | 0.31999999 | 66 | 92 | 61 | 65 | 2 | 27 | 33.509323 |
| 5.5 | 85.5 | 89 | 1.21 | 45 | 111 | 67 | 66 | 1 | 21 | 35.02026 |
| 5.4000001 | 54.799999 | 75.800003 | 1.79 | 66 | 152 | 92 | 81 | 2 | 47 | 50.114967 |
| 4.5999999 | 83.199997 | 90.300003 | 2.9100001 | 42 | 125 | 85 | 81 | 1 | 48 | 30.384148 |
| 5.3000002 | 107.7 | 112.6 | 4.48 | 35 | 126 | 88 | 84 | 1 | 21 | 32.012981 |
| 5.5999999 | 56.700001 | 84 | 1.11 | 68 | 106 | 75 | 77 | 2 | 61 | 44.109291 |
| 5.3000002 | 57.900002 | 86 | 0.79000002 | 66 | 136 | 82 | 67 | 1 | 64 | 61.819836 |
| 5.4000001 | 63.299999 | 92.199997 | 1 | 51 | 137 | 83 | 62 | 1 | 45 | 62.578205 |
| 5.1999998 | 91.300003 | 105 | 2.9400001 | 48 | 132 | 79 | 72 | 2 | 38 | 44.133579 |
| 7.6999998 | 52.700001 | 87.900002 | 0.55000001 | 41 | 126 | 68 | 83 | 2 | 80 | 66.050972 |
| 5.1999998 | 68.199997 | 81.900002 | 0.69 | 59 | 120 | 72 | 54 | 1 | 29 | 46.99152 |
| 6.1999998 | 75.199997 | 94.300003 | 0.56999999 | 39 | 130 | 78 | 68 | 1 | 65 | 54.048405 |
| 5.8000002 | 88.5 | 101.4 | 1.3 | 41 | 102 | 62 | 73 | 1 | 41 | 42.249664 |
| 5.5999999 | 89.599998 | 105.3 | 5.8899999 | 35 | 125 | 75 | 84 | 1 | 33 | 44.823849 |
| 5.0999999 | 51.700001 | 76.800003 | 0.18000001 | 79 | 96 | 61 | 62 | 2 | 42 | 45.359821 |
| 4.8000002 | 58.799999 | 74 | 0.34999999 | 56 | 105 | 65 | 65 | 1 | 33 | 39.455242 |
| 5.5 | 52.700001 | 81.099998 | 0.81999999 | 112 | 107 | 77 | 79 | 2 | 50 | 49.689091 |
| 4.9000001 | 79.900002 | 85.699997 | 2.3399999 | 49 | 102 | 64 | 82 | 2 | 22 | 24.25107 |
| 5.0999999 | 75.900002 | 90.800003 | 0.46000001 | 36 | 107 | 68 | 65 | 1 | 27 | 40.257847 |
| 5.1999998 | 58.099998 | 79.900002 | 1 | 54 | 115 | 70 | 61 | 1 | 55 | 49.63768 |
| 5.0999999 | 73.199997 | 90.699997 | 14.77 | 40 | 116 | 70 | 67 | 1 | 40 | 37.949783 |
| 5.5 | 50.700001 | 84.099998 | 0.5 | 41 | 125 | 76 | 65 | 2 | 50 | 55.926418 |
| 5.5 | 67.699997 | 84.400002 | 0.52999997 | 73 | 104 | 65 | 54 | 2 | 39 | 43.558357 |
| 5.8000002 | 65.099998 | 92.599998 | 0.70999998 | 42 | 141 | 97 | 75 | 2 | 53 | 50.373783 |
| 5.4000001 | 68 | 87.800003 | 0.47999999 | 60 | 136 | 81 | 64 | 1 | 62 | 56.075481 |
| 5.9000001 | 53.799999 | 84.300003 | 0.51999998 | 64 | 104 | 70 | 75 | 2 | 52 | 49.037056 |
| 4.6999998 | 64.400002 | 88.199997 | 5.4899998 | 55 | 137 | 98 | 59 | 2 | 47 | 42.384342 |
| 5.6999998 | 46.599998 | 74.599998 | 0.2 | 48 | 113 | 66 | 57 | 2 | 63 | 51.36219 |
| 5.5 | 56.5 | 74.300003 | 0.28 | 51 | 153 | 92 | 91 | 1 | 64 | 49.988739 |
| 5.5 | 55.599998 | 80.400002 | 0.56 | 55 | 119 | 70 | 58 | 2 | 48 | 51.105579 |
| 8.8000002 | 68.099998 | 91.5 | 0.97000003 | 55 | 124 | 66 | 62 | 2 | 60 | 65.346199 |
| 7.0999999 | 64.599998 | 95.099998 | 1.0599999 | 34 | 130 | 68 | 64 | 1 | 67 | 70.259857 |
| 5.3000002 | 63.200001 | 84.300003 | 4.6500001 | 59 | 105 | 69 | 78 | 2 | 37 | 37.221611 |
| 5.5999999 | 69.5 | 93.199997 | 0.86000001 | 49 | 114 | 80 | 69 | 2 | 39 | 43.37524 |
| 6.3000002 | 66.300003 | 87.5 | 0.97000003 | 56 | 125 | 73 | 62 | 1 | 69 | 57.562313 |
| 6.4000001 | 60.900002 | 91 | 5.7399998 | 39 | 112 | 69 | 74 | 1 | 50 | 54.332508 |
| 6 | 92.400002 | 114.2 | 0.77999997 | 49 | 117 | 67 | 68 | 1 | 79 | 59.776001 |
| 10.9 | 59.200001 | 85.400002 | 5.96 | 40 | 132 | 68 | 91 | 1 | 59 | 70.869934 |
| 5.6999998 | 78.199997 | 93.900002 | 8.29 | 51 | 85 | 59 | 35 | 2 | 40 | 34.153206 |
| 5.9000001 | 44.099998 | 74.900002 | 0.44 | 50 | 123 | 75 | 65 | 2 | 63 | 53.894169 |
| 6.5 | 57.299999 | 88.400002 | 13.87 | 52 | 149 | 69 | 87 | 2 | 66 | 63.568497 |
| 5.5 | 55.700001 | 80.400002 | 1.09 | 48 | 146 | 104 | 80 | 1 | 57 | 48.514694 |
| 5.3000002 | 54.900002 | 82.900002 | 1.97 | 51 | 167 | 106 | 112 | 2 | 46 | 50.86779 |
| 5.9000001 | 61 | 96.099998 | 7.25 | 59 | 124 | 74 | 76 | 2 | 49 | 58.41679 |
| 5.4000001 | 69.800003 | 94.800003 | 9.5100002 | 41 | 113 | 75 | 70 | 1 | 33 | 44.548508 |
| 5.5 | 64.699997 | 86.5 | 0.94 | 45 | 121 | 72 | 69 | 1 | 70 | 51.448536 |
| 5.5 | 95.300003 | 114.4 | 5 | 34 | 115 | 79 | 72 | 1 | 31 | 44.527248 |
| 6.4000001 | 76.800003 | 108.5 | 0.33000001 | 45 | 174 | 86 | 69 | 1 | 62 | 86.29361 |
| 5.6999998 | 49.5 | 74.800003 | 1.41 | 52 | 126 | 83 | 73 | 2 | 57 | 45.1502 |
| 4.1999998 | 60.799999 | 72 | 0.89999998 | 56 | 120 | 71 | 58 | 1 | 40 | 39.371635 |
| 10.1 | 73.900002 | 98.900002 | 0.81 | 66 | 137 | 88 | 82 | 2 | 68 | 66.668167 |
| 5.0999999 | 47.599998 | 70.300003 | 1.03 | 84 | 96 | 68 | 81 | 2 | 36 | 36.207241 |
| 10.2 | 85.199997 | 104.6 | 35.52 | 42 | 137 | 103 | 79 | 2 | 39 | 34.06155 |
| 5.5999999 | 65.599998 | 85.699997 | 0.31999999 | 56 | 123 | 79 | 55 | 1 | 59 | 51.910172 |
| 4.6999998 | 55.700001 | 76.599998 | 0.73000002 | 51 | 110 | 67 | 66 | 2 | 47 | 40.009945 |
| 4.8000002 | 43.900002 | 75.699997 | 8.2600002 | 57 | 147 | 90 | 63 | 2 | 43 | 53.262455 |
| 8.1999998 | 65.199997 | 103.4 | 7.0700002 | 52 | 129 | 86 | 85 | 2 | 59 | 63.714401 |
| 5.6999998 | 56.599998 | 76.800003 | 0.92000002 | 64 | 144 | 83 | 66 | 1 | 61 | 58.001537 |
| 5.6999998 | 57 | 85.400002 | 2.3800001 | 92 | 120 | 75 | 67 | 2 | 65 | 56.564106 |
| 6.6999998 | 53.900002 | 87.800003 | 0.83999997 | 59 | 122 | 79 | 92 | 2 | 54 | 55.998047 |
| 4.5999999 | 58.599998 | 69.300003 | 0.23 | 48 | 112 | 62 | 68 | 1 | 22 | 37.624619 |
| 5.5999999 | 79.800003 | 107.1 | 0.69 | 67 | 130 | 98 | 71 | 2 | 59 | 50.873295 |
| 6.0999999 | 52.799999 | 80.099998 | 0.49000001 | 72 | 117 | 67 | 69 | 2 | 55 | 55.989685 |
| 5 | 105 | 107.7 | 2.9200001 | 46 | 102 | 70 | 60 | 1 | 48 | 31.033155 |
| 5.5999999 | 75.599998 | 93.199997 | 2.25 | 37 | 122 | 84 | 62 | 1 | 53 | 43.822422 |
| 5.0999999 | 49.299999 | 77.099998 | 1.17 | 71 | 115 | 78 | 80 | 2 | 40 | 44.581284 |
| 5.0999999 | 45.299999 | 69.800003 | 0.47 | 86 | 104 | 66 | 62 | 2 | 34 | 45.838638 |
| 5 | 63.400002 | 78.699997 | 0.46000001 | 62 | 123 | 76 | 69 | 1 | 29 | 44.943325 |
| 5.1999998 | 50.299999 | 77.599998 | 0.68000001 | 78 | 119 | 82 | 70 | 2 | 49 | 47.419846 |
| 5.5999999 | 93.699997 | 99.400002 | 0.58999997 | 44 | 127 | 83 | 48 | 1 | 36 | 42.485336 |
| 7.5999999 | 60.900002 | 83 | 1.04 | 127 | 127 | 49 | 75 | 1 | 71 | 81.281235 |
| 5.1999998 | 41.299999 | 62.400002 | 0.40000001 | 82 | 105 | 75 | 84 | 2 | 24 | 34.357563 |
| 5.8000002 | 74 | 85.699997 | 0.69 | 39 | 118 | 72 | 76 | 1 | 26 | 40.841606 |
| 6.5999999 | 46.799999 | 73.699997 | 0.76999998 | 54 | 129 | 70 | 73 | 2 | 60 | 57.00346 |
| 5.0999999 | 47.700001 | 70.400002 | 0.50999999 | 85 | 130 | 90 | 96 | 2 | 32 | 40.798656 |
| 5.3000002 | 53.5 | 80.5 | 0.60000002 | 64 | 112 | 74 | 73 | 2 | 60 | 46.008221 |
| 5.1999998 | 64 | 82.400002 | 0.34999999 | 54 | 122 | 67 | 42 | 1 | 44 | 55.464012 |
| 6.5 | 68.900002 | 104.9 | 0.47 | 56 | 120 | 69 | 71 | 2 | 70 | 67.237312 |
| 5.3000002 | 54.799999 | 78 | 0.25 | 87 | 101 | 64 | 59 | 2 | 27 | 47.242447 |
| 5.4000001 | 55.5 | 79.400002 | 0.46000001 | 54 | 120 | 59 | 66 | 2 | 67 | 54.231022 |
| 6 | 56.200001 | 79 | 0.83999997 | 44 | 147 | 87 | 68 | 1 | 52 | 57.957848 |
| 5.0999999 | 79.300003 | 82.699997 | 0.98000002 | 52 | 116 | 66 | 50 | 1 | 21 | 38.900711 |
| 6.1999998 | 74.5 | 102.8 | 2.4000001 | 48 | 142 | 59 | 57 | 1 | 76 | 79.78862 |
| 5.1999998 | 54.700001 | 77.800003 | 0.64999998 | 48 | 95 | 69 | 83 | 2 | 39 | 32.278168 |
| 5.5999999 | 65.699997 | 95.800003 | 0.44 | 66 | 147 | 85 | 66 | 2 | 62 | 65.935982 |
| 5.9000001 | 52.400002 | 83.5 | 1.24 | 66 | 113 | 70 | 79 | 2 | 67 | 53.034515 |

| Variables | Values |
| --- | --- |
| HbA1c (%) | 4.6 |
| Weight (kg) | 74 |
| Waist circumference (cm) | 73.7 |
| hsCRP (mg/L) | 0.28 |
| HDL (mg/dL) | 65 |
| Systolic blood pressure (mmHg) | 109 |
| Diastolic blood pressure (mmHg) | 63 |
| Pulse rate (bpm) | 57 |
| Gender (Male = 1, Female = 2) | 1 |
| Chronological age (years) | 45 |

DELTA001 BA estimation
