## Supplementary File 3 for "DELTA: Fortifying Human Biological Resilience with an N=1 Digital Health and Dynamic Biomarker Protocol"

DELTA001 Health Optimization

Interview 1

**Administration timepoint:** Baseline

**Interview date:** 10 October 2024

**Recording method:** Audio

**Transcription style:** Verbatim

1: This interview, the objective of this interview is because you will be undergoing time restricted eating –

2: Yes.

1: And will just want to get some insights on your daily routine, exercise, use of technology and any other aspects that you find that might be useful to lead a healthier life. And – to start off to have a baseline, can you please describe a typical day in your life, including any routines or habits that you follow?

2: Typical day, I wake up at five-thirty AM and that is with a sleep start time of nine-thirty PM. So, I aim for eight hours, but with a realistic, my goal is above seven, between seven, eight. So, I set multiple wearables, I sleep with three wearables. One Garmin Epix, one WHOOP and one Apple watch nine and, I record sleep on all of them. I wake up at five-thirty, I wake up and brush teeth everything and I am stretching in the morning, and I am in the gym at about six, six-fifteen AM in that range. On Mondays, Wednesdays and Fridays, I will do what is called the Norwegian protocol, which is three minutes of relatively light cardio followed by four minutes of relatively intense cardio in cycles of four. So, with the warmup, warm down it’s about forty minutes almost, of that cardio – on a treadmill because it is dark outside. I do that on the treadmill so that I can also control speed. After that is done, I will typically take my weight in the gym, three datapoints and then I will do a strength training regimen. And for strength I focus on one primary muscle group per day with a little bit of supersetting. So, I don’t typically do full body workouts, and I will do weights for about 1 hour, and then after that is done. On certain days I will cold plunge, and I will cold plunge two different formats. Five minutes in five minutes out, five minutes in five minutes out, so it’s a total of twenty minutes. A total of ten minutes in ten minutes out but it’s in five by five. Or I will do ten minutes in and then then minutes out. And I will measure heart rate and stress of the WHOOP and the apple, the apple won’t give stress but WHOOP will. And then my mealtime is between eleven AM and two PM, which I consume large amount of proteins, lot of greens, lots of fibres, chia seeds and flax seed together, some rolled oats, unsweetened almond milk and I only use olive oil. And daily I will consume a drink of celery, red cabbage, ginger and now I add a little bit of broccoli and water, no sweetener of any kind. That’s about it. So, I finish about two [2pm] to ensure that I have ample time for digestion before sleep.

1: So, what I hearing is, you are almost doing one meal a day.

2: One meal a day, about three-hour window.

1: Can you describe like any challenges to maintain such a diet?

2: Ah there is no challenges if I stick to the protein, high protein, vege [vegetable], everything that I just mentioned. However, there are days when I will do plant based. Where I will do a full plant-based day. And I try to do plant based two days a week. On the plant-based days, I need to consume more carbohydrates in order to feel full. However, even if I don’t feel full, I would not consider it difficult at all, to kind of get through the evening, go to the meetings, no issues it’s just I don’t feel as full. And the carbohydrates will typically be actual sourdough bread, or I will up the rolled oats content, a little bit more in the bowl of chia and flaxseed. But other than that, it’s no issue at all, and in fact to maintain a eleven-two [11-2] window, actually I am stretching right, it’s not only twenty and – it’s like I am stretching the fasting window to keep that time frame and it’s fine. I can exercise fasted; I can conduct meetings fasted and most people most frequently tell me that they have no idea that I am fasting.

1: Just want to dig a bit deeper on plant-based diet, is there a reason why you want to have days of plant-based diet?

2: It’s a great question. I think that some of the – there is a lot of data out there, a lot of findings and I think that as we age, I think it’s the definition of what I feel is healthy, eating from meat probably should change. I think – I do try to reduce animal protein overtime, there has been some recent studies showing that, certain animals’ protein can lead to various risk of cardiovascular and beyond, but that white meat maybe have a lower risk, but maybe still higher than plant based. One thing of note is that I rarely eat red meat now. Okay I would say – so, if I think about at this moment, I don’t think I had red meat in three months and I intend to roughly, kind of keep it quite sparingly. Maybe a couple times a month total. So, it’s really only chicken and salmon and fish. But two days a week, plant based, I feel – I don’t know if I feel better, but I think just by composition it’s reducing the animal protein, probably is – I am mindful of that.

1: Interesting! Maybe you can discuss more on challenges, is there any external factors like work, or social factors that will affect your diet?

2: That is a great question. I will say though, that if I were to embark on a longer fast, so forty-eight hours fast, which is really my limit right now, I try not to do the Norwegian protocol on the morning of the day when I am going to skip my meal. Because I am going to skip it – I am going to go one day without eating essentially. Erm, and I try not to do the Norwegian protocol just because I think it makes that evening a little harder, that evening that I gotta get through. I would say that – and I don’t know what’s the intervention yet, that is helping with this, but work is – I don’t consider work a stressor. Errr – I think err – I think it’s the function of the sleep that I am getting, which has been quite consistent. I think it’s a function of regular exercise and I think it’s a function of me focusing a bit more on breathing. I think a vast majority of people do not focus on breathing, whether it’s box breathing or deep breathing, erm, and I think it’s reflected in my data. If I were to look at my resting heartrate, I think it is in the mid-forties, if I look at my heart rate variability, which frankly speaking was highly variable before I started focusing on sleep and keeping this regime, and now it’s only going up. And there is something interesting, people would ask me, “the numbers were good, but do I feel good?”. And I think the answer is yes, I don’t take afternoon naps, I don’t need afternoon naps, I would say even a year ago where I wasn’t focusing on sleep, I certainly took naps, but I haven’t taken a weekend nap in I actually don’t know how long at this point. And so, I think external stressors which by the way also measured with devices, whether we tend to feel what the data say. But I would say – to be honest I actually ignored some of the, and we will talk about it probably, but I ignored some of the updates like “oh you are –”, every day I get an update on stress, how much high stress I had and whether that high stress is within my workout or outside my workout. Cause if it is within the workout, you can pretty much know where the stress is from, some exercise. But if it is outside, it might tell you today is a more stressful day and by and large, I have rarely get the ones that I am high stress, outside., very rarely. And that data, I think it’s pretty accurate.

1: So, I am going to – because you say that you feel good, but now I am interested to know– to see if you feel good like do you actually enjoy your diet, the stuff you eat or the amount of workout that you have been putting –

2: That’s a great question. Erm, there are some days that are harder than others. But I would say that I have not skipped any day. There hasn’t been a day that is so hard that I’m like, I just cannot do it today and the data will show that. However, there are days where like I’m just really not feeling it. Let’s focus on the diet first before I get to the workout

1: Okay.

2: The diet, I actually enjoy, I actually feel good when I eat healthy food. Do I enjoy unhealthy food? Also, I do! But I weigh how I feel, physically feel and how the food makes me feel and I strongly prefer how I physically feel after works. Even more than the data. But the data comes close. If I were to – and I am going to dig a little deeper, I get a lot of question asking about alcohol. And I have effectively, virtually given up on alcohol completely. Maybe a few drinks in a year, right? The alcohol can immediately affect the day, which I am not happy with. But practically and physically and immediately for it affects sleep, it affects athletic performance, it affects the next day. And I – it’s so infrequent that I am drinking, that the effects are clear. And I much would rather to prefer to feel better and the data to also reflect that than have a drink which is a [inaudible 11:59-12:00] thing. And the same kind of goes for the fried food etcetera which I still enjoy, but it’s like, as I get older too, I am very mindful of things like cardiovascular health and the impact of having, like one meal that’s not great, two meal that’s not great, three meal that’s not great, you don’t get kind of the flexibility to reverse that. Actually, in some case you can reverse it. So, it matters to me – sorry I am going to take some readings while we are going on but go ahead.

1: Yeah.

2: For – So unhealthy food, enjoy but not really worth it for me but I still, I still do it. I would say alcohol we conquered – Is that anything on the food side that you still wanna know?

3: Just to elaborate on days that feel harder, what keeps you going?

2: Ahhh that is a good question. So, I had tried little tricks in order to get me to like get into the gym and actually finish the gym. One thing I do is – I have I have Airpods, I have wireless earbuds, I like music when I exercise. But what I will do is I will get in the gym, and I will start exercising without the Airpods and I will tell myself if I can simply get myself to thirty minutes of exercise, the second half of the exercise I get the Airpods. Incremental, incremental kind of rewards that I have. And part of it is, you know, we don’t know any data on whether that stuff is good for you or not, all these devices. But it’s also a thing where I kind of try to limit too many devices on it any given time. So, it kind of serve two purposes for me but I do little – I put little rewards into them I try to gamify that. I will say that the data is a big reward for me. Getting just data out is a reward for me, so I think it’s important to understand like how my blood glucose responds to glycogen depletion. That’s an immediate reward, it’s a reward that – I will tell you I did pushups, my glucose was at 3.8 which is actually a little low not hypo but low and one set of hundred pushups and I took glucose right after, which is immediate – you talking about the whole thing was a span of a minute or so – it went to 5.1, from 3.8 and for me that’s a really cool immediate shows that physical activity did something. So, for physical activity, by and large vast majority, I really enjoy it. Part of it is I really strive this mind muscle connection, when I am lifting, I really try to feel the contraction and for me it’s like I just feel good, stretching feels good. Breathing deeply feels good. And I think really it really helps maintain my energy. People talked about this for generations, but I really feel like it impacts positively, my energy, my ability to adapt to travel that I have, different work situations, different deadlines, yeah.

1: Building on this, is there any moment where you have strayed away from your diet and how did you get back when if there is such moment.

2: Erhh, I went on vacation with my family for about two and half weeks, about now pushing two and a half, three months ago. And what I did was –. My non-negotiable was I had to exercise the same every day. And so, I, and in fact I did even more – and I try to add a little bit of a milestone – I was doing less cardio, typically when I am a little bit more tired a little bit more lax, I tend to do more weights than cardio. But when I was there, I actually dramatically increase my cardio, which I feel like carried over to here, which is leading to this Norwegian protocol, which I really focus on which is really great. And I was running outside, air is good but did ordered weights, I had weights shipped to make sure that I can still lift every day, because I don’t belong to a gym back in the state. So, working out, non-negotiable. I did convert closer to sixteen/eight (16/8), typically outside of the shorter eating window and because it was vacation, I let myself basically eat whatever I want. I still bought high protein food, I still had meals that was quite healthy. I did do, I believe OMAD a couple days during that trip, but I let myself otherwise eat whatever I want. And what was really cool was, without kind of even dreading it or thinking about, I immediately switched back when I got back. It was immediately. There was no kind of “let’s do another unhealthy day, another healthy day “but I just immediately switch back. Yeah yeah.

1: Yes, and while we are on this, you mentioned that when you feel tired you tend to do more workouts, more strengths training.

2: More weights.

1: Weights okay. And do you think that there is association between the food you eat maybe on plant-based days, will it affect the type of workout that you do?

2: Yes, I think I can be a bit more tired the day after, doing plant-based. Erm but I haven’t checked any performance data but I – if – the plant-based days I am kind of making sure now that I am full. So now I actually make sure that I am consuming more carbs. I don’t try to maintain perma ketotsis, like I am always in ketosis. I think people are curious about that, like do you eat carbs? I do eat carbs, like some carbs like resistant starches, some of the stuff is important and as we know we are going to look at gut health too. I think gut balance matters and so I think that I could be more tired, I haven’t seen any major trends but I do make sure that I am full.

1: So, it doesn’t really affect your –

2: I don’t think so, I don’t think broadly speaking it affects it. I won’t probably even know if it affects muscle growth or it affects muscles loss, but I haven’t seen any negative impact on – because I am still doing mostly animal protein, so I don’t know if it really negatively affects anything, in terms of performance.

1: Okay, and I think that you have been tracking your sleep your resting heart rate and other health indicators. What are some of – what are some important benchmarks or criteria that you will use to assess your overall health?

2: Er yeah. I think aside from the data – the data yes, for sure because I have enough wearables on me that – by the way I don’t compare and say this one is off, this one is right, or I prefer to believe this one. I just look at all the data to kind of get a general sense of change overtime, I think that matters. I had days when one wearable says seventies for sleep and the other one gave ninety, I had once when they were ninety-one and ninety-two. I had day –. And so, I tend to – but I haven’t had days when I had really bad sleep on one and really good sleep on the other. I hadn’t had one I believe. So by and large I think I take each once for what it is. Everyone certainly have different algorithm, different prioritizing of what is the score, but I look at change over time. And that for me – I think overtime somewhat reflects in what I feel. Ermm, I had days where I feel like I woke out a fair amount during sleep but transiently. But when I woke up, I noted that I was really like sleeping hard, but I woke up and I went back to sleep. But it has only happened several times, and I wake up and the sleep score will be pretty good actually. And I will wake up rested knowing that I slept – I feel energetic but knowing that I woke up several times, but I woke up from really deep sleep. And you can tell, and I think people know themselves, the score might say ninety and you just know that I did not sleep well. And – so I still trust what I feel like the most. But I would say that objective markers like I have not even attempted to take a nap. I have not even attempted; I have not even thought about it. I have flown, I travel long distance, I flew a three-legged flight of which I slept well in the beginning, and I had a good – with layovers, I had probably a good twelve more hours of flying and I didn’t sleep at all on that 12 hours. And to be candid those twelve hours were upright, I wasn’t laying or lounging – the first part was relaxed but the remaining legs I was upright the whole time and so that’s performance, to me that is performance. I stayed awake the whole time, got some work done and by the way getting work done generally what can make me tired on a plane, I kept going watch the movies, got to the hotel, and it’s about seven-thirty, eight, went to sleep by eight-thirty, eight-forty-five and slept through the whole day. My data will show that. I think the performance side is inarguably clear, my run times are getting better, and I think that’s pretty clear.

1: And how would you define a healthier version of yourself compared to now.

2: Compared to now?

1: Yeah.

2: Something I don’t really know for real – I would love to know an objective VO2max, and I am betting that I can do better. And a healthier version of me now frankly it’s probably one where I am actually eating even more plant based to be honest. Erm, I don’t know if I will ever convert into plant-based and the reason for that is we are Asian. And it’s a balance, I am mindful of using carbohydrate to compensate for to make sure that I am full so that I can still exercise right. You gotta, you can do plant-based but some days you don’t have the energy to exercise. We know people have found those sweet spots to be able to do both. But for me because I am still lifting a lot, I am still doing a lot of cardio and I prioritize that a lot, I don’t wanna start packing carbs to do that. Even if you look at protein sources for plant-based, where I am doing chickpea and all the beans that are out there, the carbs contents are also high. Now does that lead to blood sugar issue? I actually don’t know; I don’t know enough about that and especially for myself yet. But I am not prepared to just go full plant based but I think being a bit more mindful of keeping the animal protein lower probably could make sense. In terms of things that are not good for me that I – that could be cut, the alcohol could be cut from two drinks a year to zero a year, which I don’t think it’s going to be a huge gain. Refined sugar, I think I am pretty good about not having that and my ketone measurements will reflect that too. I’ve really been conscious in trying to stay asleep and I think my data actually reflect that too. Generally – broadly speaking, I am actually decent at that – erm – and I would say other than that, something I would like to figure out is how to ensure I can continue to socialize properly while I maintain this, because I have very understanding friends and colleagues, and I kind of w [Inaudible 25:29] – because I don’t think my regimen now is the permanent regimen. So, I need to try to figure what is palatable to me. Maybe, it's Friday nights and Saturday nights, I allow myself some leeway. And part of that may include not recording. Because I think the recording is the driver of my behavior, and maybe it's recording, who knows. Maybe I will find that it’s not that bad, so I will do it all. So I am okay with that too, but I think seven days a week of aiming for like seven hours plus of sleep, my kids, something I go to bed before my kids. I am not entirely sure I want that; I actually don’t think I want that. And they actually sleep relatively early, so I think understanding social engagements – sometimes I had social engagements in the evening where I did not eat, a single bite and by and large people joked about it with me. I was cool, I knew most of them. I don’t know if – that’s not going to be possible for every social event moving forward and I am okay with that. But I think I need to maybe understand that not shutting off any – which I don’t, I still see the people, but I just don’t eat. But I think I wouldn’t mind integrating a bit more when that’s happening.

1: Because you mentioned that you want to sleep after your kids sleep.

2: Or at least at the same time.

1: Yeah.

2: But not before.

1: Did your diet affect like your family mealtime or –?

2: Great question! It does not because my wife also follows the same eating schedule. And I will say that that is – was not always the case. And I think that was challenging in the past. So, that is something to note in that when I was off schedule from her or when she never fasted that was very hard. That leads to disagreement, quite frankly. And that is not a one off, not an N of 1, I – from just the number of discussions that I have had in the last six past years of being here, I know that there is a challenge for other people who want to try fasting, safely. Erm, for my kids, we hang out with them while they eat the dinner, and we are totally fine. My wife and I are totally fine sitting around food, that’s mine – [Drawing blood]. We’ll take our kids out to eat in the evening, we’ll hang out them, but we won’t eat. Because for my wife sleep is also important for her.

1: Maybe just a general question, do you have any advice for people that might want to try IF [Intermittent fasting] that might face similar situation like dispute between –

2: Ya, I think that – first of all if people want to try intermittent fasting, we’ll put the clear thing – they need to make sure that they can safely fast. Okay. So, they are not prediabetic, diabetic, they consult the doctor. Erm for people that want to navigate the family and friends’ dynamic of fasting, if somebody wants to skip a meal – I would say people should not try OMAD as the beginning or one meal a day as the beginning. Err I think that people should maybe try twelve hours fast fourteen hours fast working up to sixteen hours fast and if they were working up to a sixteen hours fast, they may want to try to skip the meal where it causes or it’s the least potentially complicated for the family dynamic. Alright, so if they are working in the morning, they work early they can maybe skip that one, because they see the family. But they will see the family in the evening. Erm I will say that whoever that is considering fasting, fasting is not a “get of jail” free card for eating whatever they want – Erm when they break the fast. Because there already have been studies showing that people can end up more sauced [29:36]. Right because if they overcompensate if they gorged and they binge eat other. I gotta max out this– [continue to draw blood for ketone and blood glucose measurements] yes, other questions?

1: Maybe we can move on to another topic while you are taking your blood glucose. What is your overall experience collecting your own data?

2: I love it! I absolutely love collecting data because erm– we tend to think about data collection as a fixed time point. Right, maybe getting a lab at a clinic etcetera. But in my case, I’ve been able to really understand my dynamics. Err to really see how fast I can change overtime is fascinating and when we talk about time scales, we talk about seconds right. Measurable change over seconds, the impact of lifting weights on my data the impact of resting after workouts it’s just fascinating. Yes.

1: But maybe to a more human aspect, the process of pricking your finger every fifteen minutes is that –?

2: Yes, er. The process of pricking fingers, I am so used to now. And I, so we all know there is different ways to collect things like glucose and ketones data. Erm glucose, we can do continuous glucose monitoring now and there is a technical difference right. CGM is a – continuous glucose monitoring is interstitial fluid and blood glucose is blood right. And I have found in some cases a substantial difference between the two, it could be a function of body fat percentage, position of the wire – so there is a lot of reasons for that. And they are both still are quite accurate. But the sustainability of collecting with CGM for me at least is challenging because it’s a hundred dollars for two weeks. Err and – it’s dramatically cheaper to measure glucose from blood. And then erm another thing is there are some ketones wearables out there, I haven’t used them, so I have no comments on those. But when I do things like glucose ketone index, the G K I measurement, I would prefer that they are both from blood, in fact they are from the same draw right. One – you prick once, one drop comes out get glucose, squeeze again you get the second drop you do ketones. I would prefer to align that. Pain wise it’s like a none issue at all, it’s like a zero issue whatsoever, I think I have collected in a moving vehicle before, it’s fine. Er, practically speaking, most people out there do blood pricking, right and I actually wanted to understand what it was like to do it, some diabetics are pricking many many many many times a day, I wanted to know that it was like. And it has not – so pain-wise I am totally fine. It has zero impact on me doing pull ups, lifting weights, I’ve collected ketones after icebathing, when your hands are like freezing, so I have done it under all circumstances, so and I have done it on travels I have done it in malls which people have to do, right, people have to do. Part of me was like, was I self-conscious was it challenging to do it and somethings it is a little challenging. I have done it in a restaurant but that’s what people have do, especially for those who need to do it. You know, but I have zero – I would say it’s not comfortable the first two days, which is a long time ago but now it has just become a part of life.

1: Was it easy to use, easy to adopt such technology?

2: The first couple of times, it took some times, it took me awhile it took me about, well actually not long, it took me about three tries to draw out enough blood to read, because you know ketones you need a little bit more than glucose, took me three tries to figure that out and from then on, I am fine. I – once every hundred finger pricks and I will take a reading, and it wasn’t enough blood by accident. But otherwise, it is really easy to use, very durable in terms of how long this system work and how consistent they are. I’ve taken reading right after the other and there is no drift, that’s why they are FDA cleared. So, it’s been enjo – I would say that it has been very enjoyable versus any sort of inconvenience.

1: And when adopting a new technology apart from consistent like accuracy, you mentioned being easy to use, are there any other features or factors you look out for?

2: Er, I – the time it takes to get the reading matters to me. I am, to be frank I am sometimes a little self-conscious collecting. Most of it is because I don’t want other people to be scared of a guy pricking fingers next to them right. That’s really the reason, I am not self-conscious of my reading the data in the public, the ability to do it quickly matters to me. I don’t wanna have a whole operation set up and I have – and I don’t – and actually it’s very – in my opinion I still consider it discreet. If you’re drawing blood, I would prefer that it’s as discreet and part of that is speed.

1: Okay, and how do you harness all the data from your smart watches.

2: Okay so I was getting to the wear, perfect time, I was getting to the wearable next. Which is erm, I don’t know what to expect when I put on three wearables and er, it was – I try to let this be as organic. I am trying to understand what the – actually I am an optimist, so I like to see what I like about each one and then I kind of just let go whatever that seem harder for me. So, I like the ease of the apple watch, such that it is very easy to start and record exercise. I think that is probably one of the most critical things for me. And I would say the sleep analysis is easy, you wake up in the morning your data is right there, you can look at kind of breakdown of different sleep type over times and erm I love that aspect. I love the ring closing, I think the ring closing is pretty important, because as a gamification tool, I think for me it helps a lot. It’s one of the primary reasons why I am still wearing it, in order to kind of completely close my rings for a year, it’s a goal that I have. Erm, the ring closing works, like I specifically stand up in meetings to make sure that I can close standing rings. I will specifically stand up and go for a walk just to do that, and sometimes just standing you know, given the way this thing works, it’s not going to give you 1 minute of standing it doesn’t register, so I actually go for a walk. And of course I am okay with that, that’s fine. It’s the clearest for me that I am meeting my research goals, and I don’t really pay attention to the, what the calorie number is. If I’m like – I don’t go around saying that I burn 600 calories. I don’t know, I don’t know if I did. But I do trust it, it just helps me know that I am meeting some goals for the day. There is a function on the apple watch that I liked that a lot of people I think from at least people, most of my friends wear apple watches, that they don’t look at which is the basal metabolic rate. Okay, which is the total calories you can burn for the day which you can get by clicking on the rings minus your, the exercise burnt, the two numbers right next to each other, you can just subtract. And I like that it’s dynamic, I don’t really know, I don’t pay too much attention to what that number is, but I generally try to think about what the range is and that actually changes day by day, which is actually different from err, say from a Garmin for example. The Garmin I think is based on the body height, weight, whatever that kind of just max you out, everyday it’s the same number. And I like to see some dynamic there, so I think that is pretty cool. Err, so Apple I think it has been the easiest, gotta charge it frequently, so that has built a habit into me, every day I am checking it and I making sure it's charged so that I can keep collecting the data. So, this is usability, I am going to get to like how do I synthesis data later, in a second. So, the WHOOP, I was apprehensive about adding the WHOOP and the Garmin, because I has been so accustomed into using the apple. But I found both to be really easy to take up. I would say that the kind of warm up period was a day or two. You know, I was used to battery needing to be recharged on the apple watch, so I think I turn on the low power mode, for the Garmin. And then I woke up the next morning and it’s like “no data”, then I like what happen “no data” and I was like I think the watch is broken and I realized low power mode it won’t talk. So, I think I didn’t record it for one or two days, so after that both of these, I – I’ve come to appreciate. Erm, I think the WHOOP user interface is really cool. The bright colors – I will say, I am not sure, maybe I will get some more, I can collect some data on this, when I share some of my WHOOP screen on social media, there is way way more views than on the Garmin. I think the vibrancy carries through – and even before I saw that, kind of crowdsourcing feedback, I was already drawn to the vibrancy and the way the layout’s done and must I – the UX/UI is pretty cool, I think for the WHOOP. I do like the Garmin’s dashboard, I like that you can click on like “show more” and you just see all these different things together, I think it’s pretty cool. That’s probably, that actually one of the reasons I am still using the Garmin. Because I did stop using the workout recording for the Garmin. I don’t record reps, I did find though that rep recording is make – me causing me more diligent about equal reps on both arms etcetera but the kind of changing the weights and having to click on click off, I actually found out later on, later in the game how to turn off the beeping sound when you start and stop recording, I was self-conscious actually, I was self-conscious in the gym because everyone can hear it. You know and I don’t want to drive people crazy. I turn it off eventually and other people in my gym have it on right now and it kind of brings back weird memories. So erm, I stop using the weight and I still prioritize weight training, I don’t use that anymore, but I still like the dashboard. Because I like the HRV balance part, now let’s get into the data, I like the HRV balance data, because if I kind of just resort to the public, social media, just trying to max HRV as high as you can is what people think. But they have this HRV balance kind of insights which I think is pretty cool. I do like looking at the resting heart rate data on this, I like that I can sit there and look at my phone and just get live heartrate data, I think that is pretty cool. On the dashboard, sometimes I’ll sit in the meetings, and I’ll just turn on the dashboard and I will just breathe and just watch my, see if I can kind of decrease my heartrate by doing some breath work in the meeting. You know and then I –if the meeting has like some more deep conversation, some more deep challenging things we have to address, I like to watch live, how my heartrate is responding to that, and I think the Garmin is cool for that. So, I used each one for very specific things. I see look at the sleep score, I look at my body battery to see if it thinks that I recharged well, I do feel that the WHOOP is a bit more interventional, I think the WHOOP is a bit stricter, it is almost – it’s is like a real coach. Right, when it tells you “Your sleep debt went up” you should try to do this, “You maybe overreach on your workouts, today making you can chill out”, “you are high stress, today was mostly from your workout” versus it is outside of your workouts, which makes you wanna think” hey am I actually stress right now”. You know, so I – I derived different data insights from each one so it’s less of a comparison really. I go each one for something that I need and that’s why I like having them.

1: And how did you interact with these three wearables, did you like use it directly on like use the screen or did you use it through your phone?

2: Good question, I use – outside of the ring or the apple watch, I use my phone entirely. So, I don’t use the apple watch like by clicking, I almost exclusively operate through the phone for everything. The WHOOP for sure there's no screen yeah and then for the Garmin it is the same I virtually don't really directly work with the – the screen and which actually was a little surprising to me because when I first got the WHOOP I was so used to screens and now I realize I think I'm totally okay without the screens I actually really like the WHOOP yeah charging matters too as we all know the Garmin and the WHOOP charging requirements are less stringent and I think that has also helped

1: And is there any particular features or biomarkers you wish that you can track with any of these wearables.

2: Uh certainly, I mean I think glucose is something that people have been striving for. If there was a better way to track, better way to track reps and I know that other wearables are trying to make that easier too. But the – the – the process of having to manually record reps I think has been challenging for me and I didn't really know that that was going to be so hard for me to do that because I – I do like lifting so much… and you know what didn't draw enough flip gotta force more through and please don't give me any almost almost – almost here we go here we go it's gonna make it it's gonna make it yes, we did it [Drawing blood].

3: If I could just ask, please for someone starting out like which one which wearable would you recommend for them to like –?

2: Apple Watch probably. I think it's the easiest – I think it's easiest, if anything just to kind of for the rings etcetera, but I think that overtime people probably have things that they're more interested and I think some people don't prioritize strength maybe sleep you know take a look at an Oura or any of those the WHOOP or the Garmin. Yeah, but I think for just general insight on people who wanna like start exercising more – because I think that – and I don't know if this is by design, not sure, but I think that a wearable – I like more data I want someone I want to know if my sleep was bad or good. For a person who maybe is starting from scratch some of the wearables might be a bit harsh, actually right. It's like ohh you sleep really bad – it's really bad it's like that I don't know I don't know how that people will respond differently to some people will be like that I'm going to get better some people like I now I know that I don't want to know, so I'm done you know so I think that the Apple probably I what would start with yeah.

1: And is that a particular way you like your data will be presented by – by the wearables or –?

2: Yeah, I need the – I need the numbers the – the quantified outcome to be as clear as possible for one thing. Okay, which you know for apple there is no sleep score right, and so I would – I wouldn't mind that I would like some degree of – because I and and this getting to the next part which is it's really important for me to see trends. And because I knowingly modulate the intervention, I might be fasting forty-eight hours, I might be fasting twenty-four hours, the longitudinal matters even more because that's an even richer dataset. It's modulated intervention with longitudinal output. When I cold plunge, I already modulate five by five or ten and to be able to really clearly see those trends in a like cleanly presented manner matters. I can – there's – there's this is where there is variability, okay, across the wearables. The Apple Watch you can just turn on cool down, you can turn on cool down, and then you can use that as an exercise to track your heart rate. Because I can't turn on – if I want to turn on my heart rate monitor then I think I gotta go pull the raw data in order to see what that looked like during the – the – that that span of time, I won't even know, okay. Or do I cool down then I can map it out but then I can't like zone in on specific time points. Whereas on the WHOOP I can, right it'll give me the readout and then I can just put – hold my finger down and it gives me a marker to tell me that was my heart rate at that time. And then it gives me comparative stress during that exercise, and I've seen I'm already seeing evidence of dynamics of really interesting responses. And so, I think the ease of being able to dig a little deeper matters and having really clean images to look at matters. I don't think that's going to be a priority for other people and that's maybe not the topic right now but but for me I need some of those insights pretty quickly and consistently because that's part of my whole my whole gamification process. Yeah.

1: And you mentioned that when you share the WHOOP data because it's brighter.

2: Yes.

1: You you gain more views, so have you been actively sharing?

2: Yep, and we actually consulted, we've consulted on that and that was it was deemed to be okay, and I share on occasion, and I don't, and I also share data that I think is pretty simple data one will be my resting heart rate. One will be my sleep score and then rarely I'll share some I've I've previously already shared ketone and glucose data. Really interesting when I do this to get feedback from the community, in terms of people really having not seen this before actually. And so, I think it's really cool to still understand really where are my friends at in this kind of rapidly growing area of health, wellness, longevity, healthy aging, where the people I interact with the most where they're at, the questions that I get I think that's really cool.

1: And the sharing data like having these social circle, finding where your friends are at etcetera, sort of motivates you to lead a healthier lifestyle continue maintaining one?

2: I think it does because we've – I've had friends – I've had friends who were like I wanna join and I've had friends who have joined right. Not for research just to live better. Just to, because some of them were like ohh I never thought that Dean, because I tell people I never prioritize sleep, but I tried it and feel better right and they're like I'm gonna try and they're they feel better. There are people who tried fasting, got to be safe, but they try it and they it's actually worked well for them. And then I have people just curious saying like ohh what is this data show and is that good or bad. And for – and there there are people who don't know that lower resting heart rate generally speaking is a good thing, and even just imparting some of that knowledge on somebody is great. I've had friends who have refused to give up luncheon meat because it's a way of life, I get it and I'm not telling them to give up luncheon meat right, but then they're like wow the sodium's really high, right so it's not like they see me eating spinach and they're like I'm gonna eat spinach now, they're not gonna – they're – I'm not asking them to nor are they planning to. But then they're like Oh well you know maybe I could eat a little less luncheon meat. And then they look at the back and they're like this is packed with sodium nitrite E250, I read online as horrible you know. So it's people do some of these things do motivate some people to look at whether it's sleep directly or it's other aspects of their life that they are willing to make changes to. Yeah.

1: And maybe we have touched this before but just wanted to know it better. Like how have these wearables helped you to maintain a better lifestyle how does it affect your routine, exercise and diet.

2: Uh but exercise for sure it is kind of I have a built-in habit now of knowing to track workouts and to close my readings for sure. At some point I'm going to give this watch [Apple Watch] to my son, soon. And so I am curious to see how that may or may not impact my – I don't think it will change it. But I might miss it I might and I have to give him the watch by the way okay I can't like tell I already knows he's getting it so I can't be just kidding so you never know I might give it to him, just to go buy another one after, you never know but I had I had planned on not buying another one for myself but you never know, that might end up buying it even more expensive one. But it really has become a part of my life in terms of closing the rings to make sure I burn minimum number of calories stand a minimum number of minutes and get a specific amount of exercise in. I, I look to the data every day. I actually look at the screens every single day to kind of take stock of that day for myself. And then I look at trends I look at my sleep efficiency trend every morning it's it's one of the first things I look at when I wake up, not just the score the efficiency. I want to make sure that I'm actually doing pretty well at spending that time there asleep, every that's the first thing I do in the morning when I wake up. Yes, it's I think it's a key stimulus for me.

1: And if the data doesn't really match your expectation or if it's not trending towards what you expected do you notice like any behavior change or do you like incorporate new ways to improve?

2: On days where my sleep isn't great, I will – I the first thing I do is try to evaluate maybe why, it wasn't that great, and I tried to kind of reevaluate my sleep hygiene for the night before. I do – and and you know sometimes I feel like maybe sometimes maybe I need a little bit too late. Sometimes I have to you know do the meetings I push but that's it's rare my my really bad thing is super rare. But if if – if I wake up tired or and or scores come back not great, I don't I don't really change much of my routine though. It could have been that I was just running some thoughts through my head the night before on a thing that needed working on. I don't really modify anything. I'm still doing my work I will still work out in the morning. The WHOOP might tell me to work out a little less harder actually it will tell you it'll be like this is your you know try not to go past this, but I don't think I've had days where I did suboptimally two days in a row I don't think I've had that. Yeah.

1: You mentioned sleep hygiene what what does that means?

2: My sleep hygiene is I generally finish eating by two PM every day where that is not eating late into the night, and I try to finish that early. I try not to drink water so if my sleep time is nine-thirty right, I I try to stop drinking water by six-thirty PM. And and I also drink I try to drink a lot of water throughout the day. I think I'm well above minimums that people should be doing I drink a lot of water. Used to think that I'm gonna wake up super thirsty at night and since I've enacted this, I've had zero I've actually had zero wake ups yearning for water, but my sleep has been pretty good, so the water drinking is something important because I just don't want to wake up to have to use the restroom. I have eliminated screen time completely right sometimes; I'll lay there catch up on social media I don't I don't do that at all anymore. I – I do sleep in a cooler room I think it's at about twenty to twenty-one [degree Celsius], some people can sleep even lower than that, I keep it like twenty to twenty-one and and I just do some deeper breathing when I close my eyes, I'm usually asleep pretty quickly. Yeah.

1: And yeah maybe –.

2: One thing I will add. There are times when I wake up, okay it's like it's too early, okay it's 3:30 in the morning right, I have started to view sleep as exercise right. Sleep is a form of exercise and when I wake up at three-thirty I would say I used to panic a little bit my bag [57:40] Oh my God it's three-thirty 1'm gonna be tomorrow's ruined right. And this is maybe years ago and then I went through a phase maybe a couple years ago where if I wake up at three-thirty guess what I'll just like surf the web at three-thirty until I get tired again and then I will fall back asleep. I'm like I'm not gonna worry about it I'm just gonna surf the web I'll be awake again and I'll fall back asleep I'm all good I'm not even stressing. Which is also not great by the way, but then I now view sleep as exercise so I've actually laid there with my eyes closed and I'm like you know I'm just gonna start again, I'm gonna start again, I know I can go back to sleep I know it's too early okay. I know I can go back because I used to watch and, in my mind, and I'll laugh you know I used to wake up watch cartoons I go back to – watching the cartoons didn't make me tired yeah, it didn't make – it's not exercise in fact you probably woke me up even more. Right and so I will – did I say cartoons not cartoons? Because I've been watching Disney documentaries which aren't even cartoons but watching you know watching movies whatever, so then I just lay there with my eyes closed and I breathe and I'm like, I'm gonna go back to sleep right. The data is gonna be awesome I'm gonna go back to sleep, this is exercise, getting back to sleep is exercise for me and then it worked! it totally works! I just do some deep breathing, do a little box breathing, I I treat with equal non nervousness as before which is a much better thing to do versus what and I go back to sleep yeah. And the next morning I wake up and Apple will catch your awake period. And it goes right back to sleep.

1: But when – when you are practicing like – when you're watching movies previously, do you wake up later compared to –?

2: Yeah, and I would need to – I would – I would – it would – you know, it will have to be days where I would maybe allow my – be able to wake up half an hour to an hour later, and I would have sleep deficit for sure.

1: Yeah.

2: I won't like me like I'm going to sleep in now until nine-thirty in the morning and that – I just can't do that so, it's days where I'll – I'll – I'll still be in deficit, but I'll get another hour in or something like that. And I won't be up for hours I'll be up for another thirty minutes or something and I'll – I'll get it back somehow.

1: I see it.

2: Maybe I would nap later, which I don't do anymore, because now I can go back to sleep no more naps.

1: And for this study because you'll be taking your blood.

2: Yeah.

1: Like what you're doing right now.

2: Yeah.

1: And there will also be fecal testing.

2: Yes!

1: So, what are your overall experience doing fecal test, do you enjoy taking your own stool, guide me through the experience.

2: The experience fecal testing is really easy, so it’s actually really really easy. I was initially intimidated it was a new thing for me, but it's become totally fine. Its – its instructions are super simple and that both samples were successfully analyzed so no issues there.

1: And what are your knowledge on the – the results that you received from your microbiomes, I mean the – the fecal testing and –?

2: Yeah, so far when I – so my baseline – by the way just so I've got about twelve minutes left is that okay and we can –

1: Yeah, I'm closing, I am closing it soon.

2: Okay let me just tell I have another – let me just tell her I'll be about five minutes late ten minutes looking for one second here – one sec(ond) [Texting someone to inform that participant will be late for next appointment]. Okay, so data wise um my – my fast – my OMAD eleven to two is my baseline right. That's what I'm used to, that's my baseline reading, and my gut balance is actually rated as great, which was interesting to me. My pathogenicity scores which everybody will see if it's – it's really low, low obesity, low serious disease, low type two, super low they actually give a spectrum score on that. My fusobacteria is like nonexistent which I – I hear it could be a good thing because it's that's the one that's the biofilm and you know that's the one that contributes more to pathogenicity of disease. Um I could – I might be able to use some more actinobacteria but the populations back to royalties [1:02:14] and another population they go up which is actually seen in other fasting populations. Something that I am curious about is, what this means for me. Um I – I – I may have misread but it appears I have – I'm low on microbes that deal with processing carbohydrates. Which is understandable I don't eat a lot of carbs but, I'm rated low there but I don't know if that's bad or not bad or bad good whatever because I think it I don't know if it's in comparison to the population and it there's something in there that says this this is low in comparison to a population where of – where this particular type of dietary thing is common but mine is uncommon. So, I don't know if low is just low, it's just a reference point or if it's not good. That's why I'm doing dynamics and if I compare against my three meal a day gut, which was mostly plant based and had more carbs my gut balance went down – my gut balance actually went down like the balance of the different microbes. I don't think that's a bad thing necessarily I think it's – I would estimate it was down relative to what – it may have increased another type of microbe that you know or – or one that already exists went up even more right, that that's an example of balance. My pathogenicity really wasn't a huge difference at all but something that's unique though is gut health tests look for potential diabetes and – and metabolic health I'm willing to think that most people who do gut health tests don't have the degree of metabolic data that I do. So at least for that maybe we can try to draw some interesting conclusions, where maybe this particular microbe at least for me is not that important.

1: Yeah.

2: Right, because there's things that say ohh this may tend you towards type two diabetes even though my type two risk is super low but there's like – but this one has been in – this particular microbe has been shown to be important. But it appears to me has no bearing right because my HH – my A1C as we know in the other paper super – super Okay. You know, and I if you – if we average out all my other blood gluc [glucose] I'm sure it's even better potentially now even though I'm not even sure if you can get better from that one right. So, that might be cool because I doubt anybody has the degree of metabolic flexibility data paired with the – with the gut microbiome data that's going to be cool.

1: So, you're just going to observed trends and the dynamics?

2: We're gonna – I'm gonna do now – I'm pretty confident that I can do a forty-eight hours fast and collect stool both times. Yes, I'm pretty confident now. I stress tested it without doing any collection for microbiome right, but I think I can do it yeah.

1: And the last few –

2: Sure.

1: Maybe the last question.

2: Sure sure.

1: So, you – because you say mention about forty-eight hours, so I was thinking when – you're alternating between OMAD, and forty-eight hours do you notice any mood change behavior change apart from your fitness routine that you – you mentioned earlier?

2: Interestingly okay, so, the forty-eight hours fast is where some of the richest metabolic data will come out right. Because the morning of which are about forty hours that's when the ketones are high that's when I can really track dynamics that's what I'm doing. I can still exercise fine, I – I did one-thousand pushups at forty-two hours no problem.

1: Yeah.

2: Right, our body has energy which makes me wonder right like, I've not eaten any calories for forty plus hours we have the energy to do – to do great okay safely, right. Mood wise I will say I have I – I will notice that that night where I've you know I've skipped the lunch that day I'm going to go into the evening that I – I – I can get hungry for sure, but I know that when I wake up in the morning, I'm not going to be hungry at all. I just know that right and – and hundred percent of the time it's held true. On certain occasions I'll have a little bit of sleep interruption. But I think I've gotten over that, in fact my most recent forty-eight hour fasting my sleep score was like ninety some [something], ninety-one and that was fine. Mood wise, I think there's zero difference I – I attended a meeting the other day when I was thirty hours fasted, and nobody knew, and I told them they were like that's shocking right they would that people joked that they wouldn't be non-functional you know it’s, but it takes practice for sure. But I - - I don't – I don't – I don't notice mood change. I've done impromptu forty-eight hours fast and I've done planned. Um on some I – I – some days I’ll feel better than others though like not less less hungry but then there's no real difference in the end yeah.

1: Okay thank you thank you for your time.

2: Anymore?

1: Is there anything you want to share it – is there anything that you want to share that we have not discussed?

2: If there are, am I allowed to follow up with you?

1: We will have one more, we will have one more yeah.

2: Oh good! I probably will have more.

1: Okay, actually I have one last question if we have time.

2: Please, yeah

1: Do you have any advice for someone that's trying to make a sustainable change in their lifestyle towards healthy?

2: That’s a good question, I think that uh I think people should start as – as with – with one thing but that is profound to them. Okay so I don't think it's realistic to say, “let's fast sixteen/eight (16/8)”, “let's give up refined sugar”, “let's sleep nine hours”, it's not something happen yeah, I think that's like a combo therapy too much too fast.

1: Yeah.

2: I think people should find one thing that they deem to be important to them, that's doable, but not easy right. That's why I always use example of removing sugar from coffee because it sounds easy but it's not easy. Especially people who've done it for twenty years but it – it is like measurably impactful on your health when you consume that much less sugar in the morning and in particular in my opinion. And so, I tell people it's like four weeks of commitment where you can potentially see substantial gains. What does this substantial gain? You cut out sugar from your coffee for four weeks you – you – your – the – the weight may not change dramatically, although it actually still might to some extent. But when I say potential gains is if a person can get over that hump, the next thing that they take on, especially if it's self-initiated, I think it's more likely to be stuck to, adhere to as well and then then may come the third. And that was me! Right, I was sugaring coffee I remember I used to go to IK – IKEA and free coffee for – for every member right, so I was milk and – milk and sugar and eventually no sugar and eventually no milk and then now I even cut down the coffee. Ohh by the way, I didn't mention my coffee intake generally stops by seven AM, seven AM. I do basically one cup of coffee that's it. Okay no more [inaudible 1:10:24] because I don't want any impact on my sleep. Okay, uhh – but anyways back to you know finding one thing. And eventually if – if that one thing they pick can have data tied to it that might be a great – even more powerful. So hypothetically if a person's gonna cut sugar out of their coffee what's the data they can get? They – if – if it's okay with them cost wise, put a – put a glucose monitor on, before and after they will see a difference. They will see a difference in one day, they will see a difference in one minute. Right, in the sense that yesterday you know what the peak looks like today you know what it’s gonna look different.

1: Yeah.

2: Okay, they will see that change clearly, and then from that population there will be a sub population where it this could permanently change their behavior just by being so fascinated by something that they never knew before, right but find one thing.

1: Okay, yep that’s a very good advice thank you.

2: Thank you we'll talk again.

1: Yes.

2: We will talk a – [audio recording cut]

(End of Interview 1)

DELTA001 Health Optimization

Interview 2

**Administration timepoint:** End of 3MAD Phase

**Interview date:** 24 March 2025

**Recording method:** Audio

**Transcription style:** Verbatim

1: Go okay. So because I think you you went to through multiple cycles of three meals a day for this study and um can you like describe a typical day in the three meals a day phase?

2: Yeah, I would say the first thing I mention is for three meals a day I have to actively schedule it because or else I'm afraid I will forget... To eat the first meal. And the way I actually have to do it is I have to put uh one of the like mackerel. I eat mackerel as my first meal, cause it's I don't have to cook it. It's canned, packed in olive oil, it's healthy. I should put the tin on my dining table the night before, so I don't forget. Um one thing I will notice my three meals a day, it's important that I don't use it as a chance to like eat whatever I want. So I I don't do that. In fact, um I will still keep it to my … almost everything is the same, I would say. Um, because it's three meals a day, I... I may inadvertently prioritize a little bit more high intensity training, uh, just to just to process the food and so to do that, maybe do a little bit more oats, uh rolled oat uh in my meals, which I still supplement with all the same stuff. Flax(seed), chia(seed), etcetera. But I have to remember, and um and it's yeah, it's it's a lot of food for me. It it's I don't I would say calorically, it might be not a huge difference, but in the end I still think it will tend a little higher. Yeah.

1: What about the timings of when you are consuming…

2: I tried to do it around we first meal around 8:30, second meal around noon, and then the dinner I try to actually eat it pretty early still around five or six if possible, because sleep is still a part of this, you know?

1: And… talking about sleep, did it affect? Like…

2: Um, okay, so I will say I do wake up a little bit less comfortable from the sleep, but I think that I will add there may be one minor confounder for part of… we just finished a week, um cause I was on travel before, but not immediately before, but by the way, my data is very clear, my HRV on the Garmin is like good, even even in travel, post-travel drops a lot and then back to normal. Um I will say that I actually don't think the… if it if it affected the sleep, it was minor, maybe on the recovery side of it, which may or may not be real, but I say I do wake I will say every day when I woke up, I was like, I'm not as comfortable.

3: Could you elaborate…

2: Like I feel like I'm packed with food. Not an upset stomach per se, but I can, my stomach is not super happy. Because as we know, my my usual is I don't eat until after like after 3 PM. Right, so I would feel like I have to go to the bathroom, basically.

1: And did it, like, affect your mood or the affect anything?

2: No, I wouldn't it affected my mood. I would say that it was definitely something I had to prioritize on my schedule. And I think that is not didn't make me mad, didn't make me sad, it would just made it more like that's something I have to allocate time for that I can't allocate for something else. I would say it was inconvenient. That might be one way to put it.

1: And and because you feel, can you maybe elaborate more, like, what do you mean by feeling incomfortable uncomfortable?

2: I wouldn't call it an upset stomach, but it's more like I had to go straight to bowel movement directly.

1: It’s easier to collect the data haha.

2: Yes. Which I did that. Right. So that part made it easy, but it's just like I felt very full. Right? I felt very full I felt heavy. Not like weight heavy, just felt like weighed down, right? Um un I wouldn't call it uncomfortable, but I didn't feel as sprightly. Maybe even slightly less energetic. Yeah, a little bit more groggy waking up.

1: And and did it affect, like your exercise routine? Because you're feeling...

2: Nothing too noticeable, but I would say it the immediate wake up part and to be fully candid, I wake up, I go straight to clearing out, which I do every day, by the way, very healthy BMs bowel movements every day. But this one was more like urgent. I would say the urgency was more there, and it did wake me up. I did wake up fine after. But uh I don't know if it affected my performance, but I'll say like, I wouldn't do it. For me, I wouldn't do it regularly if I wanted to keep performance up. Just because like I think if I'm gonna wake up groggier or heavier, I'm pretty sure I had some point, if not already, it will affect performance.

1: Okay. And because you … I … I guess you on intermittent fasting right now, is there any frictions between the switch from the three meals?

2: Oh no err no. In fact, there's no extra pain of switching back. In fact, it's it's it's simple to switch back for me. Yeah, it's I and I did it immediately.

1: And maybe you can talk a bit about any challenges that you face during three meals a day apart from having to schedule it. Do you are there any other challenges that you do you encounter doing this phase?

2: Ummm… Not not fully aware of any. I suspect it's because I don't eat I don't take it as a chance to eat unhealthy. I think it's just a bit more. Just more over. I don't I wouldn't say no increase in irritability. I wouldn’t … Maybe it's a … for me, it was a little annoying. I would call you know, inconvenience was my nice way of putting it. I think it's a little annoying to have to keep eating. Yeah. Yeah. I think I've developed I really think I've developed a routine. And I would say heading into sleep, heading into sleep. I just yeah, I like feeling like I feel like I like feeling I'm not still churning food and processing food. You know, weighed down. You know. Although what? Let me let me just think about this for a second. Did I… Actually, it would have to be this week. So I I do think I had one or two days, one day where I woke up because I think maybe salt content got to me for whatever I ate the previous night. And I a little sweaty uncomfortable, but it was only one night of the week. And I'll say that that I don't I can't recall of that ever happening when I'm on my usual routine, because I'm done eating so much earlier before sleep.

3: You mentioned you tracked like your HRV and your sleep as well. Uh, how do you see any other noticeable differences in the biomarkers when you engaged in this three meals phase?

2: Ummm. There may have been some… so the HRV is down oh, I mean, I don't know if this stretched too much in, but I think that our resting heart rate was up.

1: Oh.

2: Yeah. Which ermm let me just uh just send this. Yeah, um usually, if my uh HRV is down, my RHR will go up. And I think there was a couple days, but I cannot tell if that was a remnant of recovery from travel or if it was – am I allowed to look?

1: Sure.

2: Yeah, let's look. What was last week's dates?

3: Sixteen.

1: I think you had it three meals from 17^th^ to 19^th^, right?

2: So you know actually I was in the green already, but if we look at the what was it the 16th?

3: 16^th^ to 20^th^, if that's the week.

2: March, yeah, that was 17^th^. That was last week right?

1: Yeah.

2: 17th, it looks like I started to climb back, so this would imply that it was mostly jet lag. But it was low, still low. It was still low and and declined. It's still climbing back. Now it's actually pretty good. Oh, you know what? Oh, but you know what? I have this. Let me see if um overnight averages ah, I can't pull it up anymore. From this yeah, I mean, it was 49 millisecond overnight on the Tuesday, which is the 18th. And then now that I've cleared Tuesday, Wednesday, Thursday, Friday, now I'm at like 64, 68. So it's climbed substantially since, but I cannot know, I don't know if that's a recovery thing, but I will say that it definitely didn't help my sleep. Eating three meals didn't, like, make me sleep better. And I think that's regardless, because the comfort I I just the comfort wasn't there. Yeah...

1: And because I… moving back, I… just talk a bit more about exercise, because moving back… to talk about the energy level a really a bit groggy. And talk about doing like the…

2: I'm gonna pull up some info on that…

1: Do you like notice um any differences, like, doing the workout before the workout or after the workout, like recovery… and more more muscle aches, etcetera.

2: No, uh, but interestingly, very interestingly. If you look at my recovery and the WHOOP recovery it is on WHOOP. Uh, because I my Garmin recovery is not dependable, because I don't record working out on it, so I don't think I used that component. But waking up on the 17th. Okay, so this would be attributed to the 16th, I think, which is the Sunday night. My recovery was 97% on the WHOOP. Monday I ate, so Tuesday when I woke up, it went from 97 down to 71. When I woke up the next day I went down to 66. So 71 is still green, by the way. Actually, you can just see it. This is my so this is uh Sunday night. I usually sleep pretty well Sunday nights, night. Because I'm usually very well rested Saturday nights too, so I actually time. This was the next night, then it goes down here, then it goes down even lower, and then it slightly gets better from Thursday night, but then from Friday night. But, you know, I'm allowed more sleep on, usually on Saturday and it goes back. It literally dips, it goes down, down, down, down, dips and then comes back during that week. Do I remember anything about it? Uh, I don't know, but I do remember at least one night where I was like ,… I woke up sweaty. And that's what happens. If I get a steak I didn't have steak, but if I eat something salty, Korean barbecue late into the evening… when I say late, I mean like six or seven for me is late. I will not sleep well. Like I'm going on a trip this week, Father's 80th birthday. I specifically tried to book him a nice steakhouse for noon, but they emailed me back saying we're not open because of uh Ramadan.

1: Oh.

2: And so they were like, we only open for dinner. And so I was like, please give me the earliest seating, which is 6 PM. I already know I'm not gonna sleep well that night. I already know it.

1: And I guess I see that you are interacting with data. So do you know there any differences in the way you interact with data or like, do you check more regularly for…

2: I'm more nervous. I'm a bit more anxious because I I'm and the thing is I based on experience. By the way, this didn't I didn't, you know, this didn't change, I think my sleep performance very much because I still prioritize the sleep. I still got pretty good hours actually during the week. But but it makes me more anxious because I it's like I know that eating into the evening will not help me sleep. And I think if anything, it'll be worse. And I think I also remember some other info from that, but yeah… Yeah, in fact, if you look, I think it was one night, yeah…Thursday, Thursday to Friday, not good on the sleep performance, Thursday to Friday on the recovery was probably Thursday night. And then what did I do Thursday night?... Oh yeah, I had to eat later because I had to go do an interview. Yup.

1: What about your blood test results er...

2: Uh that was interesting. Microbe, I won't know. Testing testing samples delivery easy. Um but the blood test was what I expected, but super cool, by the way. So I do Wednesday, Friday as well, because it I this is gonna be a control for the fasted Wednesday Friday reading. And so I take the same timepoint Wednesday and Friday, unfasted. In fact, I will eat like the mackerel packed in the olive oil and then go straight to the clinic after. And um unfasted my ApoB was like 73. Okay, which is which is good. Totally normal. Um, when I'm … when I'm fasted, when I'm fasted my first one could be… by the way, I'm already fasted on the first one a little bit, right? Because I don't eat until… I eat until three the day before. If I'm talking about nine, I'm almost 18 hours fasted already. And I already know for multiple, multiple repeats that went unfasted my ApoB goes up, right? Because I'm metabolically flipping. Um, but typically when I'm fasted the first reading, even though I'm fast, it's a little elevated, the second will be even more elevated, okay? And then the third one, when I come back to do the test again and in several weeks, it goes right back to normal. So and our GPT will shows us that that's actually pretty cool. It's actually a pretty cool reading of metabolic health. Um, but when I'm unfasted, I wanted to see… in theory it shouldn't climb, okay? Because I'm uh my metabolism is … I just ate, right? And it actually went down. It went seventy… it went from like 74 to 73. Or 73 to 72. For the second reading. Right? So if anything, it's not supposed to go up and that's what the control shows. And uh the the ratio of ApoB and ApoA doesn't really change. But normally it does. Normally actually the ratio climbs a little bit and it goes right back down. But this time around, and I will say, I think one of my most elite ever ApoB ApoA (ratio)… it might even be the most elite was taken unfasted. Yeah. I think it was like .4 almost down to .39, which is like super elite.

1: Do you like, dig into it like I try to understand why is that the case?

2: Um, part of it is I think they they discuss kind of the ability to process fat quickly, which makes sense, because if I eat the food, take the ApoB, and then I proceed to not eat, it's gonna start climbing pretty quickly, in fact. So it's basically metabolic fat usage efficiency. Yeah. Right? Because if it's not efficient, I take this, I should elevate. If anything should go super high, but it doesn't change. Oh, by the way, I added CRP as well. I have CRP. And the CRP data, in effect has been helpful, but useless. What do I mean by that is I've taken now, I don't know, five, four, six that's even numbers, six CRPs. The first reading I ever got was like 1 to 1.2, which is super low, but at least it was measurable. Uh not awesome. It... But it could have been so that worked out super hard, who knows. But um since then it has been undetectable. It's too… so I got regular CRP, so the reading shows less than 0.6. So I have I have now ordered highly sensitive CRP, but I would say that whether it was fasted or unfasted really at scale is really no measurable impact, I can see for regular CRP. So now I'm gonna tighten it up a little bit and dig a little bit deeper into the highly sensitive.

1: And when you look at this kind of data, can you walk me through like how the interact with your health data, like, does it motivate you to to adhere to your diet?

2: Super excited..., super I'm always very excited to look at this data. I'm checking nonstop, by the way. I … by the way, I'll add I love the process of going to collect the data. It's it's it's streamlined. I'm excited to go do it. It's become a routine now. And when I look at data under different conditions, always super excited. Yeah. Always checking.

3: You mentioned a bit of anxiety, like to checking your data, how has the three meal a day…

2: It's more anxiety, not like nervous of what I'm gonna see, but it's just like, I need to check. I just really need it to get into it. Yes, actually making me want to dig even more. Yeah. And I was and part of this is because I think we're gonna conclude the sleep deep I'm gonna keep recording, but I think it becomes haywire because April, I'm traveling multiple times, may I'm traveling multiple it's gonna be all travel sleep. And I've really got a lot of travel sleep data, so I'm still gonna record it. I'm still gonna try, but I know that I'm nearing the end of the sleep parts. It's been about almost seven months, which qualifies as a longer-term sleep study. And I'm I'm just what but now I think no matter what happens, it's not gonna really change the numbers anymore, right? So we got so much data collected. But um I don't know… part of that anxiety I, you know, I'm eating late into the evening. I don't want this to ruin my good sleep. That's part of what that's what I'm actually more proactively checking it.

1: Because previously, I think in our previous interview, you mentioned that you used data, for example, resting heart rate, HRV [heart rate variability] and you want to incorporate sleep into measuring your overall health. And to measure to do this, you need technology. And I and I guess sleep is like a new thing for you. Like, do you see yourself continuing to track your sleep after this study? Or…

2: I do. I definitely will. Um I think along the way there was a little bit … I actually think about this. I think sometimes there's a little bit of um like the score makes things a little too it makes things a little bit less relaxing. Yeah, like trying to hit this score. And me, I think something we're gonna … my personal goal is to get at least an 80 every night on at least one of the wearables. Right? It's an arbitrary number I came up with, right? Because 85 is optimal for the WHOOP. I don't think it's for me, because of work, et cetera, it's still, I don't think it's totally realistic to get 85 or more every day. I aim for higher, but the goal is eighty, and I think trying that that arbitrary goal kind of sometimes is a burden. But at the same time, I'm also amazed. I'm also amazed because I'm like, I kind of went into this, even though knowing I wanted to sleep well, um I didn't feel like I was stressed every day trying to get 80. Right? And if anything, I just put it I just deposited some data yesterday or the day before of my sleep over time. And you can clearly see on the Garmin. You know, it's like green as you met the sleep need, blue as you didn't. It's like lots of blues intersperse, it's like green green, green, green green. It actually gets better over time. So if anything, I don't think it made me sleep worse. But sometimes I'm kind of like, oh, am I am I too methodical about this? But regardless what it is, apparently it hasn't affected my sleep. Yeah, but I will still keep trying to sleep well and still trying to reach a threshold. Yep.

1: I think this this most of for me to do you have any questions...

3: I know you described it a bit earlier on like how you set the threshold, but do you set thresholds for other biomarker data and like, would you feel it helpful or like, does it motivate you, or the contrary…

2: Ya, I do have other thresholds. ApoB, ApoA ratio was elite and good. Of course they have risk like once you cross this, things are not good, um I do pay attention to all those for sure. Um I had more markers coming up. We were gonna look at the BDNF. Um Those, it's more like I would I would like to be normal. I don't know if there is normal, actually for BDNF, but I would at least like to increase see if fast and can drive it up. So I do want to drive deltas. That's what I want to see happen. In a in a positive trajectory, uh or something that reflects, like metabolic... So, my ApoB going higher is actually not standard. It's not favorable, but it's because I'm switching to fat burn. And then watching it go back down when I take the next set of readings, it is that part's cool. Right? To show that it restores to normal, which is also a reflection of metabolic adaptation. Right? So I do, and I do set um specific goals, right. Like keep trying to keep my … my unfasted ApoB/ApoA below uh, 0.5. You know, uh, keeping triglycerides HDL ratio, below X amount, below point eight or point seven or whatnot. I do keep those goals also. Yeah.

1: And what do you do like, just a follow up. What do you do if you can't hit a goal? Are you going to optimize your routines or…

2: Ya I would. I actually haven't. So thus far, I haven't not met those goals. Uh, but if it gets to a point where, um whether it's the process of aging, et cetera, I will I'll I'll explore prescription medication, um, but I am on none. I'm on none, no prescription medication at present, which we should definitely report. But there's interesting literature, right, saying that at a certain age, keeping markers optimal with some prescription medication low dose, maybe, is potentially still helpful. But I'm not there yet. So if it ever came to a point, I would explore it. If anything, I'm I'm just going a little harder, right? Like my I don't have a specific…I mean, I think for VO2 max, I have a goal of being higher than 50. I don't know what mine is, but we will be able to test it soon. But at the bottom line is even in the absence of it, I do record kind of my… have you guys seen that data yet, my run on the treadmill performance? So I do the Norwegian, three-minute walk, four minutes run, four cycles. And I as I'm running, I'm taking a photo, and you can see the corresponding heart race. I'm not like standing there and not doing anything. And you can see my heart spike up, goes back down on the walk. Spike up, um and I'm trying to record gradual increases in… uh so like last I think two days ago, I ran a … my four runs were like 14.1, 14.5, 14.5, 15.2 (km/h) Right. And before that, so I'm I'm I'm slowly climbing for sure. And my goal is to be able to do it at least one cycle of 16 at four minutes. So I'm getting there. I'm getting there. And I'm also modulating. Like, I'm not, you know, maybe I'll do three at 14, but then do the last one at 15.3. Who knows? Just to kind of keep it interesting and mod. I and I always do that for my exercise. I don't I'm not super methodical like, 14.1, 14.5, 14.5 this. Next one will be 14.2, 14.6, like, I do mess around a little bit here to see if it helps me conserve some energy to blast it out big here. But I'm not gonna like run at 11. And then the next one the last one will be 17. Like, I'm not gonna do that. That might actually hurt me, who knows, right? And so, but you're gonna see this gradual climb. So I have my goals, and no matter what I'm just gonna keep trying to get better and better. If anything, I'm gonna go harder, yes...

1: Just curious I want to pick your brain because you mentioned like quite a few flaws. I mean, you felt annoyed to eat three meals. Are there any pros to having three meals a day?

2: Yeah, that's a great question. And I think to be fair, um when I do my fasting regimen, there are times when I'm a little bit hungry, right? Um, and then there are times where I'm not. I'll give an example. Um you know, there are days where, like, at around five is, sixish, I'm like, oh, you know, I might I could maybe use some food, but then I'm like, nah, like, I'm not gonna sleep well. And I just kind of forget about it. Um, when you're eating three meals a day, I'm like never hungry at the end. I'm which is also not bad… right? I'm always feeling nourished. I just that I think it does weigh me down, does mess with my sleep to some extent. I actually think if I eat anything with salt, 6 o'clock onward, I'm pre- I'm I think it's gonna affect my sleep.

1: So So So… maybe so do you think that it is the timing of consumption that affect more on the number of meal? So you maybe you can have…

2: I think it is timing, because I, the number of meals in the end I don’t think it’s a… the amount of food, I don't think is a huge thing. The annoyance is the act of having to do it. You know, uh that part, it's not I don't I do not think eating three meals is bad. If people are heavily endurance driven, I think they should watch out, in fact, and not like over fast. Um, I think it's just part of like scheduling food and it's become it's actually a way of life, eat here eat here, you eat here. I'm like, if I actually just eat around this time, I'm done. Right? And um oh, I will add uh, my as my data will show, I can spontaneously enter ketosis when I'm in my OMAD. My first fully recorded three meal days zero. Okay, zero ketosis, which to the layperson actually might be interesting. For me, it's like not that surprising, because I but but the thing is I do eat carbs on my OMAD and I eat carbs on that on this the three meals a day, but zero like not even close, which not that surprising. Even when I'm doing my OMAD, and eating low carb, you're still gonna get a drop in your ketones. But I will say this. I took a cup and I I sent this to Peter. During I think it was Wednesday or Thursday of last week. I happened to take my uh in it was Wednesday. I had a lunch. I had a lunch scheduled. Yes, Wednesday, I ate lunch at WHO. I went and went off site to eat lunch, came back towards the tail end of lunch here, grabbed a plate of the chicken and the broccoli quinoa, which is that was a little sweet, by the way, maybe some soy sauce, there's soy and before I had dinner, because I was collecting blood every day last week for the ketones. Before I had dinner, I I sh I went home and I was 0.5. And I was like, what the heck? And I just ate like a few hours ago. So I took it again and it was 0.8. Then I took it again, it was 0.5. Okay, so I'm like, hmm. You know, did I and I sent to me, I'm like, Peter, what's going on? And to be fair, that week was nuts. Right? That I was running around all over the place, cognitively very involved. I was still working out heavily that day. Um you never know. And I've actually I consulted ChatGPT like, is there a scenario I could totally flip within even a few hours and it's possible, right? But then quickly the and I was like … and I told Peter, there was a weirdness of the sensor the following day, but then I ended up it ended up being, you know, I think it settled back. So, but I took three in a row, right? And then the next day, no ketosis again, and the day after that, no ketosis again. And so I will say by and large the three meals a day at least for me does prevent me at scale from metabolically flipping, however. And that's not something I I want. I I kind of view it as metabolic exercise. Right? And uh that's not something I I willing to sustain. The lack of the switch Yeah. Yeah.

1: And I just caught it, you asked ChatGPT about it for for advice about your data. So maybe like, how has to ChatGPT or Gen AI in general, like affect the way you interact with…

2: So you know, I don't consult it for like major medical decisions, but when it comes to its opinions on, hey, you know, today I did a grip strength of this. How does that look for someone my age? I metabolically flipped why? I actually like I like having some insight and it's secured by the fact that I'm not using it interventionally right? I don't go “GPT tell me what to do.”. I do it as a checkpoint. And I think that is super cool because the questions that I ask it may not be readily answerable by conventional staff that's the conventional clinic visits. You know, um and so I think having that feedback longitudinally keeps me excited. You know, GPT, I metabolically flip zero to keto. Right, zero to keto in 16 hours. What does that mean? You know, and I'm I'm mindful to not lead GPT, right? Um, and it's it's pretty interesting to see the feedback that it gives. Is there some… and we we're looking at variability between the same questions, et cetera, or throwing in confounders? But by and large, um it's it's I find it as fun just to have something to get some feedback on. Because especially because it's health related. Yeah… And by the way, it it does amass over time, right? So I'll be like, uh, my grip strength was 79. Whatever kilograms. How does that rank for 45-year-olds? And it'll come back and be like, oh, this rank says elite, very athletic. And by the way, on top of your ApoB ratio of this and this, uh and this data, that further as so it remembers, right? And then the other component is, um I one time I was like, hey, my fasted ApoB is this, my unfasted is this. Ratio, unfasted, ratio fasted, can draw comparison what this means for me. And it totally drew it out and gave really good insights with references that were applicable. I I think when it comes to preventive health, and even offering some of these insights to and I I still work with standard medical professionals who are great as well, it's mutually educational. Right? So I think it's actually fun. And I think it actually is one element that keeps me going. You know?

1: I think I'm done.

2: Other questions?

1: I don't have any more. Do you?

3: I… one weird one. Do you miss the feeling of being hungry when you are on three meals a day?

2: Uh, that's a good question. No, that's a good question. I don't think so. I think it does provide some but, you know, I think it provides it eliminates one discomfort, but I think it adds more other discomfort. You know, like I don't like eating in the evening. Like I I just don't feel good when I'm eating late into the evening. And for me again, late is like six or seven. I just don't like it. I I did it before, but I think I've adapted it. And I've always felt that eating midday was much more conducive for me. I will exercise in the morning, so I get some food shortly after. And then once I eat really well now, I don't have to eat for the rest of the day. You know, even on vacations or on work trips, even though because of necessity, I may have maybe have to eat a little bit later, um I will still kind of compact the meals together to try to get it all done within a certain window. And by the way, for my trip, which preceded the three meal a day week, I don't think I ate in the evening once on that trip. Yeah, zero. In fact, I went with our colleagues to get food and I was I actually thought about eating, very accomplished meetings. I thought about eating and I got there and I was like, you know what? I'd just be eating for the sake of eating. I actually ate very well for lunch. And I sat with them as they were eating we got takeout, we had some more meetings to do, and I just sat there, they ate I was fine, they didn't mind, they know what I'm doing. And so I actually didn't eat at all. And my sleep was phenomenal every day. Worked out every day. You know, uh, it was great.

3: Would you say it's the time between, like your meal to sleep instead of like the the fact that eating?

2: Yeah, yeah, I think the amount of well…

3: The time frame you get to digest…

2: Exactly. Right. So I think the amount of food, it was kind of annoying that had to do it, but, you know, maybe not so hungry was good, but it was the kind of the timing and how it affects my sleep, that I'm not okay with. Because it affects my sleep, um it may affect my performance, but I don't know. I might mentally be like, you know what? I'm gonna go hard anyways, who knows? You know, but if it was like an objective marker, if I was testing VO_2_ Max as a function of eating every night for three months and then not eating, I I would estimate maybe there could be a difference or run time. But, yeah, it's really the time eating before sleep is probably the most, yeah, non-negotiable.

3: Thank you.

1: I just have a follow up question that I thought about. It is like because like normally after you eat a meal, you'll feel a bit full and you feel like a bit tired. Or full coma, in a sense. Do you feel like having three meals a day, like makes you experience three food coma, affects your performance or…

2: Not really, because I'm still eating whole foods, like healthy food, so probably not, but I will tell you if I was eating like French fries and stuff. I definitely would not feel good. I would definitely, definitely not feel good. Yeah, and I still brought food, like, I think the second day of the WHO meeting, I brought a bunch of food, um third day I I specific because I was going out for a meal. I picked a Mediterranean restaurant, and I was even though I ate one plate when I came back to make sure I had enough calories, it was chicken and veggie. Maybe had a little bit of marinade, but it was still quite healthy. I still made sure to bring the chia and the flax, et cetera. Right, and today I have a tight eating window. I have a tight eating window, and so I actually packed I'm gonna order food. I have just make sure I get enough the healthy fat. I've got my macadamia nuts, healthy fat. This is a mix of chia seed Psyllium husk and flax seed [point to his food]. I'm gonna go buy like a zero-calorie drink and dump this in there and and get some of the fiber and then into my vitamins. Supplements. Yeah.

1: That's a lot.

2: And like I could put this in water, but that would not be pleasant.

3: Will not taste as nice?

2: It would be gross. And this [Psyllium] should not be put into like, just taken, because Psyllium absorbs a lot of water. And people have been able to have difficulty swallowing should not be just thrown into the mouth... Chia it's it's disgusting if you just throw it in, super dried, but Psyllium has way more fiber content than chia. Yeah. And so today, because I'm gonna be podcasting and then I'm gonna order my lunch from there, I'm gonna eat… unfortunately for the people who have to meet with me later, they're gonna watch me eat, but then I'll be done. I'll be done by about three yep. Yeah. Other questions. We still got some time.

1: I don't have any questions.

2: Yeah. All good?

3: Thank you for your time.

2: Thank you. I guess we're gonna do another one, right?

1: The last one.

2: Okay, yeah. Um just for timing…

(End of Interview 2)

DELTA001 Health Optimization

Interview 3

**Administration timepoint:** End of Study

**Interview date:** 6 May 2025

**Recording method:** Audio

**Transcription style:** Verbatim

1: So this will be our last interview.

2: Okay!

1: And before we start, maybe I can just go through like why we are doing this interview to better scope the responses for this session. So, like, because you have undergone um a series of meal regimes.

2: Yes.

1: And we would like to learn for your insights from your daily routines. For example, diet um exercise and your use of technology or any other aspect that you'll love to share for healthy living to lead a healthier life.

2: Yup you know if I think about the whole experience, there's a few things that I tried to keep consistent as consistent as possible. Um that I I view as pillars of the whole thing. If we start with diet, it was that if I look at the numbers I I fasted I think could consistently for at least 18 hours for I would say greater than 90 plus percent of the time I did at least 18 hours. I could arguably say I probably did at least 20 hours if you look specifically, I think a bulk of that was at least 20 umm the 48 hour fast were not were about were clearly a part of this regimen as well. Um I don't know if everybody has to do those, but I would say what those durations do for your biomarkers to see if you can stress test them to rebound back I found fascinated. So fasting was probably a pillar of this regimen. Dietary because there's a tendency to have so much variability. I tried went on the ground in Singapore, I tried to keep certain things absolutely consistent. So I would say 99% of the time when I was in Singapore, assuming I didn't have to be away from being able to bring my own food or being able to to eat at home, um had I would say three things were super consistent. One was this kind of delta delta drink, delta shake, if you call it. Which was blueberry, a green, typically broccoli, red cabbage and ginger, blended with water, not juice, but blended egg all the pulp. Everything consumed. And I generally hadn't I documented that in the photos of the food. You'll see, it's recurring. Even if you do an image analysis with ChatGPT, it'll catch that this is a recurring thing. So this the drink. The second one is trying to get chia, flax psyllium, um every day, right, from almond milk pudding, every single day I was here. And I had cases where I couldn't bring it to uh I was in meetings running around town, so I'd actually bring a little plastic bag of those two, I was just buying something to drink and pour it in there and just drink it. Okay, so the chia… so the fiber, the chia, the omega three component, that was that was to address omega threes. And then third is some sort of additional veggie, green, whether green or red, either red cabbage or kale. Right? A separate bowl, olive oil, chili powdered and salt. Right? As as often as I can. And it's it was to a point where there's some images in my food, where I actually open a bag of kale and I just eat the kale out of the bag, because I don't have time to bring it so I don't have time to bring olive oil, salt, and I would just straight up keep the kale out of the bag. All right, so those three items I tried to keep as consistent as possible throughout the whole duration. And on rare occasions I was even able to get pieces of it, deconstructed even while on travel. Okay, um and then finally, uh two things, fitness and sleep, moving every day was a priority. Um strength training, I would say more or less every day. I would lift weights, I would do dips at the airport, I did dips in airplane seats, I've done all you can imagine. Um even in airports, I will go for walks in the airport. And so moving every day was a part of this, and then sleep, um, as you know the sleep portion is over, but I still prioritize the sleep. Um getting to bed on weekdays by 9:15 is still a priority, waking up at about six. Um I have started scaling back on the wearables. Like I said, I would uh, but I'm open to even trying more. Um as as I continue, but sleep always a priority. And on travel, sleep is also a priority, whether it's leisure or work, always a priority. And I still time my flight. When I fly, um all of the above. So all of those are the key pillars.

1: Okay. I have questions on sleep but before that, maybe like have you discovered or noticed any new challenges towards each of these pillars? To adhere to it? I mean to when you change the change your meal regimes are there any new challenges that you encountered?

2: No, I find that like not falling off the wagon at this stage has… not falling off the wagon, so sticking with it has been easier than I imagine it would be. So I in terms of challenges um I think I have become quite accustomed more accustomed than I thought I would be right. So I regularly and at events where there's like pizza, surrounded by pizza, surrounded by french fries, surrounded by everything, and I'm quite good at just, um avoiding it. One thing that I was I'm grateful for through this is the people I'm around, because I still socialize. The people I'm around have become quite have always actually been accepting of it, right? They they I'm I'm happy they enjoy what they want to enjoy. They don't and I'm totally okay either being there, not eating, or eating all veggie uh if if need be. In terms of sleep, I would say, uh I'm I'm totally okay sleeping at 9:15 every day. Um, I wouldn't mind pushing it back a little bit to ten PM. Uh, just because, um I don't know if I really have to sleep at 9:15 every day to get good sleep. I I've done 10 PMs before on a weekend. I've had to do it because of travel. And I sleep is still good. You know, I don't think it affects, um my sleep, but I will always so so I would say there might be an adjustment to be made there. Um exercise, um I would say the HIT training is is more challenging to sustain just because I I do have to think about not getting injured over time. And so I I think I'm gonna need to prioritize more recovery days, that that could include walking, that could include stationary biking, but I think doing like three days of hit every week constantly, I think, um, even with a week off, I think I might need a little bit more recovery. The recovery may not be taking two weeks off, but it might be like taking a week off and then returning with maybe a few days of easier cardio to keep the aerobic base going, but doing constant HIT training is pretty punishing, even on a treadmill.

1: How do measure your recovery because…?

2: I I don't completely. I think I do kind of impromptu monitoring of heart rate recovery. Uh but I think that ended up itself is not a full reflection of performance recovery. You know, there are d- and I don't know if the wearable recovery is a true indicate... Like there are days when I feel really good at the right away, wake up and the recovery is green, like really good, and I think it aligns. There are days when my recoveries yellow and I'm like, I don't know what to make of this and I get a really good I feel really good on the HIT. I think days when it shows red, as long as there's no discrepancy, I think those are believable. But wearable based recovery, I don't place a huge emphasis on, but um I do on my own taper? [9:16] back some of the HIT training, just knowing like I did four straight weeks of heavy HIT, I'm definitely going to give myself a week off. Yeah.

1: And um when you mentioned about your diet, I remember in our first interview you, imagine like a healthier version of yourself consuming more plant based, uh protein. Have you …?

2: That has actually… I would say it's escalated. I don't I don't always do like today's plant only day, for example, but um if I kind of just survey what I've been eating over the past… I I would estimate there is a trend towards more plant-based. I would say chickpea purchasing in my household has gone up a lot. And uh and I'm going to start I've already bought lentil, right? I'm starting to expand kind of the more plant-based approaches. Um I I think that will continue to grow slowly.

1: And um have identified any changes that you want to make moving forward, like to your diet?

2: Umm I actually might so I've been quite, I would say restrictive about red meat. I've been quite restrictive. Um There's been literature about red meat, maybe being a contributor to higher homocysteine levels. But I I now I'm in the past few weeks, I've been able to look at my homocysteine. I've had a little bit of red meat. Um, even before readings and it doesn't seem too negatively impact, because I think I do take B vitamin seriously, plant-based, seriously, not over consuming meat-based protein seriously. Um, so I might actually at least allow myself a little bit more red meat, maybe I don't even I don't think I have red meat once every two weeks, even. Maybe it's once every three weeks. Maybe it's once a month. Okay. Maybe I'll allow myself twice a month. It's a little bit more red meat, especially because um I don't really monitor iron levels much. I think my energy is fine, but maybe a little bit more red meat, but but I'm not certain about that. I probably will start I' actually started noticing I'm reducing chicken even a little bit more and upping more fish. And I think that will be another trend.

1: Okay…

3: Uh, speaking of all that your shift in your diet, how have you found, like have you found it changing like, uh your biomarkers or like your perception of your energy levels and stuff? Since you have shifted from … towards what are more plant-based diet?

2: I would say I think my energy is level very high. I think it's very high, so I yeah, I I I don't think the plant-based has certainly I don't know if it's made it better anything, but I think it hasn't negatively impacted energy. Um and I in terms of biomarkers, I don't know if the food has improved it. But I could pro(bably) I I can say this. Um for the unfasted ApoB readings. Um I had definitively opened cans of mackerel. It drenched in extra virgin olive oil. Eaten it during the three meal day weeks, that's when I do the unfasted. I've eaten it and and gone straight to the clinic. Okay, we're talking about the time between uh food to blood being taken is maybe 30 minutes. Okay, truly unfasted, and some of my very, very best biomarker readings came from that., right? And so I would say that uh I have a lot of a piece of mind knowing that healthy fat consumption, um taking blood mark… biomarker readings that are okay to be taken unfasted, and taking them clearly unfasted, with no fear of skewing of markers in the wrong way, et cetera, has made even more confident about the the items I picked.

1: So…, sorry.

2: No worries.

1: Um, I hear that you you are taking like blood test on homocysteine, ApoB... Because you have been taking like such a comprehensive, like an extensive health test. And you mentioned just now that after reading looking at your homocysteine levels you think that you can maybe consume a bit more red meat in in a sense, it doesn't really affect it. How has this kind of like health test affect your daily routine? Does it influence your decision making?

2: It does, I think I think this is important to note, which is the average person is not part of a funded study. Right?

1: Yeah.

2: Um, fairly speaking like, if somebody's is like Dean, we're gonna give you X amount of dollars, we know you're the subject that's been disclosed, there's an IRB. I think it's less likely that I'm gonna slack off. Okay, that that's the logistical component. Knowing, like that there is uh, you know, competitive funding that has been set aside. I think that, to be fair, influences behavior. And you know, but you know what? It's and if you add to if we if we kind of devil’s advocate, right, someone say, you're it's totally unfair because you're funded, you have a responsibility and I'm actually I am also the faculty member, so I think there are multiple responsibilities, hats, affiliations, et cetera, that make it more compelling for me to stick with it. At the same time, we do know that even paying people who participated in other studies, not me, but others, is not enough to drive better behavior. But for me, knowing that there's a funded study overseeing this does, I think, influence my behavior to stick with it. Now to go down to the tests themselves, the tests as an insight into my health, I think influenced my behavior. Absolutely. If you just look straight objectively at and I'll give you an example, tomorrow, I'm gonna go in for another homocysteine that's kind of what the baseline homocysteine control. And um I would say even though I have no plans to go to, you know, fall off the wagon today, um knowing that I have a test the next day makes it even more if I if I were the person to think about having a bag of chips today, which I don't eat. Okay, but I would I would be careful. I think that having deep, sustained insight into my markers is a gate for me. To keep me on the edge and I can confidently say that when the marker portion is done, um, I will be very mindful, because I have enough insight into what can be affected by what I do and knowledge of beyond like, oh, you don't want high homocysteine. I get it. You want you want a below but… knowing that homocysteine levels can impact cognitive health long term, metabolic health, heart health, and seeing the variations that occur over time, like, I'm quite mindful.

1: Like you mentioned, accessibility and financial respects can be a challenge for average person I mean, to do such an ex expensive test. And if you … in your opinion like, what, the other practical values, such testing brings…?

2: Ya, you th... Okay I think the key thing is you don't need, like, homocysteine, ApoB ApoA [inaudible: 17:52] of your readings to the frequency that we've done, we don't need that. I think to offer a periodic insight into flexibility, into metabolic and agility, I think it is valuable. But doing it an N of 3 N of five constantly over time, not necessary. Um, but I think having some insight into homocysteine status, for me, I something that I didn't realize would come out of this was how it would prioritize my B vitamin consumption. I don't I'll never stop ensuring that I take enough vitamin B now. Right? Looking at the homocysteine marker and the role of vitamin B deficiency, and by the way, aside from homocysteine level, vitamin B is for hemoglobin. I fast. I think it might matter, right, to make sure I'm I'm looking out for that. I think just even a little bit of insight into homocysteine will has will has changed me forever. Vitamin B paying attention that will always matter. Not over consuming red meat. It's made me look into what types of meat, have what levels of methionine, et cetera, right? To compare between fruits, between nuts, et cetera, um, I won't need to do homocysteine five times three times a week forever to care. But having this a little bit of a pocket of learning, a little bit more not just about what my single baseline is, but how we can fluctuate as a function of my behavior, and then maybe not fluctuate as a function of my behavior uh will be like lifelong insight.

1: And apart from homocysteine level, are there any other tests that have influence daily routines?

2: I would say ApoB dynamics I never real … like I don't get super excited when I see a good homocysteine baseline. Like I got a baseline reading yesterday. And it was it was great, right and I was like this is a means to an end. I need to know Wednesdays. If I get especially the homocysteine intervention arm, which is like pre fast, post fast recovery, that first reading for me is merely like the appetizer, right? And I think that anything with dynamics is what for me mattered the most. Because I don't think we can really determine what is optimal for us until we stress test a little bit and see is our system able to respond to perturbation. Right? I think it's getting one reading if you it's about depth, right? Yesterday, my reading was great. It was ultra low risk. But if I hadn't known that if I had fasted that it would do this, right, jump up and that it would take this long for it to come back down. Or to realize that a second time I fast it doesn't go up at all. I goes down more. You won't know any of that. And it's it's like looking at the surface of a pond and not even taking the time to look deeper inside of what's going on. Right? And I think that while you don't have to do it forever, by any means, periodic looking at dynamics is is important.

1: What about the experience, user experience of this test in general, how do you feel?

2: Yeah, I I've never been afraid of needles. I don't think anybody's loved getting jabbed, but I would say that I'm even less like mindful of getting jabbed now. You know, um I think the experience has made me view kind of lab testing as something I really look forward to. Yeah. I I'm excited. It's it's like just some it's like you know, uh it's like I'm gonna go get something really interesting… kind of uh obtained from my body today. And that's I found it to be super cool. And, you know, I think that lab testing is institutionally been viewed as infrequent, for sure, because there's a cost involved in voluntary nonfunded, right? Um, it can also be viewed as very scary. It can be viewed as, um not even a thought in people's minds also. But for me, like, I would love to be able to have something like this be a part of my life, not every week by any means, but um sustained. I I with proper means, I would probably continue this, even just for my own info, not even under study. Right? I have to it's gotta we gotta keep within reason. It's it's it's elective, but I would say the process became very easy.

1: And if you are not funded, um what are the essentials health tests that you you will do regardless?

2: Yeah, I would probably do like HOMA-IR…

1: What is that?

2: It is as an insulin resistance test. I've already consulted with a number of clinicians informally about my existing data to the point where they're like, yeah, you don't shouldn't be spending your own money doing this repeatedly, but, you know, periodically probably a good idea. Just as a little window into my ins(ulin) we are Asian after all, uh it's good to keep an eye on it. I might look at ApoB more times over time. Nothing like not too much on the dynamic side, cause dynamics in the end if I really wanted to, I would just fingerprick with ketone. Because I could look at flexibility that way and I could just do that at home. And I find that, um you know, if I really want to look at metabolic flexibility and metabolic health, et cetera, um I can still learn a lot, right? If I'm consuming too much carbs [carbohydrate], too much sugar, it's gonna be really hard to flip, if I consume too much carbs, too much sugar, I could estimate some of my cardiac markers will get worse. And so the ketone I will never stop.

1: I am curious because you mentioned about flexibility, metabolic switching, how how does knowing your metabolic flexibility helps your lead a healthier your lifestyle?

2: Yeah, I think if flexibility is too slow it's it's a clear indicator that I'm consuming too many carbs. Okay? Because if my body needs to clear all that out before it flips into ketosis. Umm… and from a dietary standpoint, it's a clear indicator. We know that, um metabolic flexibility could be an indirect, not fully established marker for biological age, metabolic age, et cetera. And the time it takes to flex in and the speed keto tests et cetera. I think will always be a good marker for metabolic health in general, and the longer it takes to get in, if I start to see a slowdown, right? I think that'll be telling for me, like, in my becoming lax in how I eat.? Am I sleeping more poorly? That's that's why I feel in the insulin component part. We think of poor sleep as poor energy, but it's got so much other, right? Cardiac health, resting heart rate. Uh maybe cholesterol, sure, you know, so, um I think that being able to look at that agility is always gonna be a good marker.

1: And speaking on sleep, you you mentioned that… sorry, let me rephrase my question. Speaking on sleep, most people… Okay, I'm not going in generalized, but um sleep quality can be uh assessed through, like that duration of sleep etcetera. Like you just mentioned that metabolic flexibility and stuff might have some indication on your quality of sleep. Personally, how do you…

3: Experience?

1: Look at… how do you define what's a good sleep?

2: Ohh, that's a good question. So I don't think I have seen the only things I have noticed are that back in the day, okay, when this is… I am tying this to metabolic, but imma go back to sleep on sleep alone. Tying sleep to metabolic health, when I first started doing the longer fasts, that that night where I kind of don't from the full day of not eating, sleep can be pretty rough. It can be interrupt, and that's well known, is that sleep interruption can occur, but eventually it came to a point where I felt sleep was… I'm fine. I sleep like any other day now. So I think there was some sort of adaptation there. To being able to sleep wall fast. I I don't know if that's surprising, but I did go through that. Um, in terms of assessing whether sleep is good, um, I I feel like I know when I wake up if I am well rested, right? Just, you know, how quickly I jump up to, you know, am I staying in bed for an extra two to three minutes? I would say I never stay in bed for more than like two minutes in general. I probably… the first thing I do is I um turn on and I check for my out my sleep outcomes. I wouldn't be surprised and I'm not making any determinations here, but I wouldn't be surprised if somebody was able to uh assess my keyboard agility, my my agility at being able to pull up because for whoop, you have to you have to open the app. It asks for a daily journal entry, which I close. I don't do a journal entry and then you get hit process. It'll process your sleep. Right? And then I quickly I used to have a Garmin on, so I would quickly switch and turn wake up, you know, end morning, something or end something and then to access. I'd be willing to bet that my sleep quality can be assessed by how fast I'm able to get all these tasks done. Okay. You know, that that doesn't surprise me. Some days I'm probably like hitting the wrong hidden you know, missing the button, but I think that, um that would be a cool way to assess. I think I'm I just know if I'm like really awake.

1: [whisper] Awakeness after sleep onset.

2: All the way to like I keep my phone relatively equidistant how fast it takes to turn off the alarm. I'll bet you there's a correlation to all of that.

1: There is a structured way to do it uh. But but coming back sleep, um because of you you you do have your own sleep hygiene practices, uh not consuming food after certain period of time, et cetera. And during this study, you do have to consume like three meals a day and not at some point of time. Did it really affect like your sleep?

2: You know, I thought it would I I would estimate in the end it didn't, do much, because one thing is I I don't if you look at even my weight, which I don't remember, so it's good. It's gotten quite blinded to it, even though I have recorded all of it. I didn't I don't think I overate during the three meal a day. I didn't gain substantial weight, maybe in some sometimes I didn't gain any weight. I think I really tried to portion control it to the point where there was some sort of relative consistency of overall calories., even with the three meals of day part, I made it very clear not to eat late. I'll get the three meals, but I'm gonna try to finish eating by 6(pm) or 7(pm) at the latest, because that's that's a couple hours before sleep.

1: Yeah.

2: Yeah.

1: And um because you you do travel and I know there are like some strategies out there that you can adopt. So that you can switch you can adapt to the jet lag um more efficiently or smooth smoother. Are there any strategies you adopt for that?

2: So I like taking night flights. Yeah. If I take night flights, I would never eat on takeoff. I think it's for me, it's a recipe for very poor sleep on the flight. Right? Because eating a food, sitting down and going straight to sleep and around at a 11 PM flight, I think it's speaks for itself. Right? At knowing that that that the only how matter how long the flight is, you only have this much time to get you rested. Right? And a lot of these flights I take you will land in the morning. Right? And so, the last thing you want to do is go into any morning with poor sleep. Right? Um, I so and I might have my now habit is put on the eye shade, eye patch, earplugs and just go straight to sleep. All right, um I would say, and this is documented, I have not used any prescription medication at all. And this is not to say that others shouldn’t, but I did not, because I didn't, I wanted to really try to understand if I could I could do it. Right? Okay, other habits are compacting meals together, so I skip the take-off meal. I will combine the meal that I didn't eat with the landing meal. Okay, so I'll literally I didn't so if I basically take a night flight I had been eaten for most, I… after my lunch, I haven't eaten most of the day, get on the plane, go to sleep, and then when I'm about to, when it's about time to land, I will ask to take all this food, get to where I'm going, if I need to eat a little bit more, so it'll be a little earlier than usual, but it's kind of pick all your meals together and just finish out the rest of the day, which is exactly what I'm doing tomorrow. Right? And so it's about still maintaining the time restricted eating and um if there is continued flying through the day to get to a destination, no sleeping, on those. Because I should have slept well that first path in. And I think the Portugal flight will be a good example of that. Um, so and then on travel, um I at I'll typically if it's leisure, still 18-6, typically, maybe 16-8, but I cannot remember a time that I did like three meals a day, on travel. But the sleep happens … oh, I will add, if I have to take a daytime flight, that is long… that's a that's a long haul. What I will do is I will wake up a little earlier than usual and I will exercise super hard. Okay, so that I can be extra tired earlier to get and we'll do is all if the flight is at 10, 10 AM, 12 PM, I will eat. I'll actually eat every meal that I take off in this case. Both meals moved up front, because it's like lunchtime. And then I will kind of do some motion. I'll do crunches, I'll do planks if I can, you know. Um safely compliant with, you know, uh air safety guidelines, right?

1: On the plane?

2: I've done plank. I'll do crunches in the I'll do crunches, no problem.

1: Okay.

2: Yeah, I've done it. I'll do dips if I can, if there's space, I'll do dips... I will get my standing hours, right? I'll go to the bathroom, like, nonstop, if I can, just again, under safe uh, you know, seatbelt sign is off. Um and then I will I'll I'll sleep about four hours into the flight, just to try I'll aim for seven hours on any long haul, I will aim for seven hours. No matter how I do it, whether it's sleep immediately or whether it's work out really hard in the morning and then um, so I have extra tired. To be able to sleep earlier.

1: Okay. And and are these strategies effective…?

2: I think they are effective, I think they are effective, if we continue to look at the sleep… I think they're effective. Like my sleep dur(ing) like the the flights, the flight sleep is never gonna be amazing although I've had I have had a couple that were quite good. Um but if you look at maybe even one night, even if it's a red eye that's 5 hours, I think you'll quickly see that my remaining sleep is quite good, on travel and even for work related travel, um, I think I've slept really well.

1: Okay.

2: Yeah.

1: Oh, just...

2: Oh I will add, I don't think people everybody will do this. I don't think everybody will gamify sleep duration, push meals together, et cetera, nor do they have to, but I think that to kind of stay energetic, to really kind of not I think it's important to try something if possible, like not doing anything is it can be you're talking about losing a week or two of your wellbeing. Yeah.

1: Just to be clear, I want to ask that for your flights to usually take business class or economy?

2: I mostly take business class for long haul.

1: Okay.

2: Uh for short haul I take economy.

1: Okay.

2: Um and if it's but for the short hauls in economy, I will I because it's a short flight I'll typically specifically pick midday flights. There's a reason for if possible the reason is midday flights, what I can do is I can actually eat before the flight to get my calories in, eat on the flight, which are almost exclusively low fat meal, by the way. I ordered low fat, which are documented, and then I don't eat for the rest of the day when I land. So I I picked the mid days because it allows me to totally stay in my routine. No sleep. I do not sleep on short hauls. I don't fall asleep at all.

1: Good because I was, I was wondering like because, might be unrelated, like the economy seats are tighter.

2: Its tough!

1: Like do you, how do exercise. Are there…?

2: But economy it’s is likely gonna be short haul. I had exercised already.

1: So there's not like strategies on the seats?

2: I will still, I will still do my stand if I can. I will still keep mind of being able to to stand and stretch. Yeah. At a minimum stretching out on a plane is important.

1: And the aisle seat is the better seat in that case, right?

2: Yeah, yeah, yeah, definitely.

1: Do you have any questions? Okay, that sorry? [question directed to 3] Yeah. So, uh now I would like to move on to use of technology. Had… do you will notice any change in the way you interact with your wearables or any technology?

2: Erm… Oh, let me just my lunch person is uh yes…

2: Uh, in engaging with technology as in I would say I like it even more… during the course of the process. I actually love checking the data. Uh I would say there were moments where I was maybe a little stressed out by like some sleep scores, maybe after travel, where I'm getting the hours, but the sleep scores aren't that good, could be a recovery thing, but by and large, it's minor. Like I do really enjoy using using them.

1: How do you manage those stress, when you receive a bad result?

2: I I just try to make sure to assume and usually it's I had no deviation. It's it's usually I'm back from long haul. Um I would say that it is possible that some of the sleep scores take time to recover is because I actually had if you look at my San Francisco trip, I think I adapted super well to the time zone there. Okay, and I'll give you I think I should mention this. That one, it was a night flight outbound from Singapore. Slept okay. Challenging because I slept uh, I slept in the flight, woke up for a few hours to eat, but then I land in the evening, not in the morning. And so the real problem is you land in the evening, you're like I'm not tired. I'm gonna stay up till 4 in the morning. That's the problem. And I would never do that. So what did I do? I got to the hotel then I I went straight to the gym, and I did a brutal, brutal HIT regimen. Okay, to date it was the hardest HIT regimen I can remember. It was very, very hard, okay? And I slept fine. And in fact, I I would argue I remembered definitively I had days there were, slept all the way through to the alarm, woke up, it felt amazing. Right? And sweeping through the alarm, like to the alarm, not through it, but to the alarm is always a great day. Right? And I if you can even look, the switch was clean. And the switch back was clean, but for duration, I would say, but I think the recovery, some of the wearables, recovery matters heart rate, I would it could very well be that I slept the time of my heart rate was high. Right? And so for me, I didn't do anything different, because I I knew I was sticking to the plan, you know? Yeah.

1: And and for technology-wise I identify any new markers or digital biomarkers that you're interested in throughout the course of the study?

2: Ummmm I would say I became very not new, but um I became very mindful of uh resting heart rate. And and the kind of that evolution over time or what it was. I think that's something that um and the impact that even one good night of sleep can have on resting heart rate. I think there's I'm always interested in markers that are that can change off of one day. Quick wins or quick losses.

1: And how how do you incorporate like tracking this resting heartrate into…?

2: That’s’… it's probably the that's the second thing I check every day on every. First thing is probably the score. Second is maybe duration. And third is what my resting heart rate is.

1: Do gamify let's say the results your markers, results of your markers to the routine.

2: I don't that I know of. All I know is I just want to keep my resting heart rate as as low as I can get. I I'd say one maybe gamification is if I notice any creep up uh actually to be fair. If I notice any creep up of RHR, um I might step up the uh the non-recovery cardio, which could be more zone two or more HIT. To frankly make myself maybe a little bit more tired to make sure I'm not under strain so that my sleep is not kind of I'm not really ready to go to sleep.

1: I'm just curiously, has any of these wearables or technology helps you adhere to the daily routine better?

2: Yeah, I think that would be fair. If I it's I think it's positive reinforcement more, right? Um, like when I sleep well, right, it kind of reinforces for me like sticking with this regimen is good for you. You know, and and the wearables do encourage that too, right? They'll offer nudges like your sleep consistency is really good. Try to get to sleep at this time today if you wanna keep that consistency up. Um uh you know, watching the HRV go up watching the RHR go down, for me is a total positive reinforcement. So to keep things up. Um I would say that uh not waking so I would say another notable one is making sure that I don't wake up at night. I really try to not drink water within three hours of sleep. I really like not having I really like keeping the awake moments low/sleep efficiency high. And I think I will say this too. I still wake up to go to the rest room. But I will always be able to go back to sleep in it. I would say that for the wearables, if if I were to say wake up to a bathroom, lay down in bed, surf my phone for a little bit, every wearable will will go hard on me. And then that's you'll they'll capture all of it, including the apple. Okay, which I believe certainly is generous with how it counts sleep, but it will catch it, for sure. And I have been really heartened and and I don't maybe be surprised by the fact that, like I will go back if I go back and I just lay down and just stay calm, which I always do, I will go back to sleep. And I'll tell you, sometimes I I've go to the rest is only an hour left before I have to wake up. I'll still go back to sleep. Sometimes I'm a little up earlier in the night. I will I'll definitely go back to sleep. And I just remain calm. Nothing's like no expectations. Oh, I woke up earlier than I thought, bam, I go back to sleep. And I would say every so often, I had one where I to go all the way through to the alarm.

1: And because you you are wearing that multiple wearables, you have results coming in from different wearables. How do integrate these results?

2: I only I kind of um I kind of just like to look at I like to look at concordance, but it's not it's not really kind of like I don't go oh my they're so different from each other. I'm actually just more interested in why. Yeah, I don't blame a wearable. This one gave me 90, this one gave me 85. I believe the 90. I would never do that. But I would say I believe that their ability to kind of track general trends. I think they're all quite good.

1: And speaking on trend, it feels that you you would love to check the changes, the trends. On the user interface, aspects, and delivering of health insights. Do you have any perspective on…?

2: I think having easy access to the to the actual trajectory, because it's not always the case, right? I think it's better now. In fact, it's it's… I think the wearables have done maybe even more effective than conventional healthcare. I think that making sure that there's quick access to because every day you're gonna get a score. Every day you're gonna get HRV, RHR. Like as a as a straight up number. The easier you can, if it's not readily available, the easier you can access the trend. Which I think apple is quite good at. Okay, and being able to see the trend over time, I think matters. I think people want to see more of that. Some of them are really good at this huge score right on the screen. Like if I can quickly get too well, how about the last five days, last ten days, last 12 days? I think that would help a lot. Because some of it is a score in an arrow, like, you went up from yesterday. That's good too, by the way, day to day, but kind of easy access to trends, important.

1: And on a broader perspective, because these health care data, is collected by the user itself, how how do you feel like it can be incorporated into healthcare industry as a whole like, hospital...

2: Erm… I think, yeah, I mean I think that being able, with consent, to port this into a certain records would be huge. Giving an example. Um I had an instance where I was with the Garmin specifically, and I was um I was on a holiday with the family and all of them caught a pretty bad cold. Okay, um and I was housed with them to the point I'm like, well, I'm next. But as my records will show, I not only didn't get sick, I actually ended up working out even more because everybody else was kind of needing more rest, right? I had some phenomenal workouts, including multiple, multiple HIT sessions and the gym was massive there, right? Cause it was an executive apartment. I was using the gym like. So I think once I went a couple times I went three times a day. And then I came back, and I had to travel again, um and by the way, this is during this time there was that big uh uh HMPV human metapneumovirus that was everywhere, okay, so I was in Korea, massive out, you know, issues there, pneumonia, and then I went to India, same HMPV outbreak. And I felt fine, I travelled to India very shortly after, gave a talk, felt fine, but my HRV was low. It was literally like, bam. And I don't I don't think it would be a time zone change, because Korea is the same time zone and India is not … it's two hours or not 2 and a half hours, right? And so it's kind of dipped low. I felt I worked out super hard in India, too. Right? And then it just kind of went back, came back to normal. Right? And I think for some higher risk people, I am not, again for the record, I'm not, but for higher risk people who happen to be able to have that type of data could matter a lot... I think it could predict hospitalization for some people. And I think during COVID, they did do some of those studies. You know, and I think that so wearables can be immensely helpful, including for people with cardiac risks, which I'm sure there is studies done on that too. But for people who are the ones who could benefit the most, when we talk about the delta, for people who are not well and would benefit from lifesaving outcomes from this, it's even more important for the interface to be easy for them. Because for people like me that are like trying to say ultra optimized, et cetera, even if it's not an optimized interface, we'll we'll keep using it. Right? And we'll think on those terms, like, I think this would be cool if you tried this, but for some where it's like I don't know what I'm looking at. I don't care what I'm looking at. Is even important, but it's gotta be even more important for them to be aware… Yeah.

1: Erm…Oh, yeah, yeah. So with the rise of AI in everyday life. Have you incorporate incorporated the use of AI assistant tools for… to make any decisions to lead a healthier lifestyle?

2: Yeah, I think that generally I mean, I would say ChatGPT uh has been really cool to kind of uh look into insights um into, especially what some of the dynamics could mean. Um and what some of the wearable stuff means. I've I've asked for things that, you know, I've come to be aware of like, you know, one good night of sleep equals to good resting heart rate. I I've looked into generative AI in terms of how is there evidence on how aware people are of these insights in general? Um converting images to converting plots into images, you know, to see if there's a way to reinforce good outcomes visually. I think it's been cool to try to play with, um including some of our own um I I haven't say I I have I would say looked at my data with fasting to look at look, you know, kind of less intense fasting my usual fasting and then very intense fasting. And to look to see if there's actually some sort of second order effect with regards to ketone levels, which I believe could exist, right? I've seen some some sparse evidence of it, right? Because I think over-fasting in healthy people may not result in excess or pro extra prolong fasting doesn't necessarily result in highest ketones, may not be fully linear. Right? The body can also find ways to compensate a light start tapping into other energy sources, other glucose stores to drop the ketones. So from our own learnings with curate, et cetera, I poked a little bit at my own data to see if there's any evidence there.

1: I’ve two follow-up questions. How comfortable are you, for sharing of your personal healthcare data on ChatGPT and…

2: Chat(GPT), totally comfortable. Um But I but I'll be mindful that the healthcare data that I have is frankly speaking, I think our healthcare providers are not fully aware of what this data means. And I think it's part of a study where I'm putting all my data out there. I think there there is a there is a component of I'm gonna look at things that are not some sort of like high risk, like I don't have sequencing data put out there. So I think I would that and I argue that's a good gateway to get people to care about their data, to care about their health. Look at things that could be kind of indicator lights for your health, but not something where the outcome is going to terrify you or haunt you. You know, if I have a homocysteine reading that's a little high, I'm not gonna freak out. I'm gonna eat some plant-based veggies, make sure my vitamin B hang and it's gonna be fine, right? Um But by the way, my only high reading was from fasting for 48 hours, which brought right back down. Um, but I think that sharing my data has been really cool, actually. I think it's actually hopefully helped some people think about their own health. And that it's accessible. It's it's within reach. Some of these markers elective are like 30 bucks, right, and hopefully that's allowing some people to think a little bit more about what they can learn about themselves, versus being afraid of doing nothing, like, oh, I don't wanna know. And I'm, you know, I think we can we we should do a little bit more than that.

1: And my, my other follow up question is like, how do you approach the information provided by? Because there are some…

2: I will check references. I will check references, And by the way, it's pretty funny. I forgot to send this to the team, but I asked Chat GPT to give info on prior work looking at dynamics. And digital twins, et cetera. And it came up with three references. I dug deep, right, to refine searches. I dug deep, and it gave me two papers, and I was like, oh my gosh, people have totally done this. And the first who actually our worked. Okay, but it gave the wrong journal, gave the wrong journal, and uh I think one of them gave the wrong location or something, right? But I was like oh my got this what's N of 1, N of one health optimization and the title was a little bit different, I think? Wow, somebody else has done this work. You know, and then and it was said it was in like scientific reports or something like. Oh, I gotta check this in I clicked on the link in with our paper. Right? So different different journal, but so I do check and I'll dig. I'll dig and actually find the paper…

1: I know we are short on time…

3: Sorry I got one final disclosure questions, you, you know…

2: Continue we let's we can push another few minutes…

3: For the ChatGPT that you use in the AI language learning model, which version of the ChatGPT do you use?

2: Free.

3: The free one?

2: But as we know the team has a better one. And I I would prefer that for kind of this analysis is done separately for me. Yeah. So that I don't run a bunch of my own on that, and then I go to them and then there's kind of any sort of uh so that the that's why we ensure that the actual version that we're gonna talk about is done with my data but basically separate from me.

3: Cool, thank you.

1: Still on ChatGPT, like because because, like, you mentioned that you need refine search.

2: Mhmm.

1: So, I would say that the way you frame your problem is quite important for for AI tool….

2: And I play with both right? There's a way… let me give an example, like, oh, my ApoB is this I give all the good readings. You can you can almost prompt GPT to basically tell you you're super healthy, right? Or you can be like uh based upon the simple, can you do you think my based on the info, do you think my uh cholesterol will be normal? It's gonna, of course, is gonna say yes. Right? But if you were to say, but can you estimate what my cholesterol levels will be, what the range will be? Right? That I think is a more interesting question, right? So I've played around. I' played around and even so you know, I think it's quite possible to drive GPT towards just always positive stuff. Um, and so I think that, um other questions like, are there things I should look out for in addition to these versus like, here's all the supplements I take, does it look healthy, probably be like yeah, you're quite healthy. Can you please help me further address deficiencies from this?

1: Yeah. So I think like, is there any framework for the prompt that you to incorporate in your searches?

2: I haven't any frameworks yet. A good idea, though.

1: Okay, thank you.

3: Or would you, say like maintaining a sense of neutrality is important when phrasing your point.

2: It does. And I think that's why when we look at… our team looking looking at confidence intervals, looking at not I think the phrasing and the fr I think it would be um I think eventually the framework should actually be built in, like whether it's a questionnaire or something to that effect.

1: Let's wrap it up, so…

2: You missing stuff though? I don't wanna drive you to the point where you're missing.

1: Nah Uh, just like some general questions like lessons learnt, so what is the most significant but a few more significant insights. You have…

2: I think that no matter the age and we're not seeking perfection, sleep can be improved. I think if sleep is prioritized, it can be improved. It doesn't mean people will never wake up. It doesn't mean everybody's gonna get eight hours will see if they try. It doesn't mean and I talked to people, right? I'm like, I worked on my sleep, and it improved oh, but if I sleep, I always will wake up. Right? There are I think there are always ways to counter the attempt to try to improve, but I think biology generally works. If you support it.

1: What do you mean by that?

2: Like, if you want to see better. And when you wake up, you try to sleep and you wake up, okay, you sleep early. People have told me they have to actually slept earlier, and the sleep is really good. They wake up, you know it's 3 in the morning. And they feel really rested. Okay? And then I'm like, okay, cool, three you wake up and see, what do you do? And they're like, oh, I feel really rested, so I surf the phone. Okay, it's like that the story you make great strides keep going, keep going, like you you will get even better. Right? I think if a person pushes it's not gonna happen overnight. You push sleep earlier, probably get better, deep sleep, you might wake up or lose your body's not used to sleeping earlier, but then if it's like, oh, I feel rested, it's it works, I feel great. I said five hours when it was good five hours yet, but you if you if you continue to relax your body will lengthen over time. You might still wake up, but if you go back and you try to stay asleep, you will go back to sleep. Right? Aside from any sort of major medical events, brain injury, et cetera, I think biology works in that if you give it a way to shut down and you support it and you stay consistent with it, it will work. People will and can sleep better. If people I think there's this view that once you're a little bit older, a little bit older, a little bit older, you know, you have to sleep less. And yes, during different life moments, people will lose sleep, but you can come back. It does come back. I think that uh this is this is maybe more obvious, but when you try to sleep well and you … I would say sleep is probably the most important aspect of what has helped me feel better and feel good during this time. I would say the diet has been number two. I think the clean eating and its impact on my numbers is pretty evident. At my age, I don't think it's luck. Okay, I don't think it's luck at my markers are pretty good. Um, and I think fitness plays a third ranking supportive role in that you know. Um but uh for me, I think another lesson note is for me. I have found the kind of intermittent fasting to be a fairly straightforward way for me now to… uh I think it works for regulating my metabolic health. I think it's clear. Um with what my implied A1c is with my metabolic switching, with my speed ketosis, um I think it's and I have data on that, by the way, right? On days where I'm doing three meals a day, I with the exception, I think one time where I flipped in quickly the ketosis, I do not metabolically switch. But when I'm doing my usual routine, I have done spot checks and I have metabolically I happen to flip. Right? And so I think that part has been important. And it appears that it has no negative impact that we know of on my gut health, as well.

1: And last question for me, like, is there any aspects of the study that you like to explore in for the future studies?

2: Yep, um I would like to now look at more of the impact on my cognitive health. There's some implication with homocysteine as a marker. Um, but there are other markers I would like to look at, and then, of course, I would like to scale up uh performance hence VO2 max. Uh we will look at that um and then and again, cognitive yeah, because there's implications that ketosis, et cetera, good for, you know, mental clarity, I don't know. Um there are implications that certain markers will go up that really support cognitive health we'll see. Um, but uh, I think from the first part, inflammation, cardio metabolic health, uh some implications of neurological health with the homocysteine uh gut health, sleep hygiene, sleep performance. I think we really were quite comprehensive, but I think cognitive is next. Others?

1: No. I'm good.

2: You sure? Yeah, yeah, yeah. I would sum it up by saying in this quest for a health span, et cetera, um, people don't have to be like biohacking all day and doing all kinds of hardcore stuff, but people should try to do something. Try to do something that works. That's a little bit sustainable. That's it.

1: Okay.

2: Because we don't have to tend towards disorder. We don't. Right? Um there's a notion that people will their blood sugar will always tend higher, that insulin resistance is a part of life, that or not insulin resistance, but increasing resistance is a part of life, that sleep getting worse over time is a part of life. Doesn't have to be that way.

1: Thank you.

2: Thank you.

1: Thank you so much. All right.

(End of Interview 3)
